# Interpreting variants associated with congenital heart disease with a regulatory map of the human fetal heart

**DOI:** 10.1101/2024.11.20.24317557

**Authors:** X. Rosa Ma, Stephanie D. Conley, Maggie S. Chen, Michael Kosicki, Yifan Lu, Danila Bredikhin, Ran Cui, Maya U. Sheth, Wei-Lin Qiu, Dulguun Amgalan, Sijie Chen, Robin M. Perelli, Neil Slaven, Michael T. Montgomery, Sarah Fan Xia, Zhainib Amir-Ugokwe, Charu Balamurugan, Steven Tran, Soumya Kundu, Helen Y. Kang, Xiangmeng Shawn Cai, Chad J. Munger, Yannick C. Lee-Yow, Tony Zeng, Yanyan Alexandra Mo, Julia Vahey, Tri Nguyen, Chloe V. McCreery, Sidney Vermeulen, Lauren Duan, Katherine Dang, Jan-Renier Moonen, Oriane Matthys Rubio, Shinwan Kany, Siavash Zamirpour, John DePaolo, Arun Padmanabhan, Birth Defects Research Laboratory, Jeffrey Olgin, Scott M. Damrauer, Robin Andersson, Mingxia Gu, James R. Priest, Thomas Quertermous, Marlene Rabinovitch, Kristy Red-Horse, Axel Visel, Len A. Pennacchio, Xiaojie Qiu, Anshul Kundaje, Ian A. Glass, Casey A. Gifford, James P. Pirruccello, William R. Goodyer, Jesse M. Engreitz

## Abstract

Congenital heart defects (CHD) arise in part due to inherited genetic variants that alter genes and noncoding regulatory elements in the human genome. These variants are thought to act during fetal development to influence the formation of different heart structures. However, identifying the genes, pathways, and cell types that mediate these effects has been challenging due to the diversity of cell types involved in heart development as well as the superimposed complexities of interpreting noncoding sequences. As such, understanding the molecular functions of both noncoding and coding variants remains paramount to our fundamental understanding of cardiac development and CHD. Here, we created a gene regulation map of the healthy human fetal heart across developmental time and applied it to interpret the functions of variants associated with CHD and quantitative cardiac traits. We collected multiomic data from 734,078 single cells sampled from 41 fetal hearts spanning post-conception weeks (PCW) 6 to 22, enabling the construction of gene regulation maps in 90 cardiac cell types and states, including rare populations of cardiac conduction system (CCS) cell types. We find that many such risk variants, both coding and noncoding, map onto gene regulatory networks in valve cells. In an unbiased analysis of 90 cardiac cell types and states, valvular interstitial cells (VICs) show the strongest enrichments for known CHD genes. Nine CHD genes are linked to common noncoding variants associated with other valve diseases or traits via transcriptional enhancers in VICs. CRISPRi experiments show that multiple CHD genes have cell-autonomous effects in VICs, including regulating the expression of other CHD genes and genes involved in cell migration and developmental patterning. Finally, we characterize a CHD-associated noncoding variant whose activity is specific to discrete structures within the heart, illuminating how such variants can impact specific aspects of heart structure and function. Together, these results suggest that genetic risk for many subtypes of CHD is influenced in part by genetic regulation of valvular cells, expanding our understanding of the cell types responsible for CHD beyond the working cardiomyocyte. This regulatory map of the human fetal heart provides a foundational resource for understanding cardiac development, interpreting genetic variants associated with heart disease, and discovering targets for cell-type specific therapies.

## Introduction

The formation of the human heart during fetal development is driven by the emergence and coordination of a vast array of different cell types^1–3^. Each of these cell types carries out distinct architectural or regulatory functions in forming the heart chambers, valves, conduction system, and blood vessels. These dynamic processes are encoded by complex regulatory programs in the human genome, involving different combinations of genes regulated by up to millions of noncoding regulatory elements and transcription factor (TF) binding sites that are yet to be fully identified^4^.

DNA variants that disrupt these regulatory programs can lead to CHD^2,5^, which are the most common birth defect and affect nearly 1% of live births^6^. Genetic studies of patients with CHD have now discovered thousands of variants in both protein-coding and noncoding regions of the genome^7–9^. Studies of such coding variants have revealed important insights into the molecular regulators of heart development and identified a growing catalog of high-confidence CHD genes that currently help to guide genetic diagnosis^1,5,7–12^. Noncoding variants are far more difficult to interpret, but some have recently been associated with CHD through genome-wide association studies (GWAS)^13–16^ or identified through molecular analysis of *de novo* variants^4,17–19^.

Despite this progress, the genetic contributions to most CHD cases remain to be identified^2,5^, and key questions about the genes, pathways and cell types that contribute to CHD remain to be addressed:

1. Which cell types contribute to the genetic etiology of different subtypes of CHD? CHD is a heterogeneous class of diseases, with subtypes involving different specialized structures of the heart, such as valve defects and malformations of the outflow tract (OFT). Early studies focused on the heart muscle cell, or cardiomyocyte, following the identification of CHD-causing mutations in TFs that affect myocardial specification and/or function, such as *NKX2-5*, *TBX5*, and *GATA4*^1,2,20–23^. We now appreciate that other cell types are likely to have roles in the genetic etiology of CHD, such as contributions of the cardiac neural crest lineage to malformations of the OFT^24,25^ and an emerging role for endocardial cells in hypoplastic left heart syndrome^26^. The development of single-cell technologies, and our accompanying awareness of the vast diversity of cell types and states in the human heart, provides a new opportunity to assess which cell types contribute to the genetic etiology of which subtypes of human CHD.
2. How do inherited noncoding variants influence risk for CHD? A primary challenge in interpreting the functions of noncoding variants has been the lack of appropriate regulatory maps to connect these variants to their target genes and cell types. Previous studies have proposed roles for *de novo* noncoding variants linked to CHD in cardiomyocytes or endothelial cells and identified a handful of individual variants that affect enhancer activity or alternative RNA splicing^4,17–19^. More recently, GWAS have identified inherited noncoding variants associated with CHD^13–16,27–29^ and related quantitative measurements of cardiac morphology^30–39^. To effectively leverage these recent genetic datasets to learn about genetic modifiers of CHD risk, severity, or subtype, we need to identify which variants regulate which enhancers, their corresponding target genes, and in which of the many cell types of the developing heart these gene regulatory interactions occur.

To answer these and other outstanding questions in CHD research, a map of gene regulation that captures the diverse cell types and cell states that drive human fetal cardiogenesis is essential. Recent studies have begun to collect data on gene expression or chromatin accessibility during heart development in mouse models^40–44^ or from human fetal tissue^4,45–52^. However, existing datasets are still limited in cell number, sequencing depth, and developmental timepoints, precluding the application of key computational models to build a gene regulation map across cell types and states. No previous study has analyzed noncoding variants associated with CHD or quantitative cardiac traits in combination with a chromatin atlas of the fetal heart. Far deeper single-cell data, together with new computational models, is needed to interpret the functions of genetic variants associated with heart structure and function.

Here, we create a gene regulation map of the human fetal heart using a multiomic atlas of the human fetal heart and combine it with recent human genetics datasets to reveal new insights into the genetic, molecular, and cellular etiology of CHD. First, we collect multiome (scRNA-seq and scATAC-seq) data from 734,078 cardiac cells from 41 structurally normal hearts between 6 and 22 PCW of human fetal development. This cell atlas significantly surpasses existing datasets, both human and mouse, in terms of cell number, sequencing depth, and the developmental windows captured. We identify 90 cardiac cell types and cell states with sufficient depth, and, in each, apply predictive models to identify thousands of novel enhancers, TF binding sites, and downstream target genes. By applying this map to interpret coding and noncoding variants associated with CHD and quantitative traits, we find that high-confidence genes harboring rare coding variants in CHD patients are most strongly enriched for high expression in VICs and cardiac fibroblasts—two cell types whose roles in CHD have been underexplored. We nominate and experimentally validate enhancers containing functional noncoding variants, whose activities are specific to particular cell types or locations in the heart. Certain genes and pathways in VICs are affected by both rare coding variants associated with CHD and common noncoding variants associated with structural traits. CRISPR perturbations confirm that multiple CHD genes regulate key developmental expression programs in VICs. Together, these results identify new cell types and pathways important for genetic regulation of heart development, provide an enduring resource for interpreting the functions of genetic variants for heart diseases, and demonstrate a generalizable framework for mapping the impact of genomic variation on function from tissue atlases.

## Results

### A multiomic atlas of the human fetal heart

We collected single-cell RNA and ATAC-seq data (10x Multiome) from 41 fetal heart samples spanning 6 to 22 PCW (**Fig. 1A; Fig. S1; Table S1**). These samples included 21 hearts from 6 to 9 PCW, a developmental stage that coincides with key events such as the formation of the OFT, valves, and conduction system. For 6 through 9 PCW, we sampled cells from the entire heart (**Methods**), and profiled sections from the larger 10 through 22 PCW samples (**Table S2**). After quality control and filtering, we obtained a total of 734,078 single cells (range: 370-94,555 per heart), with an average of 1,682 RNA transcripts and 5,953 ATAC fragments per cell.

**Figure 1.**
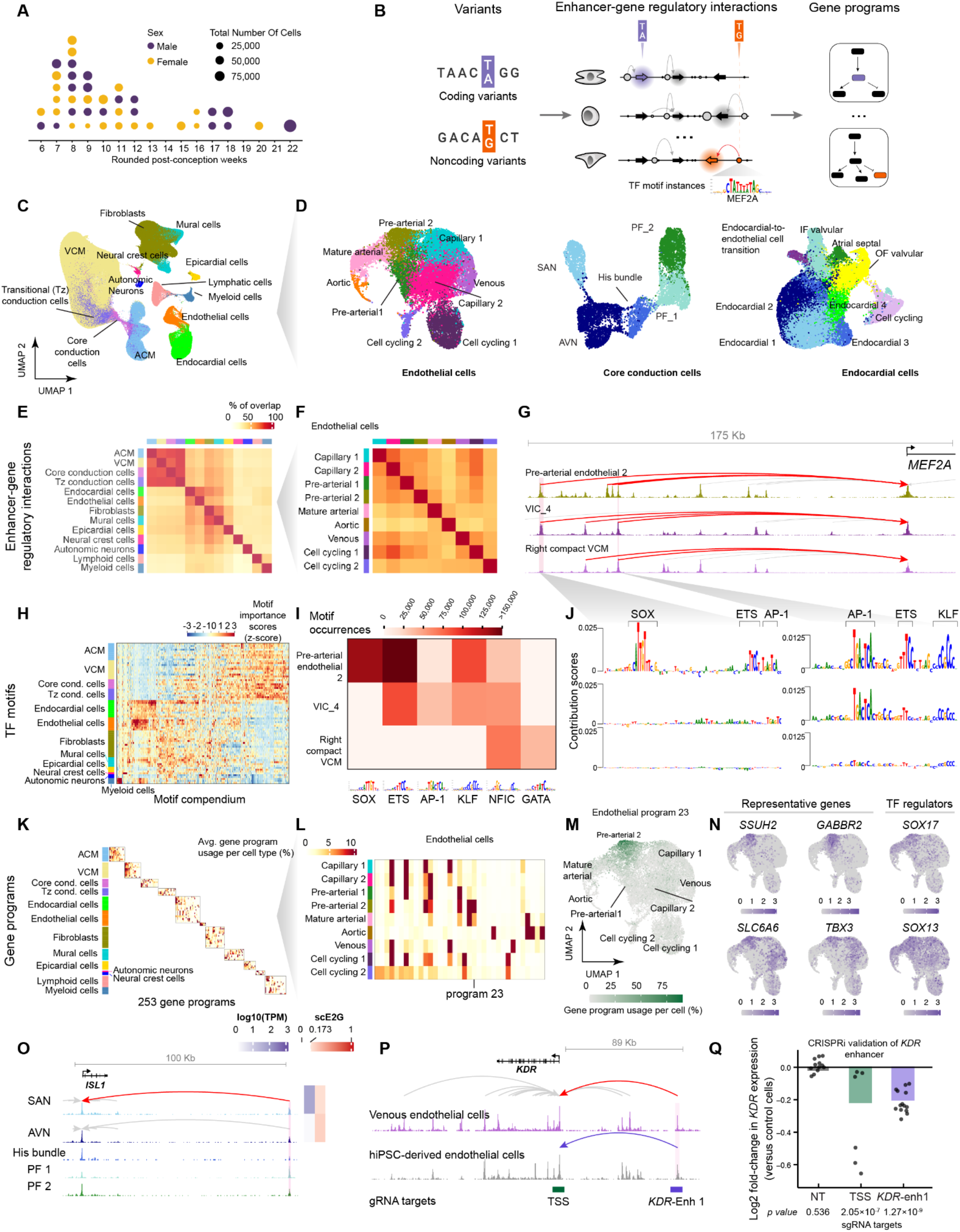
A gene regulation map of the human fetal heart. **A.** Single-cell multiomic data (10x Multiome) was collected from 41 hearts spanning PCW 6 to 22. Each dot represents one heart, plotted by rounded PCW; dot color indicates biological sex (purple, male; yellow, female) and dot area is proportional to the total number of cells recovered from that sample. **B.** Multiome data was used to create a gene regulation map, linking genetic variants to regulatory elements (enhancers), their target genes, and associated gene programs in each of 90 cell types with sufficient depth. The schematic illustrates the components of this map and how it can be used to interpret the molecular and cellular functions of coding (purple) and noncoding (orange) variants associated with disease. The middle panel shows cell-type-specific enhancer-gene interactions: arrows represent genes, with highlighted arrows indicating expression in a particular cell type; circles represent predicted enhancers, with size indicating activity level, and arcs show predicted enhancer-gene interactions. Coding variants can be annotated based on the cell types in which the gene is expressed (purple). Noncoding variants can be annotated based on the cell types in which the enhancer is active, and the target gene(s) of that enhancer (orange). Right panel depicts gene programs, which are defined as sets of genes that are co-expressed across single cells that are likely to have related functions. Both coding and noncoding variants can be annotated based on how their target genes work together in gene programs (colored genes). **C.** Uniform manifold approximation and projection (UMAP) embedding of scRNA-seq of all major cell types. ACM: Atrial cardiomyocytes. VCM: ventricular cardiomyocytes. Tz: transitional. **D.** UMAP embeddings of endothelial cells (excluding lymphatic endothelial cells), cardiac conduction cells, and endocardial cells. AVN: atrioventricular node. IF: inflow. OF: outflow. PF: Purkinje fibers. SAN: sinoatrial node. **E.** Global similarity of predicted enhancers across major cell types. Color scale: Percentage of enhancer overlap between pairs of major cell types, with the total number of enhancers in the major cell type on the y-axis as the denominator (see **Methods**). Tz conduction cells: transitional conduction cells. **F.** Global similarity of predicted enhancers across endothelial cell types. Color scale: Percentage of enhancer overlap, as in **E**. **G.** Example locus: *MEF2A* is regulated by distinct combinations of enhancers in endothelial cells, VICs, and VCMs. Signal tracks: pseudobulk scATAC-seq within each cell type. Red arcs: Elements predicted by scE2G to act as enhancers to regulate *MEF2A*. Gray arcs: Elements predicted to regulate other genes. Coordinates (hg38) of highlighted enhancers: chr15:99425061-99426373 (left), chr15:99454858-99455373 (right). **H.** Compendium of TF motifs and predicted effects on chromatin accessibility across 90 cell types, as learned by ChromBPNet and TF-MoDISco. Color scale: Z-score normalized effects of predicted motifs learned as average differences in predicted accessibility into 100 random sequences in each cell type. Core cond. cells: core conduction cells. Tz cond. cells: transitional conduction cells. **I.** Predicted occurrences of selected TF motifs in cell types relevant to *MEF2A* enhancers (see panel **G**). Color scale: motif occurrences in the corresponding cell type. **J.** ChromBPNet contribution scores highlight TF motif instances with cell-type-specific contributions to chromatin accessibility at two selected enhancers for *MEF2A*. Height of each nucleotide in the genomic sequence represents the predicted importance of that nucleotide to chromatin accessibility (by DeepLIFT). Coordinates (hg38) of the two example regions: chr15: 99425308-99425388 (left), chr15:99455071-99455122 (right). **K.** Expression patterns of 253 gene programs across cell types. Gene programs were inferred using cNMF in groups of related cell types. Color scale: average gene program usage per cell type. **L.** Average expression of endothelial cell programs in each of the fine-grained endothelial cell types. **M.** Projection of Endothelial program 23 expression onto the endothelial cell UMAP. Color scale: gene program usage per cell. **N.** Feature plots illustrating the expression of known arteriolar endothelial marker genes identified in Endothelial program 23 and the expression of predicted TF regulators of Endothelial program 23. Color scale: normalized gene expression level. **O.** scE2G predictions and normalized scATAC-seq signals for *ISL1*. Predicted enhancers and the *ISL1* promoter are highlighted in shaded regions. Arcs represent the predicted enhancer-gene regulatory interactions. Side color bars show the transcripts per million (TPM) expression of the target genes in each cell type (blue) and the corresponding scE2G scores (red, and white for scores below the threshold). SAN: Sinoatrial node; AVN: Atrioventricular node; PF: Purkinje fibers. **P.** scE2G predictions and normalized scATAC-seq signals for *KDR*. The gray and red arcs represent the predicted enhancer-gene regulatory interactions. Bottom: boxes represent elements tested using CRISPR perturbation; TSS: transcription start site. **Q.** *KDR* enhancer (hg38: chr4:55214146-55214646) regulates *KDR* expression in hiPSC-derived endothelial cells, as measured by CRISPRi TAP-seq^75^. Single-guide RNAs (sgRNA) targeting the enhancer significantly decreased *KDR* expression compared to non-targeting (NT) sgRNA controls, based on SCEPTRE’s^76^ conditional resampling method with Benjamini-Hochberg correction. Each data point represents one sgRNA. N for sgRNAs: non-targeting=12, *KDR* transcription start site (TSS)=6, *KDR* enhancer=14: NT: non-targeting.

Clustering and annotation yielded 13 major cell types (**Fig. 1C; Fig. S2)**, which were further annotated to a total of 101 cell populations (hereafter, “cell types”). For instance, we identified 9 endothelial cell populations, consistent with findings from previous studies^42,53,54^(**Fig. 1D**). We labeled cell types using canonical cell type markers as well as genes identified in recent single-cell and spatial transcriptomic studies^41,42,48,49,53^ (see **Methods; Figs. S3-8**). Our atlas captures rare cell populations, including neural crest cells and cells from major substructures of the conduction system (sinoatrial node (SAN), atrioventricular node (AVN), His bundle and Purkinje fibers (PF); **Note S1, Figs. S8-9, S11**), and assigns 37 cell types to likely subanatomic locations. These locations were confirmed by mapping onto independent spatial transcriptomics data on heart cross-sections^48^. For example, this map distinguishes endocardial cells of the atrial septum and inflow (IF) and outflow (OF) sides of valves, based on expression of marker genes and spatial mapping (**Fig. 1D; Figs. S4, S9**).

### A gene regulation map of the human fetal heart

Leveraging this deep dataset, we constructed a gene regulation map describing the relationships between genetic variants, TFs, enhancers, target genes, and gene programs (**Fig. 1B**). To do so, we applied a suite of predictive models, some of which we have previously developed and applied with the ENCODE Project and IGVF Consortium^55^. These models have been shown to improve inferences of causal variants and genes for disease^56–59^, and in combination provide high specificity for identifying noncoding variants associated with cardiac traits (**Fig. S14**). Importantly, these models require a depth of sequencing that is not available in previous datasets (minimum 3 million ATAC fragments per cell type)^56,57^. We sequenced our multiomic atlas to a depth that enabled constructing cell type-specific gene regulation maps in 90 cell types, including cell populations with frequencies in our dataset as low as 0.06% (**Fig. S10**). Together, this atlas allows us to interpret the effects of genetic variants at far greater cellular resolution than previous chromatin atlases of the human fetal heart^4,45,60^, which include 6- to 24-fold fewer cells, resolve the fetal heart into at most 20 coarse-grained cell populations, and therefore lack the resolution to distinguish key cell types such as VICs and cardiac conduction cells.

Here, we briefly describe each component of this gene regulation map, and in subsequent sections combine all components in integrative analyses:

### Linking enhancers to target genes

To identify candidate enhancers and link them to their target genes in specific cell types, we applied scE2G, a supervised logistic regression model trained to predict the effects of CRISPR perturbations from single-cell multiome data^56^. We have previously shown that scE2G shows state-of-the-art performance at predicting the results of CRISPR enhancer perturbations, linking eQTL variants to their target genes, and linking GWAS variants to known target genes^56^. Applied to our fetal heart atlas, scE2G identified on average 41,267 enhancer-gene regulatory interactions per cell type (**Table S5**). As expected, closely related cell types shared more enhancers than distantly related cell types: for example, atrial and ventricular cardiomyocytes (VCM) shared 80% of their enhancers, while lymphocytes and neural crest cells shared only 21% (**Fig. 1E**). Within each major cell type, we observed a higher degree of enhancer overlap between cell types that are closely related in function or along a differentiation trajectory. For example, over 50% of enhancers in pre-arterial endothelial cells were also active in mature arterial endothelial cells (**Fig. 1F)**. Within conduction system cells, nearly 60% of the enhancers were shared between nodal cells (SAN and AVN), while only 48% of the enhancers were shared between SAN and more functionally distinct His bundle cells (**Fig. S8D**). For individual genes, scE2G identifies cell-type specific sets of enhancers that contribute to gene expression. For example, *MEF2A*, a core TF expressed in many cell types in the developing heart, has distinct sets of enhancers in endothelial cells, VICs, and VCMs (**Fig. 1G**).

### Identifying TF binding sites and predicting effects of noncoding variants on accessibility

To identify candidate TF binding sites and predict the effects of noncoding variants, we applied ChromBPNet, DeepLIFT, and TF-MoDISco^57,61,62^ — a convolutional neural network method and accompanying interpretation tools. This framework predicts base pair-resolution chromatin accessibility data (pseudobulked per cell type) from DNA sequence, identifies TF motif instances that drive accessibility, and predicts the effects of noncoding variants on chromatin accessibility. We have shown that this approach performs well at identifying TF binding sites^63^, predicting effects of CRISPR edits on gene expression^64^, and identifying functional GWAS variants^57^. Per cell type, we identified 19*–*39 important TF motifs and 340,000*–*760,000 high-confidence TF motif instances (**Table S18-20**), with 516 consensus motifs combined across cell types (**Figs. 1H,I**). Predicted TF motifs included known associations such as SOX and ETS motifs in pre-arterial endothelial cells, and GATA motifs in VCMs and the SAN (**Fig. S12**). Individual regulatory elements often showed distinct predicted TF motif contributing to accessibility in different cell types (*e.g.*, **Fig. 1J**)^4^.

### Linking genes to gene programs

To discover sets of co-expressed genes that may work together in similar biological pathways, we learned gene programs (“programs”) by applying consensus non-negative matrix factorization (cNMF)^59^ to the transcriptome data and annotating TFs with correlated motifs in the ATAC data (**Tables S6-8**, see **Methods**). cNMF identifies sets of genes that are co-expressed across single cells, and has been shown to learn diverse types of biologically coherent pathways, including metabolic pathways, developmental trajectories, and cell identity programs^59,65^. We applied cNMF to groups of major cell types and identified 253 total gene programs representing a wide array of biological processes (**Figs. 1K,L**). For example, in endothelial cells, Endothelial program 23 is expressed in pre-arterial endothelial cells (**Fig. 1M**), includes genes previously observed to be expressed in arteriolar endothelial cells such as *SSUH2*, *GABBR2*, *HECW2*, and *SLC6A6*^42^, and is regulated by TFs with known roles in the developing artery such as SOX17, SOX13 and MEF2A (**Fig. 1N**). Endocardial program 33 (“Atrial septal endocardial cell identity”) is exclusively and uniformly expressed in atrial septal endocardial cells and includes known marker genes such as *LRRC4C*, *LSAMP*, *ALCAM*, and *NEGR1*. This program was predicted to be regulated by GATA6, a TF where loss-of-function mutations cause a spectrum of CHDs including atrial septal defect (ASD)^66,67^, and NFATC2, a TF essential for atrial septum morphogenesis^68^. Endocardial program 29 (“WNT signaling in valvular endocardial cells”) is expressed in a subset of IF-side valvular endocardial cells (“IF valvular endocardial cells”), includes WNT signaling pathway genes such as *WNT4*, *WNT2*, and *WNT9B* and is regulated by flow-responsive TFs known to regulate WNT signaling, KLF2 and KLF4^69^ (**Tables S7-8**). Notably, this unsupervised framework successfully captures discrete gene programs and their upstream regulators even within exceptionally rare lineages (**Note S1, Fig. S13)**.

We compared predictions from this map to functional genomics studies of cardiac enhancers. Our gene regulation map includes 12 previously validated enhancers implicated in embryonic stem cell-to-cardiomyocyte differentiation^70^ and the regulation of two key cardiac genes *ISL1*^71^ and *ACTN2*^72^, as well as 151 out of 179 enhancers previously observed to have activity in the murine heart at E11.5 using *in vivo* reporter assays^73,74^ (**Fig. 1O, Table S10**). For additional validations of our findings, we used CRISPRi to perturb a predicted enhancer for *KDR,* a gene essential for vascular development, in endothelial cells differentiated from human induced pluripotent stem cells (hiPSC), and observed that this enhancer indeed regulates *KDR* expression (**Fig. 1P,Q**).

Together, this gene regulation map of the human fetal heart describes how DNA sequences in our genome regulate the expression of particular genes, and how those genes work together in cell-type specific pathways. This map provides a rich resource for understanding the regulatory logic of heart development across cell types and for interpreting DNA variants that affect heart structure and function.

### Expression of genes associated with congenital heart defects

We applied this gene regulation map of the human fetal heart to interpret the functions of genetic variants associated with CHD. A growing catalog of genes have been implicated in various subtypes of CHD through analysis of rare or *de novo* coding variants in families or cohort studies^2,8,9^. A smaller number of noncoding variants have been associated with CHD through GWAS (see below)^13–16^. For both sets of variants, the cell types of action remain largely unknown. With our more complete atlas of the human fetal heart, we first examined genes associated with CHD through rare coding variation and tested whether they were preferentially expressed in certain cell types and states.

As a positive control, we first analyzed known genes for other cardiovascular diseases where the critical cell types are already established (**Table S12**). As expected, genes associated with cardiomyopathies, diseases of the heart muscle tissue, were significantly enriched for high expression within cardiomyocytes (up to 94-fold). Similarly, known genes underlying increased risk for developing aortic aneurysms were enriched in smooth muscle cells (SMC), the most abundant cell type in the aortic wall^77,78^ (up to 92-fold; **Fig. 2A, Fig. S16, Tables S11-12**).

**Figure 2.**
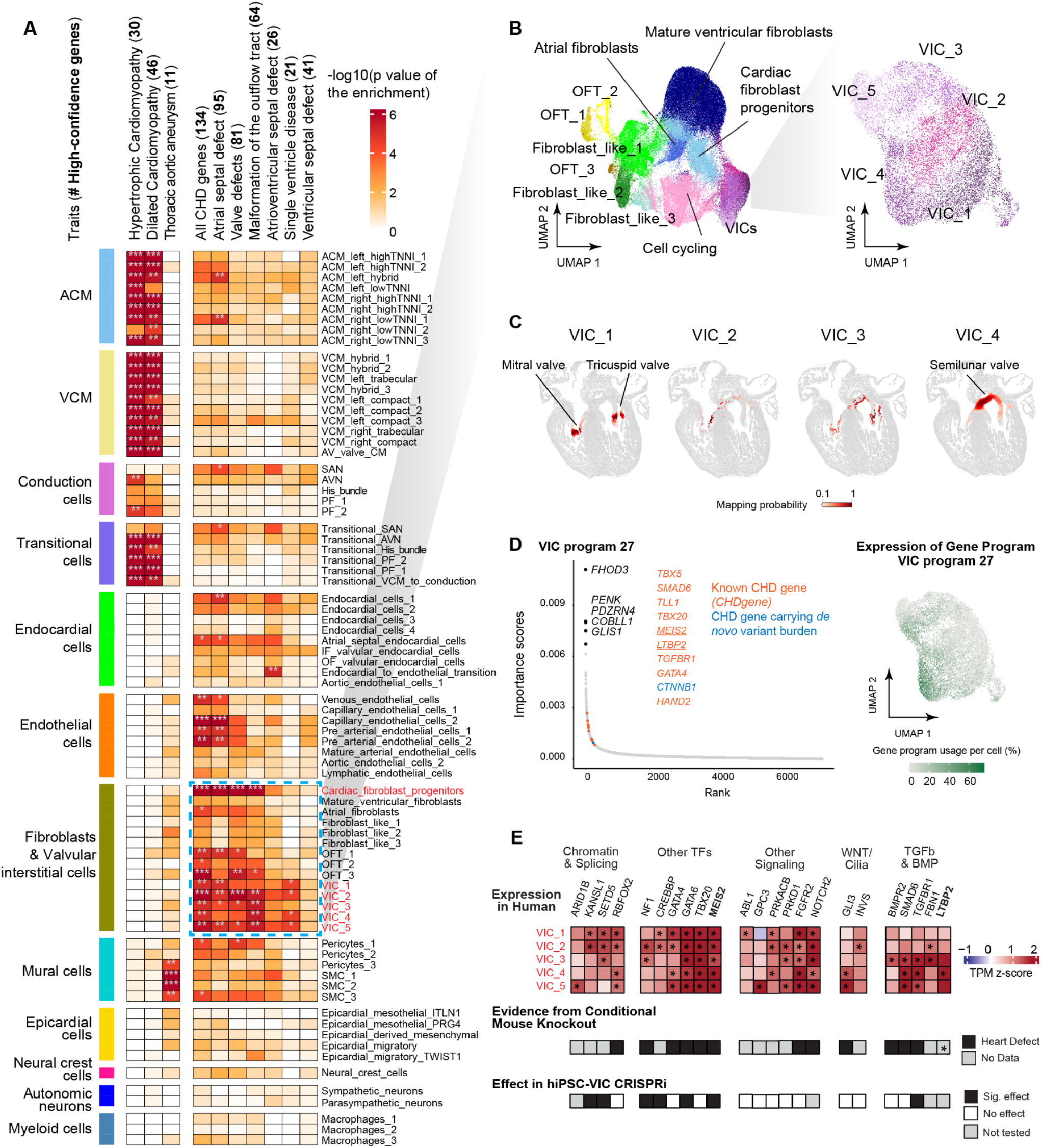
Congenital heart disease genes are enriched in valvular interstitial cells. **A.** Enrichment for cardiovascular disease and CHD genes in cardiac cell types (top 300 expressed genes per cell type, ranked by *z* score across cell types). Color scale: −log_10_-transformed p-values. One-sided Fisher exact test with Bonferroni correction (*P_Bonferroni < 0.05, **P_Bonferroni < 0.01, ***P_Bonferroni < 0.001). # High-confidence genes: the number of CHD genes expressed in any cell type in the heatmap (TPM>1). Only 134 of the 137 curated CHD genes were analyzed (FOXH1, GDF1, and ZIC3 were undetected). **B.** UMAP embedding of scRNA-seq of all fibroblast cells and UMAP embedding of scRNA-seq of VICs. OFT: outflow tract. **C.** Spatial mapping of VIC subpopulations onto a cross-sectional heart image from^48^. Mapping was restricted to cells from PCW 12, matching the age of the heart profiled with spatial transcriptomics. Color intensity indicates the probability for each cell in the spatial dataset mapping to the given cell type from our multiome atlas. VIC_5 was excluded due to an insufficient number of cells at 12 PCW. **D.** Ranked importance scores of genes associated with Fetal VIC program 27; The top 300 genes by importance scores in Fetal VIC program 27 were considered as program genes. Black: Top 5 genes associated with Fetal VIC program 27; Orange: high-confidence CHD genes from CHDgene in the top 300 genes^9^; Blue: CHD genes carrying a significant burden of heterozygous *de novo* or rare transmitted damaging variants^7^. *TBX5*, *SMAD6*, and *GATA4* are both high-confidence CHD genes and genes carrying two or more *de novo* variants. Inset: Projection of Fetal VIC program 27 expression on the VIC UMAP, showing expression in VIC_4 and other VIC populations. Color scale: gene program usage per cell. Underlined genes are discussed in the text or Note S3. **E.** Top: Z-score normalized TPM values of 23 CHD genes highly expressed in VICs. Bold genes (*MEIS2* and *LTBP2*) are discussed in the text or Note S2. *: the gene is considered to be highly expressed in the corresponding cell types (see **Methods**). Middle: Summary of experimental evidence from mouse knockouts that a given gene may act in VICs or cardiac fibroblast progenitors to lead to heart defects (see also **Table S13**). Black squares indicate that a heart defect is observed in conditional knockout in the epicardial, endocardial, and/or neural crest lineage. Gray squares indicate that such conditional knockouts have not been reported. *: For *LTBP2*, heart defects are observed in a constitutive knockout. Bottom: Black squares indicate genes that affect gene programs when perturbed with CRISPRi in hiPSC-derived VICs (see Fig. 4).

For CHD, we examined a curated list of 137 high-confidence genes (hereafter, “CHD genes”) for six subtypes: ASD, ventricular septal defects (VSD), atrioventricular septal defects (AVSD), functional single ventricles, malformation of the OFT, and valve defects (**Table S11**), based on CHDGene^9^, which compiled genes with three or more case reports for syndromic or isolated CHD.

Overall, CHD genes were broadly expressed across many cell types in the heart (**Fig. S15**). While most genes for cardiomyopathies or aortic aneurysms were expressed only in specific cell types, more than 60% of CHD genes were expressed (TPM>10) in at least one cell type in all major cardiac cell type groups (see **Methods**). These broad expression patterns might help to explain why many CHD mutations are associated with many different subtypes of heart defects.

Despite these broad expression patterns, CHD genes did show higher expression levels in some cell types than in others. We identified 20 cell types with significant enrichment for genes associated with at least one subtype of CHD (Bonferroni-adjusted *P* < 0.05) (**Fig. 2A, Fig. S16, Table S12**). For example, the ASD-associated gene panel showed a significant 6-fold enrichment in ACMs, driven by the high expression of 10 out of 95 known disease genes, including *TBX5* and *MYH6* (*q* = 0.007). Similarly, we observed a 5.5-fold enrichment of ASD-associated genes within atrial septal endocardial cells (*q* = 0.045), where nine known disease genes, including *NR2F2* and *FOXP1*, were highly expressed. Genes associated with valve defects were significantly overrepresented in VICs (up to 9-fold, *q* = 2 x 10^-5^). Genes associated with single ventricle disease, which involve valve lesions together with other defects, were also enriched within VICs, to an even higher degree (up to 16-fold, *q* = 0.02; **Table S12**).

Thus, the patterns of expression of high-confidence CHD genes suggest that different cell types, beyond just cardiomyocytes, may play an important role in the development of certain subtypes of CHD.

### CHD genes in VICs

Out of all cell types, VICs and cardiac fibroblast progenitors showed the strongest enrichments for high-confidence CHD genes. These mesenchymal cell types are known from prior animal studies to have important roles in heart development, but their direct involvement in the genetic etiology of CHD in humans remains underexplored.

We focused here on VICs (see **Note S2** for discussion of cardiac fibroblast progenitors)^79^. Our atlas includes three populations of VICs (VIC_1-3) expressing markers of the valve free segments, and two valve-related mesenchymal cell populations expressing markers of mesenchymal cells located in intervalvular fibrous tissue towards the valve roots (VIC_4 and VIC_5). VIC_4 expresses the neural crest-derived mesenchymal cells marker *PENK*^49^, and VIC_5 expresses genes primarily observed around the semilunar valves^49^. All five populations were spatially mapped to the valves (**Fig. 2C, Fig. S5C**). Together, VICs were significantly enriched for genes for four of six subtypes of CHD (ASD, valve lesions, malformations of the OFT, and single ventricle disease; 5-16-fold enrichment, *q* <0.045; **Fig. 2A, Table S12**).

Altogether, 23 high-confidence CHD genes were preferentially expressed in VICs. Twelve have previously been shown to lead to heart defects in mouse models through conditional knockouts consistent with functions in VICs (**Fig. 2E, Note S3, Table S13**). All 23 genes are expressed throughout human heart development (from 4-22 PCW); in addition, expression increases during mouse endocardial-to-mesenchymal transition (EndMT) (**Fig. S24)**. For example, *LTBP2* is the CHD gene most specifically expressed in VICs versus other cell types (**Fig. S15**), and is particularly highly expressed in VIC_4 (**Fig. 2E**). *LTBP2* encodes the extracellular matrix protein, latent-transforming growth factor beta-binding protein 2, that forms part of the latent TGF-β complex and interacts with microfibrils. Loss-of-function variants in *LTBP2* have been observed in patients with bicuspid aortic valve (BAV), transposition of the great arteries, aortic stenosis, pulmonic stenosis, and mitral valve prolapse^80,81^. Consistent with these observations, *Ltbp2* knockout in mice is embryonic lethal^82^, and mouse knock-in of a disease-associated missense variant leads to mitral valve prolapse^83^. In our atlas, *LTBP2* is part of a VIC gene program (Fetal VIC program 27, “VIC migration and TGF-β signaling”) that includes eight other high-confidence CHD genes (*GATA4, MEIS2, TBX20, TBX5, TGFBR1, TLL1, HAND2, SMAD6*) (**Fig. 2D)**. Four genes (*TBX5*, *SMAD6*, *GATA4*, *CTNNB1*) in this program carry a significant burden of heterozygous *de novo* or rare transmitted damaging variants in a large-scale CHD case-control cohort^7^. Consistent with the necessity of these genes in normal heart development, loss-of-function mouse models for *Gata4, Meis2, Tbx20, Smad6,* and *Tgfbr1* also result in valve defects and/or malformations of the OFT (**Fig. 2E, Note S3**).

Notably, 60 high-confidence CHD genes exhibited expression profiles in VICs that correlated with post-conceptional age (**Fig. S18**). For example, *TBX5*, a top gene in program 23 (“Early VIC_4 signature”), is enriched in chronologically younger cells, consistent with its role in early endocardial cushion development^84^. Conversely, *ELN*, a top gene of program 25 (“Late VIC_4 signature”), shows increased expression in later timepoints, mirroring the previously observed rise in elastin (*ELN*) production during late embryogenesis and early postnatal development^85^. Together, our analysis indicates that many high-confidence CHD genes are highly expressed in VICs, often as part of co-regulated gene programs.

### Variants for CHD and valve traits impact shared VIC genes and programs

To further explore the role of VICs in heart development and disease, we examined common noncoding variants associated with valve diseases and quantitative traits, including new GWAS we recently conducted to analyze variants associated with aortic valve function^86^. Previous genetic studies have noted that certain GWAS signals for acquired valve diseases are located near known CHD genes. For example, a noncoding variant near *LTBP2*, one of the CHD genes described above, is associated with mitral valve prolapse^87^ (**Fig. 3A**). However, the specific causal enhancers or cell types for these loci have not yet been identified. Accordingly, we applied our regulatory map to test whether noncoding variants associated with valve traits might act in VICs to regulate known CHD genes and other genes in similar pathways.

**Figure 3:**
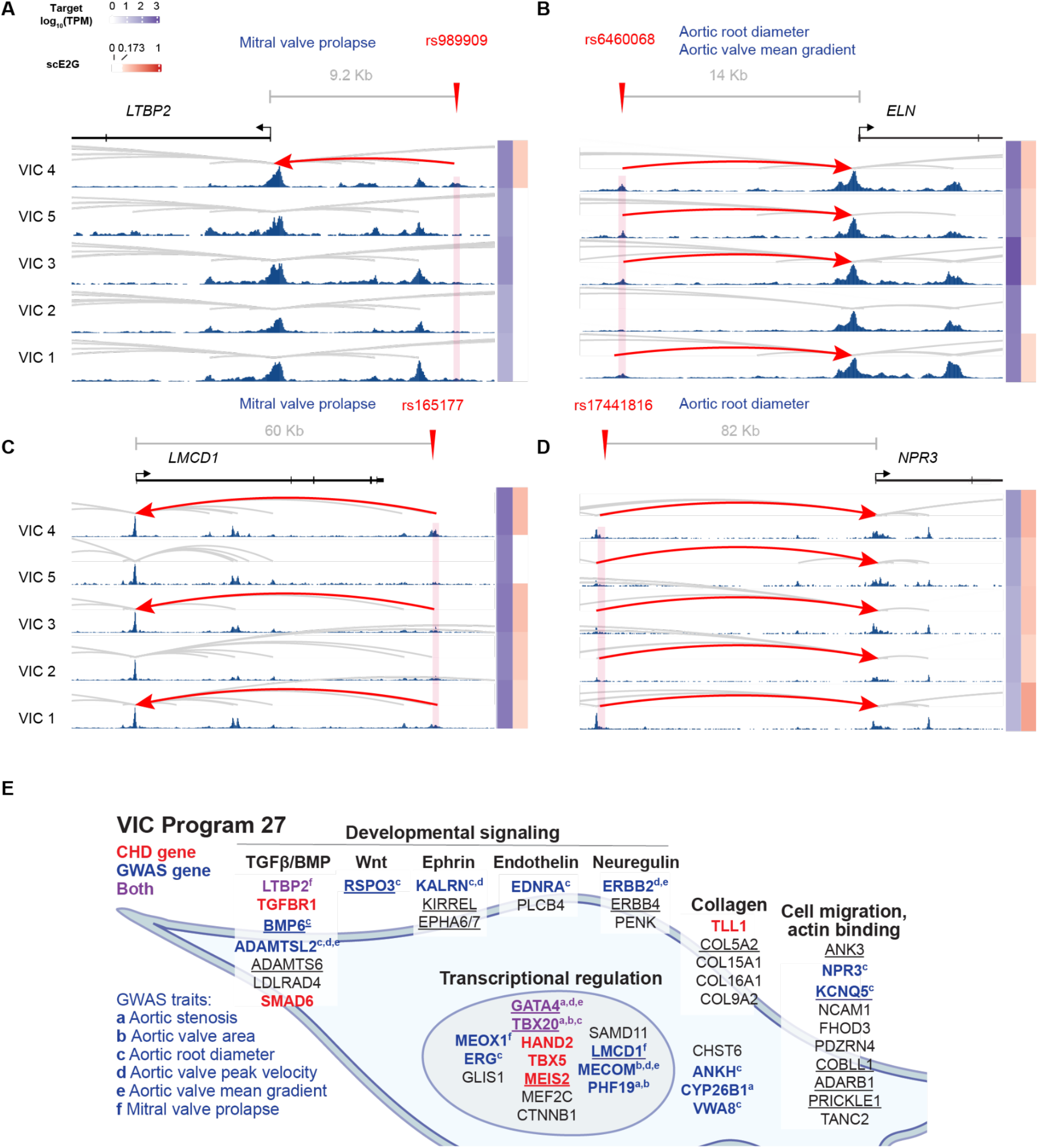
Variants for CHD and valve traits link to shared genes and programs in VICs. **A,B,C,D.** scE2G predictions and normalized scATAC-seq signals are shown for *LTBP2* (A), *ELN* (B), *LMCD1* (C), and *NPR3* (D) showing that GWAS variants associated with valve diseases or traits (red downward arrows) overlap enhancers linked to these genes in VICs. Enhancer predictions overlapping variants are highlighted in red. Arcs represent the predicted enhancer-gene regulatory interactions. Side color bars show the TPM expression of the target genes in each cell type (blue) and the corresponding scE2G scores (red; white for scores below the threshold). The hg38 coordinates of the highlighted enhancer regions are as follows: LTBP2 chr14:74621456-74621956; *ELN* chr7: 74013872-74014372; *LMCD1*: chr3:8560989-8562461; *NPR3*: chr5: 32629183-32629683. **E.** Annotation of genes in Fetal VIC program 27, which contains many CHD genes (red), genes linked to noncoding GWAS variants for valve or aortic traits (blue), or genes with both types of evidence (purple). Black genes represent other genes in Fetal VIC program 27 with related functions. Underlined genes also appear in hiPSC-VIC programs 9 and/or 16 (see Fig. 4).

We first assessed the overall enrichment of noncoding GWAS variants in VIC enhancers for six diseases and quantitative traits related to the valves and the aorta, including our recently conducted GWAS for measurements of aortic valve hemodynamics^88,89^ (mitral valve prolapse^87^, aortic stenosis^90^, aortic valve area (AVA)^86^, aortic root diameter^91^, and peak velocity and mean gradient across the aortic valve^88,90^, see **Methods**; 560 total GWAS signals, **Table S16**). Using stratified linkage disequilibrium score regression (S-LDSC), we observed strong enrichment for genome-wide heritability in fetal VIC enhancers (up to 14-fold enrichment, for mitral valve prolapse in predicted enhancers in VIC_4, a population of VICs expressing the markers of neural crest-derived mesenchymal cells; **Fig. S19; Table S15)**.

Sixteen GWAS signals for valve traits contain noncoding variants linked to eight high-confidence CHD genes (*ELN, FBN1, GATA4, JAG1, LTBP2, NKX2-5, PIGL, TBX20*) by VIC enhancers (**Table S16)**. For example, variant rs989909 is associated with mitral valve prolapse and overlaps an enhancer predicted to regulate *LTBP2* in VIC_4 (**Fig. 3A**). Variant rs6460068 is associated with aortic root diameter and aortic valve mean gradient and overlaps a VIC enhancer predicted to regulate *ELN*, a ubiquitous extracellular matrix protein in which loss-of-function mutations lead to supravalvular aortic stenosis and valve defects^92,93^ (**Fig. 3B**). These observations suggest that common noncoding variants may tune the expression of CHD genes in fetal VICs to impact valve structure.

Many other GWAS signals for valve traits were linked to genes that are part of the same pathways as CHD genes. For example, we found an enrichment of predicted target genes of valve trait GWAS signals in Fetal VIC program 27 (“VIC migration and TGF-β signaling”) — the same program noted above that contains *LTBP2*, *GATA4*, and seven other CHD genes (**Fig. 3E**). A total of 19 genes linked to valve traits via GWAS variants in VIC enhancers are members of Fetal VIC program 27 (5.4-fold enrichment, FDR = 5.7 x 10^-6^). For example, a noncoding variant associated with mitral valve prolapse (rs165177) is predicted to regulate *LMCD1*, which encodes a TF that inhibits the activity of GATA6 and whose knockdown in zebrafish leads to AV valve regurgitation^94^ (**Fig. 3C**). Another variant associated with aortic root diameter, rs17441816, is predicted to regulate *NPR3*, a gene where genetic misregulation has been observed in the context of valve insufficiency^95^ and progressive aortic root dilatation in humans^96^ (**Fig. 3D**). Using a transgenic reporter assay, we confirmed that this *NPR3-*linked variant reduces enhancer activity in the mouse conotruncal region (**Fig. S23**).

Together, these observations suggest that noncoding variants affect fetal VIC enhancers to regulate valve structure and function. The target genes include known CHD genes, where loss-of-function mutations lead to more severe heart defects, and other genes that are co-regulated in the same gene pathways. Thus, rare and common variants associated with valve diseases involve shared genes and pathways in VICs. Notably, this suggests that polygenic regulation of valve structure might also influence risk for CHD (see **Discussion**).

### CHD genes alter developmental expression programs in VICs

The strong enrichment of high-confidence CHD genes in VICs, along with their enrichment in certain gene programs, suggests that some may act in shared regulatory networks to alter developmental programs in VICs. To experimentally test this hypothesis, we used CRISPRi to knock down genes in VICs differentiated from human induced pluripotent stem cells (hiPSC-VICs)^97^ (**Fig. 4A, Fig. S20**). The hiPSC-VICs expressed canonical VIC identity markers such as *COL1A1* and *COL3A1*, as well as markers used to characterize fetal VICs in our atlas (such as *VIM, ACTA2,* and *LEF1*) (**Fig. 4B**).

**Figure 4:**
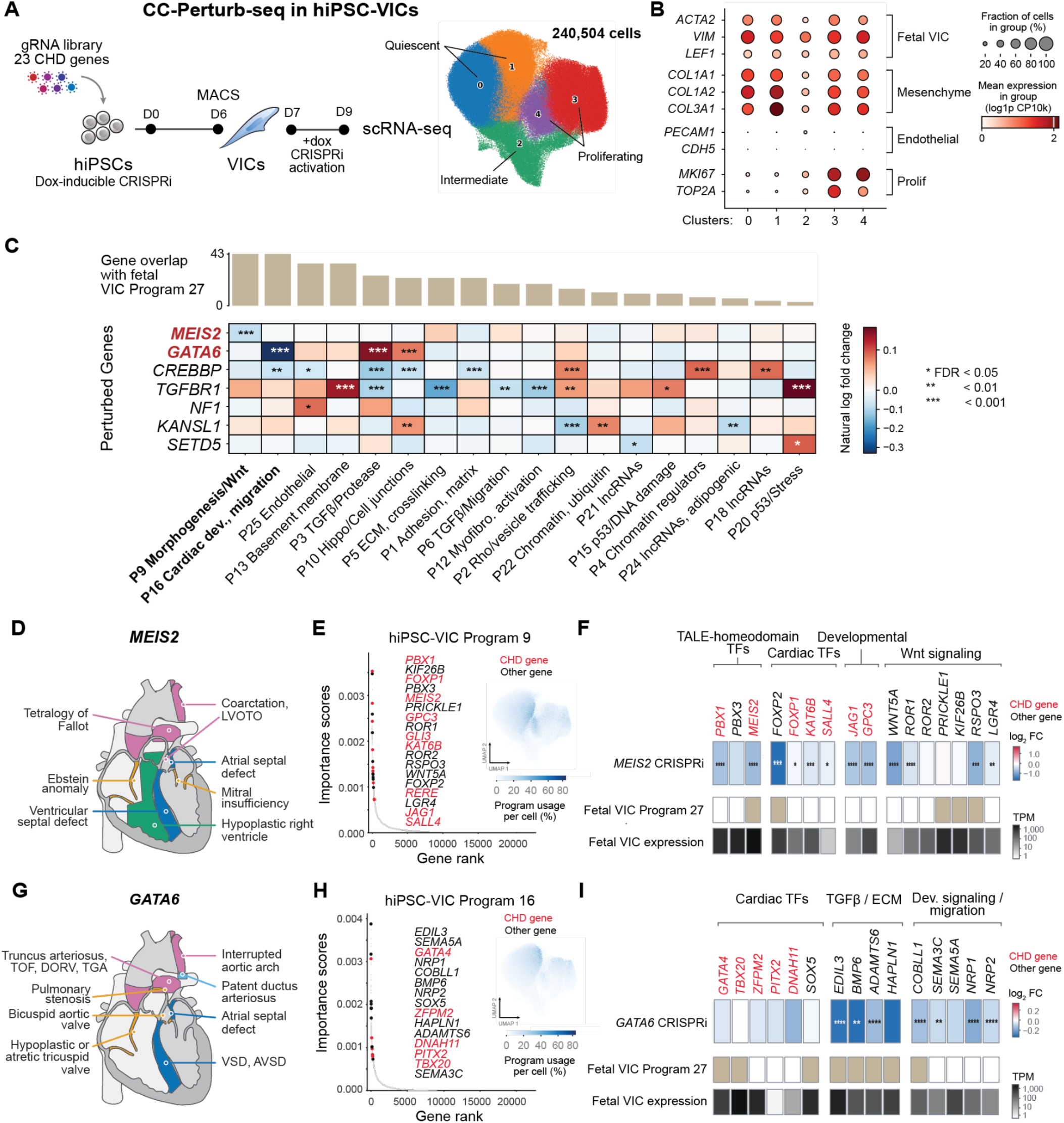
CHD genes affect gene regulation in hiPSC-VICs. **A.** CC-Perturb-seq was used to knockdown 23 CHD risk genes known to be enriched in fetal VICs. hiPSCs were infected with a lentiviral sgRNA library and differentiated into VICs and VECs (valvular endocardial cells) via cardiac lateral plate mesoderm (hiCLPM). VICs were separated through MACS (magnetic cell separation) and treated with doxycycline to induce CRISPRi expression. **B.** UMAP embedding of the guide-infected hiPSC-VICs shows canonical VIC marker expression (*COL1A1*, *COL1A2*, *COL3A1*, *VIM*, *ACTA2*, *LEF1*) and low endothelial marker expression. **C.** Propensity-score matching of perturbed to unperturbed cells per gene target to identify significant effects of perturbations on gene program expression, with effect on program usage plotted. Significance is BH-corrected (FDR * < 0.05, **< 0.01, ***< 0.001). Bars show program loading overlap (top 300 genes) with the Fetal VIC Program 27. **D.** Schematic showing CHD defects linked to *MEIS2* mutations. These include left ventricular outflow tract obstruction (LVOTO), coarctation of the aorta, Tetralogy of Fallot, Ebstein anomaly, ASD, mitral insufficiency, VSD, and hypoplastic right ventricle.^109^ **E.** Ranked importance scores of genes associated with hiPSC-VIC program 9; The top 300 genes by importance scores in hiPSC-VIC program 9 were considered as program genes. Black: Relevant pathway genes in the top 300 genes; Red: high-confidence CHD genes from CHDgene in the top 300 genes^9^. Inset: Projection of hiPSC-VIC program 9 expression on the hiPSC-VIC UMAP, showing overall expression. Color scale: gene program usage per cell. **F.** Knockdown of *MEIS2* leads to differentially regulated genes in TALE-homeodomain signaling, cardiac TFs, developmental signaling, and Wnt signaling pathways. Differential gene expression (log2fc) from bulk RNA sequencing data is calculated through DESeq2 with significance determined through Wald’s test and BH correction with 4 biological replicates per condition (*MEIS2* sgRNA vs NT control). Membership of each gene in Fetal VIC Program 27 is denoted in tan. Averaged pseudo-bulk expression of each gene in the *in vivo* fetal VIC populations is shown in mean TPM. **G.** Schematic showing CHD defects linked to *GATA6* mutations. These include interruptions in aortic arches, patent ductus arteriosus, ASD, VSD, AVSD, hypoplastic/atretic tricuspid valve, bicuspid aortic valve, pulmonary stenosis, truncus arteriosus, tetralogy of fallot, double outlet right ventricle, and transposition of the great arteries.^110^ **H.** Ranked importance scores of genes associated with hiPSC-VIC program 16; The top 300 genes by importance scores in hiPSC-VIC program 16 were considered as program genes. Black: Relevant pathway genes in the top 300 genes; Red: high-confidence CHD genes from CHDgene in the top 300 genes^9^. Inset: Projection of hiPSC-VIC program 16 expression on the hiPSC-VIC UMAP, showing overall expression. Color scale: gene program usage per cell. **I.** Knockdown of *GATA6* leads to differentially regulated genes in TGFβ/ECM, cardiac TFs, and developmental signaling/migration pathways. Differential gene expression is calculated through SCEPTRE^76^ analysis in CC-Perturb-seq, where conditional resampling is used to determine log_2_ fold change of the response gene from null, with BH correction for statistical significance. Membership of each gene in Fetal VIC Program 27 is denoted in tan. Averaged pseudo-bulk expression of each gene in the *in vivo* fetal VIC populations is shown in mean TPM.

We targeted each of the 23 CHD genes highly expressed in fetal VICs (**Fig. 2E**), measured effects on full transcriptome gene expression in a pooled CRISPRi experiment (Circular capture Perturb-seq (CC-Perturb-seq)^98^, see **Methods**), and used cNMF to learn gene programs (hereafter “hiPSC-VIC programs;” following^65^). Seven of the 19 successfully perturbed CHD genes significantly affected at least one of the learned programs: *CREBBP, GATA6, KANSL1, MEIS2, NF1, SETD5,* and *TGFBR1* (**Figs. 2E, 4C**). Four of 23 genes were not informative in this experiment, either due to low knockdown (*NOTCH2, ARID1B*) or because Perturb-seq cannot identify cell non-autonomous effects, *e.g.* of secreted factors (*LTBP2, FBN1*).

We found that these CHD gene knockdowns often regulated other CHD genes and genes in the *in vivo* Fetal VIC program 27, as well as effects on genes known to be involved in developmental signaling and cell migration. For example, *MEIS2* mutations are observed in human patients showing a wide range of CHD including septal defects, valve defects, and malformations of the outflow tract (**Fig. 4D**, **Table S11**). *MEIS2* knockdown significantly affected hiPSC-VIC program 9, which included 8 other CHD genes (**Fig. 4E**) and other genes previously implicated in aspects of heart or valve development such as: cardiac TFs (*FOXP2, FOXP1, KAT6B,* and *SALL4)*, developmental signaling (*JAG1* and *GPC3)*, and WNT signaling (*WNT5A, ROR1, ROR2, PRICKLE1, KIF26B, RSPO3*, and *LGR4*) (**Fig. 4F)**. Many genes in hiPSC-VIC program 9 were shared with Fetal VIC program 27 and/or were highly expressed in fetal VICs. These results implicate *MEIS2* as important for valve development and specification through cell-autonomous effects in VICs.

*GATA6* mutations lead to a wide array of cardiac defects (**Fig. 4G**). *GATA6* knockdown significantly affected hiPSC-VIC program 16, which contained 5 CHD genes, including cardiac TFs (*GATA4, TBX20, ZFPM2, PITX2, DNAH11, SOX5*) and genes involved in TGF-β signaling and extracellular matrix maintenance^99^ (*EDIL3, BMP6, ADAMTS6, HAPLN1*), and cell migration^100^ (*COBLL1*^101^*, SEMA3C*^102,103^*, SEMA5A*^104^*, NRP1*^102^*, NRP2*^105^) (**Fig. 4H, I)**. These observations in hiPSC-derived human VICs are consistent with previous observations in mouse; conditional knockout of *Gata6* in the neural crest lineage led to disrupted *Sema3c* expression, impaired cardiac OFT patterning, and abnormal septation^103–105^. *Sema3c* secreted from neural crest has been observed to bind to *Nrp1* in the endothelium, leading to EndMT and OFT septation^102^. Notably, these genes are expressed in all five VIC subpopulations we annotated in the fetal heart, which likely include cells derived from both the neural crest^106^ and mesoderm^107,108^. Thus, *GATA6* could lead to CHD in humans by cell-autonomous regulation of key patterning and cell migration genes in VICs.

Together, these perturbation experiments show that many of the CHD genes expressed in fetal VICs have cell-autonomous effects in hiPSC-VICs, regulating the expression of other genes with known roles in developmental signaling, cell migration, and CHD. These results indicate that genetic risk for CHD may often involve cell-autonomous dysfunction of VICs (see **Discussion**).

### A noncoding CHD variant affects the activity of a fetal endocardial enhancer

In addition to the many CHD genes discovered through rare coding variation, a small handful of noncoding variants have been associated with subtypes of CHD through GWAS^13–16^. We applied our gene regulation map to discover the enhancers, cell types, and TFs that mediate the effects of variants associated with CHD, as well as with 41 other structural heart diseases and traits^78,86,111–123^ (**Fig. S22; Tables S14, S16**).

In total, our regulatory map linked noncoding variants for 750 unique GWAS signals to enhancers and target genes (**Table S16**). Heritability for each trait was enriched in a distinct set of cell types (**Fig. S22B; Table S15)**. At individual loci, many of the prioritized enhancers were predicted to be active in specific cell types and spatial locations, providing new insights into the genetic etiology of heart structure and function (**Fig. S22, Note S4**).

While the small number of GWAS signals for CHD (18 loci across nine studies) does not yet allow robust enrichment analysis, we identified 11 cases where noncoding variants associated with CHD overlapped enhancers in our fetal heart atlas, including variants linked to *WNT9B/GOSR2*, *PDGFB*, and *MYL4* (**Table S16**).

The single-nucleotide variant rs17608766 at the *WNT9B/GOSR2* locus stood out as a strong candidate for functional testing. This common variant has been associated with malformations of the great arteries, ASD, aortic valve size, and other quantitative traits (**Fig. 5A**). In earlier work, we linked it to *GOSR2* and *WNT9B* using enhancer maps and QTL datasets from adult tissues and cell lines^35,37^. However, its activity has not been experimentally tested or annotated in the fetal heart.

**Figure 5:**
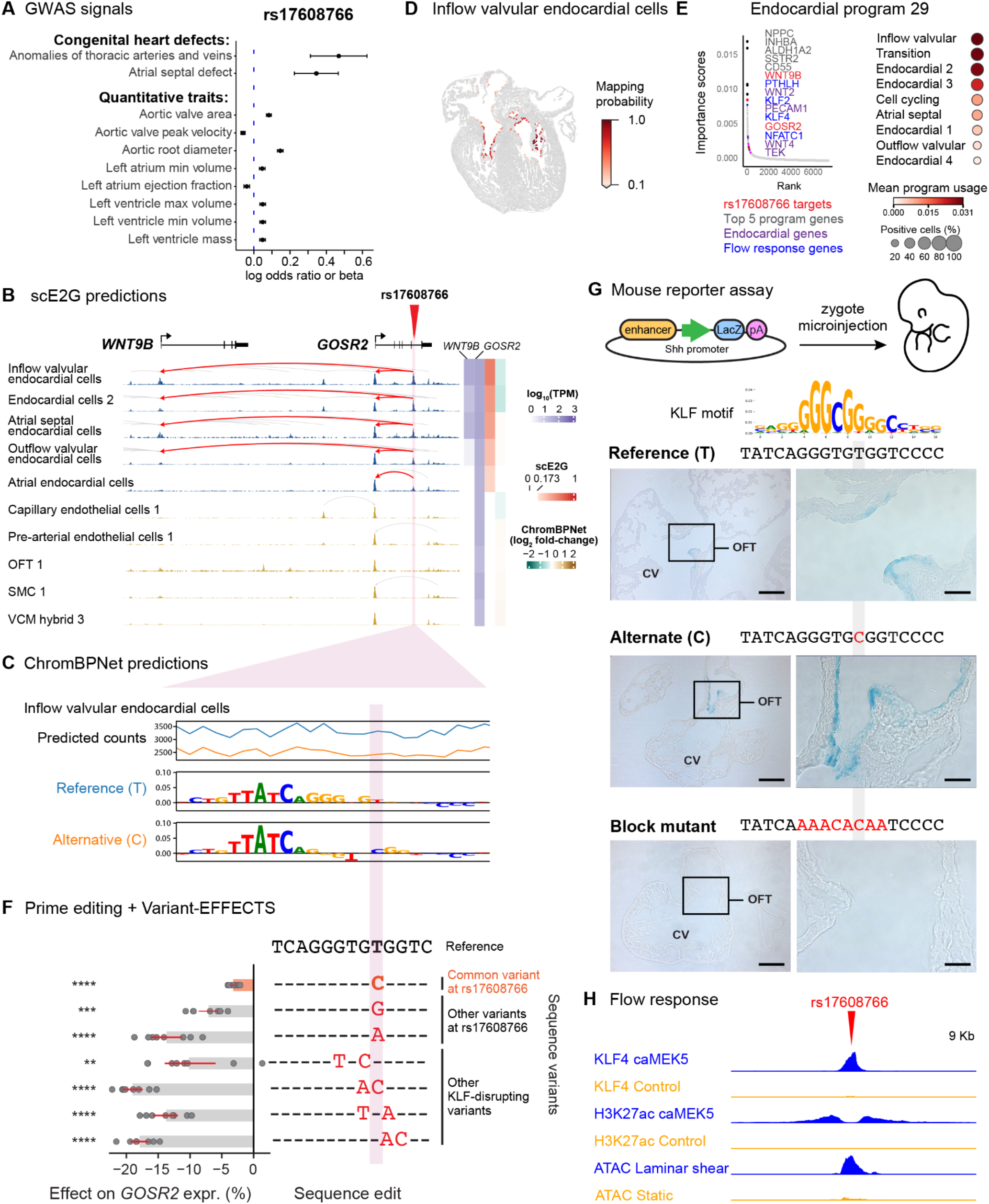
rs17608766 affects the activity of an endocardial cell enhancer activated by LSS. **A.** Associations of rs17608766 with risk for subtypes of CHD and quantitative measurements of the aortic valve and heart chamber traits. X-axis: the log odds ratio (for binary traits) or the beta per-unit change (for quantitative traits) associated with the alternative allele. Error bars represent 95% confidence intervals. See Supplementary Table S14 for data sources. **B.** rs17608766 overlaps an enhancer (hg38: chr17:46935201-46936149) predicted to regulate *GOSR2* and *WNT9B* in endocardial cells (red highlight and arcs). Signal tracks represent chromatin accessibility from scATAC-seq. Gray arcs represent other predicted enhancer-gene regulatory interactions in the locus. The side color bars show the TPM expression of *WNT9B* and *GOSR2* in each cell type (blue), scE2G scores (red, and white for scores below the threshold), and ChromBPNet predicted log_2_ fold changes in chromatin accessibility upon substitution of the reference allele (T) with the alternative allele (C) (green indicates reduced accessibility, brown indicates increased accessibility). IF: inflow. **C.** ChromBPNet predictions for rs17608766 in inflow valvular endocardial cells. The alternative allele is predicted to lead to a decrease in chromatin accessibility across the element (top). Middle and bottom tracks show contribution scores for the reference and alternative alleles. **D.** Spatial mapping of IF valvular endocardial cells (spatial data from ^48^). Color intensity indicates the mapping probability that each cell from the spatial data corresponds to the cell cluster defined in this study. **E.** (left) *WNT9B* and *GOSR2* are among the genes in Endocardial Program 29, along with markers for endocardial cells and flow response. (right) Expression of Endocardial Program 29 across endocardial cell populations. **F.** Variant-EFFECTS measurements of effects of variants on *GOSR2* expression in human aortic endothelial cells. Bars: mean (gray) ± 95% CI (red). Dots: n = 8 FlowFISH replicates. Asterisks (q) indicate Benjamini-Hochberg corrected p-values from one-sample, two-tailed t-tests (q < 0.01**, 0.001***, 0.0001****). The effect of the rs17608766 risk variant is highlighted in orange. **G.** Mouse transgenic assay: Three versions of the predicted enhancer containing rs17608766 (reference sequence, T>C variant, and block mutant) were cloned to drive the expression of *LacZ* and assayed in a binary fashion (on vs. off). Representative brightfield images show presence or absence of *LacZ* reporter expression (blue) in mouse embryonic heart sections at E11.5. Left: Outflow tract (OFT) and common ventricle (CV). Right: Zoom-in shows that *LacZ* expression is restricted to the surface of the developing valves (endocardial cells). Scale bars: left column = 200μm; right column = 50μm. See **Fig. S23** for biological replicate embryos. **H.** The enhancer overlapping rs17608766 responds to laminar flow *in vitro* in pulmonary arterial endothelial cells (PAECs; data from^124^). Enhancer shows higher binding for KLF4 and H3K27ac (ChIP-seq) in PAECs transduced with constitutively active MEK5 (mimicking LSS), and shows higher chromatin accessibility (scATAC-seq) in PAECs exposed to LSS compared to static conditions. Paired tracks share scales.

In our heart map, rs17608766 overlapped an enhancer predicted to regulate *GOSR2* and *WNT9B* in several endocardial cell subtypes (**Fig. 5B**). The alternative allele (C) was predicted to reduce chromatin accessibility of this enhancer in endocardial cells by altering a motif instance for a KLF family TF (**Fig. 5C**). The cell types predicted to be most strongly affected by the variant included a subpopulation of endocardial cells on the inflow side of the valves and a subpopulation of endocardial cells lining the inner surface of the cardiac chambers (**Fig. 5C**), whose locations we verified by mapping to spatial data (**Fig. 5D, Fig. S9**). Consistent with these observations, the target genes *WNT9B* and *GOSR2* are both members of Endocardial program 29 (“WNT signaling in valvular endocardial cells”), which is regulated by TFs induced by laminar shear stress (LSS) (KLF2/4) and is most active in IF valvular endocardial cells (**Fig. 5E, Fig. S21, Table S8**). In the developing heart, both genes plausibly affect CHD: *WNT9B* encodes a secreted signaling protein known to play a role in valve formation and cardiac cushion remodeling^69^, and *GOSR2* encodes a *cis-*Golgi SNAP receptor whose knockout in medaka leads to abnormal heart morphology^30^.

To validate that rs17608766 impacts gene expression, we applied CRISPR prime editing in a human aortic endothelial cell line (TeloHAEC, **Fig. 5F**). Endothelial cells are similar transcriptionally to endocardial cells, and the enhancer overlapping the variant was also predicted to be active in TeloHAEC^65^. We used a highly sensitive, pooled single-cell readout method (Variant-EFFECTS^64^) to measure effects of introducing the variant on the expression of *GOSR2*; *WNT9B* was not expressed in this cell model (**Table S22**). Introducing the risk allele at rs17608766 (T-to-C edit) with prime editing led to a slight but significant reduction in *GOSR2* expression (*P* = 7.6 x 10^-6^). Installation of multi-nucleotide variants that more strongly disrupted the predicted KLF motif instance led to stronger reductions in *GOSR2* expression (**Fig. 5F**). This demonstrates that the risk allele at rs17608766 indeed reduces the activity of an enhancer that regulates nearby gene expression in endothelial cells (**Table S23)**.

To validate the cell type specificity of the enhancer containing rs17608766, we tested its binary activity (on or off) in E11.5 mouse embryos using mouse transgenic reporter assay (**Fig. 5G**). Brightfield images confirmed that the enhancer is active in endocardial cells of the OFT of the developing mouse embryos.

Mutating the KLF motif instance in the enhancer abolished enhancer activity, but introducing the risk allele did not qualitatively affect enhancer activity (**Fig. 5G**). Thus, while the integrity of the KLF site is essential, the common CHD risk variant is likely to have a more subtle effect on this enhancer.

Given that KLF TFs are flow-responsive, we next tested whether the enhancer containing rs17608766 responds to LSS. Indeed, endothelial cells exposed to LSS or LSS-like conditions (constitutively active MEK5)^124^ showed increases in chromatin accessibility, H3K27ac occupancy, and KLF4 binding at the enhancer (**Fig. 5H**). Furthermore, among the 77 genes significantly differentially expressed between the two conditions, 31 genes, including *WNT9B*, were identified as part of Endocardial program 29, which showed the strongest enrichment across all programs (39-fold enrichment of differentially expressed vs all tested genes, FDR-adjusted *P*=2.4 x 10^-31^).

Together, our findings demonstrate that the enhancer identified by our gene regulatory maps is indeed active in the predicted cell types, and that the variant rs17608766 overlapping this enhancer directly regulates nearby gene expression. This strongly suggests that variation at rs17608766 alters the response of endocardial cells to LSS, a crucial force that shapes valve development, to affect risk for CHD.

## Discussion

Here, we illuminate the genetic and cellular etiology of CHD by building a comprehensive gene regulation map of the human fetal heart and applying it to systematically annotate coding and noncoding variants associated with CHD and cardiac traits—including variants associated with valve traits which we annotate here for the first time. Across 90 cell types in the human fetal heart, we implicate new cell populations in the genetic etiology of CHD subtypes, identify noncoding risk variants with cell type-and location-specific activity, and show that variants associated with CHD and quantitative valve traits converge on shared genes and programs. These findings provide a resource for interpreting variants observed in patient genomes, will help to guide future studies of CHD, and may point to new cellular or molecular targets for therapy.

By combining a comprehensive multiomic atlas of the fetal heart with known CHD genes and genetic variants, this study provides key evidence for the hypothesis that VICs and cardiac fibroblasts play direct roles in the genetic etiology and development of CHD^2,14,40,49,125^. In an unbiased analysis of 90 cell types in the human fetal heart, VICs and cardiac fibroblasts show the strongest enrichment for high-confidence CHD genes, including for multiple subtypes of CHD (**Fig. 2A**). While such enrichments may not be surprising for certain subtypes (*e.g.*, valve defects), the enrichments are equally as strong for others such as single ventricle defects, where a primary role for mesenchymal cells is less clear^126^. Consistent with this finding, previous studies have studied individual genes with effects on mesenchymal cells and heart development in animal models (**Note S2**), noted the expression of candidate CHD genes in mesenchymal cells in other datasets^2,14,40,49^, and observed differences in cardiac fibroblasts in the hearts of CHD patients after birth^127^. Our data expand on these observations by prioritizing specific genes and connecting them into molecular pathways in these cell types (**Fig. 2**), identifying noncoding variants associated with valve traits that link to known CHD genes in VICs (**Fig. 3**), and demonstrating that multiple of the prioritized CHD genes affect gene expression programs (GEP) associated with cell migration and developmental patterning in hiPSC-derived VICs (**Fig. 4**). Together, our results suggest that a large fraction of CHD genes, including genes such as *GATA6* and *MEIS2*, may lead to heart defects through primary effects in mesenchymal cells, which could include VICs and their mesenchymal progenitors. This has implications for our fundamental understanding of how heart defects arise beyond disease of the working myocardium and motivates further development of appropriate cell and animal models to study the contributions of these cell types to CHD.

We find that rare variants associated with CHD and common variants associated with quantitative measurements of valve structure and function affect many of the same genes and programs in VICs (**Fig. 3E**). Knockdown of *GATA6* and *MEIS2* affect the expression of multiple other CHD genes in this program (**Fig. 4E, F, H, I**). This identifies a specific pathway that could mediate polygenic contribution to CHD. Indeed, recent studies have suggested the possibility of polygenic or oligogenic architectures for CHD in which different heart defects might arise from combinations of causal genetic variants in each individual^13–16,34,128,129^. Patients or family members with rare variants in the same genes are often observed to have different types of CHD, and combining causal variants in animal models has directly confirmed certain genetic interactions^128^. Previous studies have noted individual cases where GWAS loci for quantitative cardiac traits are located near CHD genes^37^, and observed genome-wide genetic correlations between certain quantitative traits and CHD risk, such as between interventricular septum structure and risk for VSDs^34^. However, specific molecular pathways that mediate such effects have not previously been identified. Our data suggest a possible model in which common, inherited genetic variation tunes the structure of the valve by influencing VICs during fetal development. In this model, the severity or form of CHD would be influenced by the combination of one or more rare, high-effect variants (*e.g.*, in genes such as *GATA6* or *MEIS2*) and common, mostly noncoding variants that regulate some of these same genes or other genes in the same molecular pathways (**Fig. 3E**).

Our atlas of noncoding regulatory elements enabled the first systematic assessment of the roles of inherited noncoding variants associated with CHD and quantitative cardiac traits during fetal development. We link hundreds of such noncoding variants to candidate enhancers, cell types, genes, and pathways (**Table S16**). Individual variants are predicted to act in specific cell types (*e.g.*, VICs and valvular endocardial cells for BAV), but collectively span dozens of different cell types, highlighting the importance of considering the diverse totality of cell types present during cardiogenesis. Functional testing of four of these enhancers revealed unique, spatially constrained activity patterns, suggesting that specific spatial patterns of enhancer activity could explain how some noncoding variants affect specific aspects of heart structure (**Fig. 5, Fig. S23**). These specific patterns also highlight the utility of our cell type-specific enhancer catalog for identifying and testing enhancers with spatial specificity, which may be leveraged to develop expression vectors for gene therapies that target specific cell types, locations, or developmental timepoints.

Our study illustrates a strategy for building gene regulation maps to interpret the impact of human genomic variation on function. In particular, by combining a high-depth single-cell multiomic atlas with predictive models validated to predict effects of perturbations, we can construct regulatory maps of noncoding variants (with ChromBPNet) and enhancers (with scE2G) in cell types that are sequenced to a depth of at least 3 million scATAC-seq fragments. Here, this strategy allows us to annotate impacts on chromatin accessibility and target genes across 90 cell types and cell states in the fetal heart. This increased depth proved critical in identifying the cell type of action for variants that affected rare or spatially specific cell types such as VICs and endocardial cells (**Figs. 3, 5**), which were not resolved in previous atlases^4,45,60^.

However, deeper data will be required to identify rarer cell types, or cell states that occur only at certain time points during development. For example, we observe rare subpopulations of migratory epicardial cells (**Fig. S5**) where too few cells were captured even in this dataset to build accurate maps of gene regulatory elements. Our atlas also lacks hearts earlier than PCW 6, timepoints that are critical to the formation of the heart and that may include additional cell types and states important for the genetic etiology of CHD. As such, the variants and genes we describe here may have functions in other cell types or stages not included in our atlas. Expanding single-cell multiomic studies to include earlier time points would provide a more comprehensive understanding of how our genome encodes heart development, enabling additional discoveries that were previously only possible in model systems.

While our trajectory analysis reveals clear maturation dynamics within the VIC population, these findings must be interpreted conservatively. Asynchronous cellular maturation means that early and mature cell states can coexist within a timepoint, making chronological age insufficient to describe differentiation paths. Furthermore, cell state proportions differ across samples due to the anatomical sampling strategy. Fully mapping these temporal dynamics remains an important future direction that will likely require deep spatially resolved datasets.

Together, this work provides new insights into the genetic etiology of CHD, and a foundational map for interpreting effects of genetic variants on heart structure and function. As genome sequencing and association studies of CHD continue to expand, we anticipate that this map will enable many additional discoveries about the genes, pathways, and cell types that contribute to heart development and disease.

## Methods

### Human Heart Tissue Sample Collection

Human fetal heart tissue samples were collected from three primary sources: Stanford University, Cercle Allocation Services, Inc., and the University of Washington (UW) Birth Defects Research Laboratory (BDRL). Each institution obtained ethical approval from their respective ethics committees and ensured compliance with all applicable ethical and legal guidelines, including obtaining written donor consent. Stanford University: Approval was granted by the Stanford Institutional Review Board (IRB 25871), the Stem Cell Research Oversight Committee (SCRO 791), and the Administrative Panel on Biosafety (APB-4781-JE0423). Cercle Allocation Services, Inc.: Approval was obtained from a WCG Clinical Services Institutional Review Board (IRB 20111833). University of Washington (UW) Birth Defects Research Laboratory (BDRL): Approval was obtained from the UW Institutional Review Board (CR ID CR00003603, Study ID STUDY00000380). All fetal tissue samples were obtained from elective terminations in accordance with institutional protocols and donor consent procedures. The collection, storage, and use of these samples were conducted in compliance with ethical standards and regulatory requirements governing research involving human tissues.

All collected tissues were screened and confirmed to be negative for trisomies and other non-single ventricle defects based on prenatal screening. The PCW of the tissue was determined through ultrasound assessment and by referencing the date of the last menstrual period prior to pregnancy termination. A total of 41 structurally normal human fetal hearts, ranging from 6 to 22 PCW, were collected from these sources. Samples from Cercle and BDRL were stored in 1X HBSS+/+ (Gibco 14025-092) before being flash-frozen and shipped overnight on dry ice for cryobanking at −80°C. Samples collected directly from Stanford University were flash-frozen in-house and cryobanked at −80°C. Most samples were flash-frozen within 2 hours of the procedure.

### Heart Sample Preparation for Single-Cell Multiome

Three protocols were developed to process heart tissue samples, each tailored to meet specific sample conditions and experimental needs. Protocol 1 (http://dx.doi.org/10.17504/protocols.io.rm7vzjez5lx1/v1) is designed for fresh tissue, providing high-quality nuclei for downstream analyses due to its ability to preserve sample integrity. Protocol 2 (http://dx.doi.org/10.17504/protocols.io.rm7vzjez5lx1/v2) is optimized for handling small or limited tissue samples and is preferred when nuclei retention is critical, ensuring that even minimal sample inputs yield viable results. Protocol 3 (http://dx.doi.org/10.17504/protocols.io.rm7vzjez5lx1/v3), which was used for processing the majority of the samples in this study, provides consistency and scalability across larger sample sets due to its ability to maintain good quality nuclei and effective purification steps. The methods described within this subsection are based on Protocol 3, and all three protocols can be accessed via Protocols.io for detailed step-by-step guidance. Details on which samples were processed using Protocols 1, 2, or 3 can be found in **Table S2**.

Human fetal heart samples from 41 cases, ranging from 6 to 22 PCW, were processed (see **Table S1** for sample attribute information and **Table S2** for sample metadata details). These samples were flash-frozen before being processed for the experimental pipeline. Each sample was mechanically homogenized separately in a 15 mL pre-chilled Dounce tissue grinder (Millipore Sigma D9938) with 4 mL of 1x Homogenization Buffer, which consisted of 260 mM sucrose, 30 mM KCl, 10 mM MgCl2, 20 mM Tricine-KOH pH 7.8, 1 mM dithiothreitol (DTT), 0.5 mM spermidine, 0.15 mM spermine, 0.3% NP40, 60 U/mL Ribolock RNase Inhibitor, 1% BSA, and 0.5–1 tablet of cOmplete Protease Inhibitor. To begin, 20– 100 mg of flash-frozen tissue was cut with a razor blade in a tissue culture dish placed on an ice block over dry ice to prevent premature thawing. The tissue was then transferred to the Dounce tissue grinder. The tissue was allowed to thaw for 5 minutes in the chilled buffer before homogenization. Homogenization was performed using 10–20 strokes with pestle A (loose), followed by 10–20 strokes with pestle B (tight), depending on the tissue amount and developmental stage (older tissue required more strokes). The homogenized solution was then filtered through a 100-µm cell strainer (PluriSelect 43-50100-51) into a pre-chilled 50 mL conical tube. To rescue any remaining nuclei, an additional 400 µL of 1x Homogenization Buffer was used to rinse the Dounce tissue grinder and passed through the same 100-µm cell strainer. The sample was further filtered by gently pipetting in 400–500 µL increments through a 70-µm Flowmi cell strainer (Bel Art H136800070) into a new 5 mL Eppendorf low-binding tube. Nuclei were pelleted by centrifugation at 350g for 5 minutes at 4°C, after which all but 50 µL of supernatant above the pellet was removed. The nuclei were then resuspended in 750 µL of 1x Homogenization Buffer, bringing the total volume to 800 µL. Further purification was achieved using an Iodixanol density gradient. Each sample was mixed with an equal volume (800 µL) of 50% Iodixanol Solution (60% Iodixanol, 150 mM KCl, 30 mM MgCl2, 120 mM Tricine-KOH pH 7.8, nuclease-free water), resulting in a 25% nuclei mixture. The mixture was gently pipetted to ensure thorough mixing. A layer of 1000 µL of 30% Iodixanol Solution (1x Homogenization Buffer, 50% Iodixanol) was carefully added beneath the 25% nuclei mixture, followed by 1000 µL of 40% Iodixanol Solution to create a third layer. To prevent mixing between layers, the sides of the pipette tip were wiped with a kimwipe, and the tube was kept upright throughout the process. The sample was centrifuged at 3500g for 15 minutes at 4°C with the brake off (deceleration 0, acceleration 5). This process resulted in three distinct layers: the top layer (debris), the middle nuclei layer (target layer), and the bottom layer (more debris). The top layer was aspirated down to within 200– 300 µL of the nuclei band. A maximum of 200 µL was collected from the target layer and transferred to a fresh 1.5 mL Eppendorf low-binding tube, avoiding excess debris. The nuclei were washed in 1 mL of 1x Homogenization Buffer, topped off to 1.5 mL, and centrifuged at 1200g for 5 minutes at 4°C. The supernatant was removed, and 700 µL of ATAC-RSB-Tween buffer (10 mM Tris-HCl pH 7.5, 10 mM NaCl, 3 mM MgCl2, nuclease-free water, 0.1% Tween-20, 1% BSA, 1 U/µL RNase Inhibitor) was added to resuspend the nuclei pellet. The solution was gently mixed by pipetting and filtered through a 40-µm Flowmi cell strainer (Bel Art H136800040) into a new 1.5 mL Eppendorf low-binding tube. Nuclei quality was assessed using Trypan Blue staining and manual counting with a hemocytometer (Hausser Scientific 1490)_50,130_.

### Single-Cell Multiome Library Preparation and Sequencing for Heart Tissue

Nuclei isolated from the previous section were adjusted to a concentration of 10,000 nuclei/µL and loaded onto the 10x Genomics Chromium Controller. Nuclei were prepared for single-cell multiome sequencing following the Chromium Single Cell Multiome ATAC + Gene Expression kit (10x Genomics, CG000338 Rev F) with the Chromium Single Cell Multiome ATAC + Gene Expression Reagent Bundle (PN-1000283) and the Chromium Next GEM Chip J Single Cell Kit (PN-1000234). Libraries were uniquely indexed with Single Index Kit N Set A (PN-1000212) for snATAC-seq and Dual Index Kit TT Set A (PN-1000215) for snRNA-seq. Quality control for the cDNA and final libraries was performed using the Bioanalyzer High Sensitivity DNA Analysis Kit (Agilent, 5067-4626) and the Qubit 1X dsDNA High Sensitivity Assay Kit (Thermo Fisher Scientific, Q33231). Sequencing libraries were pooled and sequenced across multiple Illumina instruments — NextSeq 550, NovaSeq, NovaSeq X, and NovaSeq X Plus — to accommodate different sample batches, achieving a mean read depth of at least 20,000 total reads per nucleus for RNA and 25,000 total reads per nucleus for ATAC.

### scRNA-seq alignment and background removal

The raw sequencing data was converted to FASTQ format using bcl2fastq. ScRNA-seq reads were aligned to GENCODE v29 (https://woldlab.caltech.edu/~diane/genome/GRCh38-V29-male-2.7.8a.tar.gz) using STARSolo^131^ (v2.7.11a) following the parameters specified in starsolo.snakefile (https://github.com/detrout/woldlab-rna-seq/tree/main, commit f592629). For the downstream analysis, we used the “GeneFull_Ex50pAS” count matrix, which contains all reads overlapping genes’ exons and introns. We used CellBender^132^ (v0.3.0) to remove ambient RNA from the count matrices with a false positive rate (0.01). The “expected-cells” and “total-droplets-included” parameters were set based on knee plots of UMI counts per cell barcode, as recommended by the CellBender user manual. We set “--learning-rate” and “--epochs” to 0.00002 and 150, respectively. CellBender’s evaluation was used to determine if reruns with different parameters were necessary. In such cases, the learning rate was halved and epochs were increased to a maximum of 300 until the performance was evaluated to be satisfying.

### scATAC-seq alignment

The raw sequencing data were converted to FASTQ format using bcl2fastq. Barcode matching was performed using matcha^133^ (v0.0.2) with the maximum hamming distance to tolerate between overserved and whitelisted barcodes set to 1. The scATAC-seq reads were aligned to the GRCh38 reference genome (ENCODE: GRCh38_no_alt_analysis_set_GCA_000001405.15) using bowtie2^134^ (v2.4.4). The aligned reads were filtered using samtools^135^ (v1.13) view with the parameters “-F 524 -f 2” to only keep properly paired mapped reads that passed platform/vendor quality checks. Read pairs mapped to more than 4 locations were discarded. Duplicate reads were removed using Picard’s MarkDuplicates^136^ (picard-slim v2.35.7) with lenient validation stringency. We further filtered for primary alignments and discarded reads mapped to the mitochondrial genome. Finally, the alignment was sorted by coordinates and was converted to the fragment file format, during which the reads were shifted + 4 bp and − 4 bp for positive and negative strand respectively to account for Tn5 transposase insertion during library preparation.

### scATAC-seq quality control

We calculated the nucleosome signal for each cell by computing the ratio of mononucleosomal (147–294 bp) to nucleosome-free (<147 bp) fragments sequenced for each cell. Only cells called by STARSolo (see ScRNA-seq alignment and background removal) and with nucleosome signals lower than 2 were considered for further analysis. These cells were read into the ArchR^137^ (v1.0.2) and filtered based on two criteria: a minimum transcription start site (TSS) enrichment score of 6 (“minTSS=6”) and at least 1000 mapped scATAC-seq fragments per cell (“minFrags=1000”). We then reviewed the log-transformed unique nuclear fragments versus TSS enrichment score plots of each library. For libraries with higher sequencing depth or distinct clusters of cells with low fragment counts, we increased the minimum fragment threshold to either 2000 or 3000 fragments to exclude low-quality cells.

### scRNA-seq quality control and normalization

After calling cells using STARSolo and removing cells that did not pass the ATAC quality control (see **scATAC-seq quality control)**, to account for variations in sequencing depth and library quality, we performed library quality control for each library. We examined the distribution of three key metrics:

1. Number of UMIs per cell: we filtered out cells with very low (cutoffs ranged from 200 to 2000 UMIs per cell) or very high (thresholds ranged from 20,000 to 50,000 UMIs per cell)
2. Number of genes detected per cell: Cells with too few detected genes were filtered out (thresholds ranged from 300 to 1000 genes per cell).
3. Percentage of UMIs originated from the mitochondrial genes: cells with more than a library-specific threshold (ranged from 2% to 30%) for mitochondrial UMI percentage were filtered out.

The count matrix of each library was then normalized by proportional fitting prior to log transformation followed by an additional proportional fitting, as recommended^138^ (https://github.com/pachterlab/BHGP_2022).

### Doublet removal

Four doublet removal methods were used to detect doublets, using both RNA and ATAC modalities. For scATAC-seq data, we used ArchR’s “addDoubletScores()” with the default parameters to identify doublets based on their similarity to the synthetic doublets created by mixing reads from combinations of individual cells. Additionally, we used AMULET^139^ (commit 9ce413f) with default parameters to detect multiplets (including homotypic ones) by identifying nuclei with significantly more loci containing more than two reads. For RNA-seq data, for each library, we initially assigned doublet scores to each cell using Scrublet with the default parameters. The automatically generated doublet score thresholds were then evaluated by plotting the assigned doublet scores in histograms and visualizing the scores on the 2D embedding of the cells. The automatically detected thresholds were retained for doublet annotation if they were supported by the visualization; otherwise, we adjusted the thresholds based on the visualization. Furthermore, for libraries containing cells from multiple donors, we detected doublets by clustering cells based on their genotype using Souporcell^140^ (commit 54fd312) default parameters. This method allowed us to identify doublets originating from genetically distinct samples, regardless of their gene expression profiles. Nuclei annotated as a multiplet by any of these four methods were removed from the downstream analysis.

### Sample demultiplexing

To control for technical variability, maximize flow cell utilization, and facilitate doublet detection, we used a combinatorial pooling strategy for samples with sufficient quantity. This strategy allowed us to decode the original donors of the cells downstream by the unique genotype information of each donor. Specifically, this pooling strategy involved mixing cells from multiple donors (typically 2 or 3) in various combinations (e.g., A+B, B+C, and C+A). To attribute the cells to their original donor, we leveraged the cell clustering by genotype generated by Souporcell. For each pooled library, we selected at least 15 informative loci with homozygous genotypes (reference or alternative) across the cell clusters generated by Souporcell. We then combined cell clusters from libraries with overlapping donors and clustered the cell clusters based on their genotypes at the selected loci. Finally, cell clusters from different libraries with matching genotypes were assigned to donors shared by these different libraries. For samples that were insufficient for different pooling combinations, we pooled them together by their PCW.

### Sex determination of the donors

We determined the sex of the donors by evaluating the expression levels of sex specific genes in the cells from the donors. Genes used for identification of male samples included *ZFY*, *DDX3Y*, *UTY*, *USP9Y*. *XIST* was used for identifying female samples.

### Assignment of cells to major cell types

Following quality control and doublet removal, cells from all libraries were combined for cell type identification. The combined count matrix was normalized using the method described in **scRNA-seq quality control and normalization.** Clustering was performed at three resolutions (0.2, 0.4 and 0.6) using the original Louvain algorithm implemented in the “FindClusters()” function of Seurat^141^ (v4.3.0). We visualized the relationships between clusters across these resolutions using Clustree^142^ (v0.5.0) and selected 0.2 as the resolution for downstream analysis. The clusters were assigned to the following preliminary cell types based on the expression of known marker genes: ACM (*MYH6*, *GJA5*, *NPPA*), VCM (*MYH7*, *MYL2*, *FHL2*), Endothelial and endocardial cells (*PECAM1*, *CDH5*, *UNC5B*), mural cells and fibroblast-like cells (*COL1A1*, *COL1A2*, *DCN*, *LUM*, *MYH11*, *ELN*), epicardial cells (*TBX18*, *UPK3B*, *WT1*), immune cells (*CD69*, *SKAP1*, *LEF1*, *LYVE1*, *CD68*, *PTPRC*, *CD74*), and neuron and neural crest cells (*NRXN1*, *SOX10*, *TFAP2B*, *TH*, *FOXD3*). Clusters highly express genes indicative of contamination, such as *HBG1*, *TSPAN5*, *ALB*, were removed.

### Clustering and cell type annotation

We combined cells across all samples for clustering and cell type annotation. For all cell clustering and cell type annotation analysis, we used count-splitting^143^. Specifically, we split the count matrix of each preliminary cell type into a training set and a test set of equal sizes using negative binomial splitting implemented in countsplit^143^ (v4.0.0). The gene-specific overdispersion parameters were estimated for all genes using the “vst()” function from sctransform^138,144^ (v0.3.5) with the “n_genes” parameter set to NULL.

Normalization of the training matrix was performed as described previously in **scRNA-seq quality control and normalization**. To cluster the cells, we first identified the top 2000 variable genes using Seurat’s “FindVariableGenes()” function. We then excluded from the list ribosomal genes (“^RPS.*|^RPL.*”), mitochondrial genes (“^MT-*”), genes associated with cell cycling (provided by cc.genes in Seurat), and sex-specific genes (identified by extracting genes unique to the X or Y chromosome from GENCODE v29 made available by ENCODE). This step aimed to prevent these genes from driving clustering while allowing them to be detected as differentially expressed genes (DEGs). Clustering was performed at 10 resolutions (0.1 to 1) using the original Louvain algorithm implemented in the “FindClusters()” function of Seurat^141^ (v4.3.0). The resolution was selected based on the relationships between clusters across these resolutions as visualized by Clustree^142^ (v0.5.0).

For each identified cluster, we used the test matrix to identify DEGs (calculated by Wilcoxon rank-sum test) and evaluate the expression of known marker genes. Additionally, using the original matrix, we evaluated the UMI count scaled by the corresponding sequencing library. Clusters with compromised quality were removed. These include clusters with high expression of genes indicative of contamination (e.g., *HBG1*, *TSPAN5*, *ALB*), mitochondrial or ribosomal genes, markers not consistent with the assigned preliminary cell types (e.g., cardiomyocyte markers in immune cell types), or a high proportion of cells with high UMI counts from each library (these might be doublets or multiplets). After removing these compromised clusters, normalization and clustering were repeated to identify and annotate fine-grained cell types/states. This process was repeated if any newly formed clusters showed signs of compromised quality. The markers used for each fine-grained cell type and state can be found in **Figs. S3-8**. The 2-dimensional representations of the cells were generated using UMAP as implemented in Seurat^141^ (v4.3.0).

Of note, we opted to not correct for batch effects associated with experiment or donor due to concerns of removing biological variation by over-correction. We noted that the clustering results were largely not influenced by experiment or donor: no clusters exclusively comprised cells from a specific library or donor.

### Mapping the single cell data to spatial transcriptomics data

The developmental staged-matched (PCW 12) fetal heart multiome data was mapped to MERFISH spatial transcriptomics data^48^ using the unbalanced optimal transport (OT) algorithm, as implemented by Moscot (multiomics single-cell optimal transport^145^). Suppose the gene expression readout of the multiome data is XεR^NXG1^ (N cells and G_1_genes) and gene expression readout of the MERFISH spatial data is YεR^NXG2^ ( Mcells and G_2_genes). This process then generates a coupling matrix,PεR^NXM^ that encodes the globally optimal assignment of all N multiome cells to the M MERFISH spatial location. Specifically, both our multiome data and the MERFISH data were filtered to include only the 231 genes common to both modalities, with known cell cycle genes excluded. The total counts of each cell were then normalized to 1e^4^, followed by log1p transformation. Principal Component Analysis (PCA) was performed on the combined normalized expression matrix to obtain a shared low-dimensional embedding for all cells, with the number of principal components set to 30. Because the cell type distributions in the MERFISH heart sections differ from those in our multiome dataset, applying standard OT directly violates the requirement for equal marginal mass, which would degrade the mapping results. To address this, we used an unbalanced OT framework that relaxes the marginal constraints as follows: [inline] where p is the coupling matrix, Cis the cost matrix computed from pairwise distances in the shared PCA space, a and b are the marginal distributions over MERFISH and multiome cells, respectively, *p_a_* and *p_b_* are the hyperparameters controlling the allowed mass unbalance. Following the implementation of OTT (Optimal Transport Tools) package, we represent *p_a_* and *p_b_* using τ_a_=*p_a_*/(€+*p_b_* )and τ_b_=*p_b_*/(€+*p_b_* ). Since we expect the cell population measured by MERFISH to be a subset of the cells measured by multiome, but not vice versa, we fixed the MERFISH marginal to be balanced by setting τ _a_ to 1 (corresponds to set *p_a_*to∞), while allowing the multiome marginal to be relaxed by tuning τ _a_ , and perform a grid search over τ_b_ε(0,1] to maximize the average forward and backward label-transfer score. For each candidate τ _b_ , cell type labels were transferred from the multiome atlas to MERFISH cells by summing the OT coupling weights from multiome cells within each anchor cell-type group. Each MERFISH cell was assigned to the anchor label with the highest total coupling weight. The forward label-transfer score measures the recovery of true MERFISH cell types, defined as the fraction of cells within each MERFISH anchor cell type that were assigned to the correct transferred label. The backward label-transfer score measures the purity of transferred labels, defined as the fraction of cells assigned to each anchor label that also carries the corresponding true MERFISH label. The final score was computed by averaging these two quantities. The anchor cell type matching is in **Table S3**. After grid search, τ _b_was set to 0.8. This choice guarantees that every cell in MERFISH data is mapped, while allowing multiome cells not represented in the MERFISH section to receive little or negligible transported mass.

### Predicting enhancer-gene regulatory interactions with scE2G

We used the scE2G model (https://github.com/EngreitzLab/scE2G)^56^ to predict enhancer–gene regulatory interactions within each cell type. The scE2G model builds upon the previously established Activity-by-Contact (ABC) and ENCODE-rE2G models and is adapted for single-cell applications. The scATAC-seq fragments of each fine-grained cell type were pseudo-bulked. Cell types with less than 3 million fragments were excluded, affecting 11 cell types. The pseudo-bulked scATAC-seq signals (“activity”) and a Hi-C contact map averaged across human cell types (“contact”)^58^ were used to compute the ABC score for each cell type. To incorporate the gene expression information and take advantage of the single-cell nature of the input dataset, the Kendall correlation between chromatin accessibility and gene expression across single cells within each fine-grained cell type was calculated. These correlation coefficients were subsequently integrated with corresponding ABC scores to improve calibration across varying sequencing depths.

The integrated ABC score and Kendall correlation, along with 5 additional features, were used as input to logistic regression models to predict CRISPRi-validated enhancer-gene pairs in K562 cells. The additional features include 1) the number of TSSs between each enhancer and the promoter of the target gene, 2) the number of peaks between each enhancer and the promoter of the target gene, 3) chromatin accessibility at the promoter, 4) number of peaks within 5 kb of the enhancer, and 5) whether the target gene is ubiquitously expressed. The trained scE2G model was applied to score each peak-gene pair in the 90 cell types.

The resulting scE2G scores were then quantile normalized. In our analysis, we excluded genes that are in the following categories: long noncoding RNAs (‘^LINC’), gene isoforms (‘-AS’), microRNAs (‘^MIR’), small nuclear RNAs (‘RNU’), genes of uncertain functions (‘^LOC’), and also genes with TPM less than 1 in the respective cell type. Enhancer-gene pairs with quantile-normalized scE2G scores greater than or equal to 0.173 were considered as predicted enhancer-gene regulatory interactions.

### Overlapping scE2G-predicted enhancers with previously validated enhancers

To identify previously validated enhancers in our dataset, we retrieved coordinates of previously validated enhancers from their original publications. If the coordinates were in hg19, we converted the coordinates to hg38 using the UCSC LiftOver tool. Additionally, we downloaded active cardiac enhancers in murine hearts from the VISTA Enhancer browser (https://enhancer.lbl.gov/vista/browse, accessed January 23, 2025). These cardiac enhancers were filtered to only include enhancers found in humans (Organism=Human). We then used “bedtools intersect” to intersect scE2G-predicted enhancers with validated enhancers. Finally, enhancers were considered to overlap if they overlap by at least 250 base pairs.

### Validation of KDR enhancer with CRISPRi-TAP-seq

#### sgRNA library design and cloning

For the KDR enhancer, we took the summit of the ATAC peak as the central point and extended 150-bp in either direction to design single-guide RNAs (sgRNAs). We designed up to 15 independent sgRNAs according to our established pipeline^146–148^. To target promoters, we created a 500-bp region spanning -250 to +250 relative to the TSS and designed up to 10 independent sgRNAs per promoter. As negative controls, we included 12 NT sgRNAs that lack matching sequence in the genome (**Table S21**).

Pools of sgRNA oligos containing the guide protospacer sequence flanked by partial human U6 promoter (TATCTTGTGGAAAGGACGAAACACCG) and partial scaffold (GTTTAAGAGCTAAGCTGGAAACAGCATAG) sequences were synthesized by Agilent Technologies. The guide pool was amplified with a forward primer (GGCTTTATATATCTTGTGGAAAGGACGAAACACCG) and a reverse primer (CTAGCCTTATTTAAACTTGCTATGCTGTTTCCAGCTTAGCTCTTAAAC). This amplicon was cloned into the sgOpti-CS vector via Gibson assembly and bacterial electroporation. The plasmid library was sequenced on Illumina MiSeq standard v2 50-cycle kit and shown to include all designed sgRNAs with relatively equal coverage of each.

#### Lentivirus generation

550,000 HEK293T cells per well were seeded on 6-well plates and incubated for 24 hours. Then, a DNA mix was prepared containing 900 ng psPAX2 packaging vector (Addgene #12260), 360 ng pMD2.g VSV-G envelope vector (Addgene #12259), and 1.2 µg of purified plasmid library. These were transfected using 192 µL of OMEM and 5.8 µL of X-tremeGENE HPTM DNA Transfection Reagent (Millipore Sigma #06366236001). Viral supernatant was harvested 48 hours later with 0.45 µm filtration and immediately stored at −80°C.

#### Lentiviral infection of WTC11Ci hiPSCs

WTC11Ci cells were transduced at a multiplicity of infection (MOI) of 1 at a coverage of >500 transduced cells per gRNA in two biological replicates. Transduction was performed by adding lentivirus and polybrene (Millipore Sigma #TR-1003-G) to a final concentration of 8 µg/mL into the cell media for 24 hours. Then, media was replaced with media containing 0.5 µg/mL puromycin (Sigma Aldrich) for a total of 72 hours of selection, with media replacement every 24 hours. Cells were then passaged in 0.1 µg/mL puromycin media to ensure complete selection.

#### Differentiation and cell harvest

The infected WTC11Ci hiPSCs were treated with 1 µg/mL doxycycline (Selleck) to induce expression of dCas9-KRAB two days before passage. All media after this contained doxycycline to maintain dCas9-KRAB expression. Cells were passaged and seeded for differentiation in multiple 6-well plates at the optimal density as determined above in mTeSR Plus media containing 10µM rock inhibitor and 1 µg/mL doxycycline. After 24 hours, day 0 (iPSC) cells were harvested using accutase and quenched with equal volume of mTESR Plus media and centrifuged. Cell pellets were then resuspended in cold Bambanker and viably frozen. Cells in the rest of the plates were differentiated into arterial endothelial cells using a protocol published by Loh et al^149^. On day 2 (mesoderm) and day 4 (endothelial), cells were viably frozen in the same manner.

#### Data analysis

Raw FASTQ files for the mRNA and sgRNA libraries were processed through CellRanger. To assess the relationship between gene expression changes and CRISPR perturbations, we used a modified version of SCEPTRE^76^(v0.10.1) with the following parameters: high MOI (given that most cells had a median of 2 guides), cis pair discovery with distance threshold of 1 Mb, without positive control pairs specified, and with control group as unperturbed cells. To determine the net effect of targeting a certain element and to calculate the grouped sgRNA fold changes, we ran the “union” sgRNA integration strategy. To assess the individual effects of sgRNAs, we ran singleton sgRNA integration (meaning each sgRNA was analyzed independently for its perturbation effect). We then generated the log2 fold changes and p values (with Benjamini-Hochberg correction) for each sgRNA group. Log2 fold changes were calculated by comparing *KDR* expression in cells with a specific sgRNA to the average *KDR* expression in cells without that specific sgRNA.

### Identification of overlapping enhancers

To identify predicted enhancers likely representing the same active chromatin regions, we grouped enhancers into enhancer groups based on location. Specifically, we sorted all predicted enhancers (including all cell types) by length and iteratively merged pairs that overlapped by at least 50% of the shorter enhancer’s length. After each merge, the merged enhancer became the new candidate for subsequent merging. This process continued until no further overlaps were found. All predicted enhancers contributing to the same merged enhancers were considered to be in the same enhancer groups and were considered overlapping.

### Learning models to predict base-resolution cell type-specific scATAC-seq profiles from DNA sequence and identifying TF motif instances with ChromBPNet

We applied the ChromBPNet model^63^ to identify TF motif instances and predict the effects of noncoding variants on chromatin accessibility. Our application of this model to identify important TF motifs per cell type and specific TF motif instances involves four steps: (1) Training a model to predict chromatin accessibility from DNA sequence in each cell type; (2) Identifying sequences predicted to be important for chromatin accessibility by the model; (3) Identifying recurrent sequences (motifs) from the model *de novo*, and annotating them with possible TFs using existing motif databases; and (4) Identifying specific instances of these motifs across all chromatin accessible elements:

1. To predict single nucleotide-resolution read count profiles from scATAC-seq data, we trained ChromBPNet (v0.1.7) models (https://github.com/kundajelab/chrombpnet)^63^ — sequence-to-profile convolutional neural networks — for each of the cell types. ChromBPNet uses one-hot-encoded DNA sequence (A = [1, 0, 0, 0], C = [0, 1, 0, 0], G = [0, 0, 1, 0], T = [0, 0, 0, 1]) in a 2,114-bp window around scATAC-seq peaks (specifically, candidate elements defined by scE2G above) and is trained to predict scATAC-seq pseudobulk Tn5 insertion counts for the central 1,000 bp. In order to correct for the Tn5 bias, the ChromBPNet model also uses a Tn5 bias track as input, which was obtained from the ENCODE Project (ENCSR868FGK). The ChromBPNet model has two heads, which correspond to the total Tn5 insertion counts over the 1000 bp region and to the profile — a multinomial probability of Tn5 insertion counts at each position of the 1000 bp sequence, respectively. We used a 5-fold chromosome hold-out cross-validation with the training, evaluation, and test chromosomes used for each fold as follows. Test chromosomes: fold 0: [chr1], fold 1: [chr19, chr2], fold 2: [chr3, chr20], fold 3: [chr13, chr6, chr22] & fold 4: [chr5, chr16]. Validation chromosomes: fold 0: [chr10, chr8], fold 1: [chr1], fold 2: [chr19, chr2], fold 3: [chr3, chr20] & fold 4: [chr13, chr6, chr22]. Performance of the count head was evaluated based on Pearson and Spearman correlation of observed and predicted log(total counts). Performance of the profile head was evaluated using the Jensen-Shannon Distance (JSD) between the normalized base-resolution coverage profile (converted to a probability distribution over positions) and the predicted multinomial profile distribution from the model (**Table S20**).
2. We used the DeepLIFT/DeepSHAP algorithm^61^ in order to interrogate ChromBPNet models and estimate the predictive contribution of each nucleotide in a query sequence to the predicted log(total counts) from the model. The base-resolution contribution scores across any query sequence are an additive decomposition of the difference of the predicted log(total counts) for the query sequence and the predicted log(total counts) for 20 dinucleotide preserving shuffled versions (neutral reference) of the query sequence i.e. a log fold change. For each cell type, we obtained DeepLIFT/DeepSHAP contribution scores for fold 0.
3. We then used TF-MoDISco (0.5.16.4.1) (https://github.com/kundajelab/tfmodisco)^62^ to identify non-redundant motifs by clustering high-confidence subsequences with high contribution scores (seqlets) across all open chromatin peak regions for each cell type (**Table S18**). The 500-bp windows surrounding the peak centers were considered for motif discovery with fifty thousand maximum seqlets per metacluster. Following identification of motifs *de novo*, we annotate these motifs to candidate TFs or TF families by comparing their contribution scores against the MEME version 4 motif database using TOMTOM^150^.
4. In order to call the predictive instances (sequence matches with high contribution) of these motifs with high sensitivity and specificity, we scanned through contribution scores in open chromatin regions with the FiNeMo (v0.25) hit caller (https://github.com/austintwang/finemo_gpu), using alpha 0.7 and 5 as a hit coefficient threshold for reporting the instances (**Table S19**).

### Predicting effects of noncoding variants on chromatin accessibility with ChromBPNet

In order to score and identify variants predicted to affect chromatin accessibility, we used the ChromBPNet models for each cell type to predict base-resolution scATAC-seq coverage profiles for 1 kb genomic sequence windows containing reference and alternate alleles of selected variants. The effect size of each variant was estimated using two measures: the log2 fold change of the total predicted coverage (total counts) over each sequence window containing the reference and alternate allele, and the JSD between the base-resolution predicted probability profiles for the reference and alternative sequences to measure a change in the profile shape.

Statistical significance was estimated for both of these scores on empirical null distributions of variant scores that were computed as follows. We shuffled the 2114 bp sequence centered at each variant multiple times while preserving the dinucleotide frequency, made 2 copies of each shuffled sequence, then inserted each allele of the variant at the center of the shuffled sequence to create a set of 100 thousand null variants in total. We scored each of these null variants with each model using the same procedure that we used to score each observed variant. Then, for each observed variant, we calculated the proportion of the null variants that had an equally high or higher (more extreme) score to generate an empirical p-value for both the log2 fold change and JSD scores. The code base for scoring variants is at https://github.com/kundajelab/variant-scorer.

### Defining gene programs with cNMF

To identify co-expressed genes (programs) within the heart atlas, we used consensus NMF^59^ (v1.3.4). We and others have previously benchmarked cNMF and found that it performs well at identifying sets of genes that are enriched for genetic variants for complex diseases^65,151^. As such, we aimed to use the same strategy here to identify sets of co-regulated genes that we could use to annotate and interpret genetic variants associated with heart diseases. To increase the resolution of the programs identified, the cells in the heart atlas were grouped into cell type groups as specified in **Table S6**.

To create the input count matrix for cNMF for each cell type group, we first excluded gene categories that are less informative for the analysis. These categories include long noncoding RNAs (‘^LINC’), gene isoforms (‘-AS’), microRNAs (‘^MIR’), small nuclear RNAs (‘RNU’), genes of uncertain functions (‘^LOC’), and genes lacking Ensembl stable ID or Entrez ID (bioconductor-org.hs.eg.db, v3.17.0). Furthermore, we filtered out cells with low gene abundance (fewer than 200 expressed genes). This threshold was more lenient than standard recommendations because the cell had been through the quality control step as described in **scRNA-seq quality control and normalization**. Finally, genes expressed in less than 10 cells were removed to focus on genes with a broader expression pattern (260-8000 genes after filtering).

The implementation of cNMF is described in Kotliar et al^59^. Briefly, NMF decomposes each count matrix (cell × gene) into two lower rank matrices. One matrix represents the contribution of each gene to each program (gene contribution matrix), and the other represents the proportions in which the programs are combined within each cell (usage matrix). To overcome the non-deterministic nature of NMF and achieve robust results, cNMF ran multiple replicates of NMF on the same normalized dataset with different randomly chosen seeds while using the same K, a parameter that determines the number of programs. The results are then averaged over multiple replicates after the outlier programs are removed.

Following the recommendation outlined by Kotliar et al^59^, the “prepare” step of the algorithm was run with the following parameters: ‘cnmf prepare -k 15 18 21 24 27 30 33 36 39 41 43 45 47 --n-iter 100 --seed 42”. This step tested 13 different K values with 100 replicates for each K. Seed 42 was used to initialize the random seeds for the replicates. The “consensus” step was run with the parameter “--local-density-threshold 0.1”, where 0.1 is the Euclidean distance threshold used to identify outlier programs and was chosen based on the clustergram diagnostic plot generated by cNMF.

Finally, to determine the optimal K for each cell group, we sought the smallest K that maximized the number of unique gene ontology (GO) terms enriched in the programs calculated by Gene Set Enrichment Analysis (clusterprofiler^152^, v4.10.0; r-msigdbr^153^ v7.5.1: category: C5, subcategory: BP), and at the same time, minimizing the difference between the original TPM matrix and the matrix reconstructed by multiplying the gene contribution matrix (in TPM unit) with the usage matrix.

Programs representing ribosomal contamination, cardiomyocyte contamination, or expressing other markers indicative of contamination (e.g., *ABL*1, *HBG1*), as well as programs expressed in less than 5% of any cell type, were removed from further analysis. This resulted in a total of 253 programs for downstream consideration.

The K chosen for each cell type group are listed below:

ACM: 39; core_conduction: 36; Endocardial_endothelial: 49; Epicardial: 39; FB: 39; MuralCells: 41; NC: 41; VCM: 43; Lymphatic_EC: 15; TzConductionCells: 30; ImmuneCells: 41.

### Identifying TF motifs that correlate with gene programs

We projected each cell to the motif space (cisbp, chromvarmotifs^154^ v0.2.0) using chromVar^155^ (v1.16.0). Next, we identified TFs whose per-cell motif accessibility significantly correlated (Spearman correlation > 0.1) with the expression of the program and were among the top 300 genes most highly associated with the program. These correlated TF motifs are intended as an annotation tool and do not necessarily imply that these TFs directly regulate genes in the program.

### Cell-cell interactions

We employed CellChat (v2.1.2)^156^ to infer cell-cell interaction (CCI) relationships, and more precisely the cell-type to cell-type interaction in this study (**Table S9**). The log1p transformed sequencing-depth normalized gene expression counts and cell type annotations (all except for pulmonary endothelial cells) were used as primary inputs. CellChat curated a comprehensive database of ligand-receptor pairs, which serves as the knowledge foundation for analyzing the intercellular communications. Each record in this database consists of a ligand-receptor pair and their corresponding encoding genes. As a single ligand or receptor protein may have multiple subunits, it may correspond to multiple genes. These ligand-receptor pairs are systematically categorized into distinct CCI pathways. For instance, interactions such as DLL1-NOTCH1 and JAG1-NOTCH2 are consolidated into the “NOTCH” pathway, while pairs involving WNT ligands (e.g., WNT1-FZD1, WNT3-FZD5) are grouped into the “WNT” pathway. In our analysis, we subset the database records to the expressed ligand and receptor genes in our atlas. CellChat implements a computational approach to analyze CCI. The analysis generates a three-dimensional tensor of ligand-receptor interaction strengths with dimensions K × K × N, where K represents the number of cell types, and N denotes the number of ligand-receptor pairs. The interaction strength calculation involves several steps: 1) Computation of ensemble average gene expression within= each cell type; 2) Implementation of a Hill function-based model to quantify interaction strengths. The Hill function model integrates multiple biological factors: expression levels of ligands and receptors; cell type population size; protein subunits; presence of co-stimulatory and inhibitory receptors; effects of extracellular agonists and antagonists. This comprehensive modeling approach ensures that the calculated interaction strengths reflect the biological complexity of cell-cell communication systems. CellChat aggregates ligand-receptor pair level activities to pathway level activities to quantify cell-cell communication through three main steps. First, it identifies significant ligand-receptor pairs through statistical tests that randomly shuffle cell type labels, keeping only pairs with p-value <0.05. Then, using the computeCommunProbPathway() function, these significant pairs are grouped by their signaling pathways and their strengths are summed, creating two matrices (incoming and outgoing signals) where rows represent types, columns represent CCI pathways, and values show the summed strengths of all the corresponding ligand-receptor in that pathway. Note that the row sum of these two matrices are used for feature selection. Finally, these two matrices are added together to create a final signaling strength matrix, which reveals both the primary communication modes used by cells and the similarities in their communication behaviors. Reproducible Jupyter notebook for the CCI analyses can be found in the GitHub repository https://github.com/chansigit/heartmap-cci-reproduction.

### Annotation of disease genes for CHD and adult-onset cardiac diseases

We downloaded the initial list of CHD genes and their associated CHD phenotypes and classifications from CHDGene^9^ (https://chdgene.victorchang.edu.au/, accessed May 16th, 2024). In CHDGene, “Genes are only included in the database if variants in the respective gene have been reported as the monogenic cause for CHD (isolated or in the context of a syndrome) in at least 3 independent familial or sporadic cases in one or more separate publications.”^9^ We modified the original CHD classification as follows (**Table S11**):

- **ASD**: we merged genes linked to ASD and ASD with minor abnormalities into a single “ASD” category. We removed *LTBP2* and *TMEM260* from this category because ASD is not a listed phenotype for *LTBP2*, and *TMEM260* is associated with persistent left superior vena cava, which might cause ASD through a different mechanism from other ASD causal genes.
- **VSD**: we combined genes linked to VSD and VSD with minor abnormalities into a “VSD” category and excluded genes associated with patent foramen ovale, patent ductus arteriosus (PDA), and BAV. We included *DNAH5* into the list for its VSD association.
- **Valve defects**: we renamed the “Obstructive lesions” category to “Valve defects” category. This group now includes all genes with valve lesions in their CHD phenotypes. Genes solely associated with interrupted aortic arch or coarctation of the aorta but not any of the valve lesions were moved to the malformation of the OFT category.
- **Malformations of the OFT**: genes associated with interrupted aortic arch or coarctation of the aorta were added to this category.

We also analyzed a recent list of 60 genes identified in a large cohort of 11,555 probands as having a significant burden of heterozygous damaging variants, driven by contributions from *de novo* and transmitted mutations.

We obtained the causal genes for adult-onset cardiac diseases from ClinVar: https://ftp.ncbi.nlm.nih.gov/pub/clinvar/gene_condition_source_id (downloaded on July 29th, 2024).

### Testing disease genes for preferential expression in particular cell types

We defined preferentially expressed genes as the top 300 most highly expressed genes among those with z-score > 1 (top 16%) across all cell types, after excluding all cell cycling cell types, pulmonary endothelial cells and lymphoid cells, as well as genes that are in the following categories: long noncoding RNAs (‘^LINC’), gene isoforms (‘-AS’), microRNAs (‘^MIR’), small nuclear RNAs (‘RNU’), genes of uncertain functions (‘^LOC’), and ribosomal genes (“^RPS” and “^RPL”). To empirically validate this specific threshold choice and evaluate the parameter robustness of our downstream results, we performed a sensitivity analysis across 7 distinct threshold increments: the top 100, 200, 300, 400, 500, 1,000 genes, and a relaxed background baseline consisting of all genes with a z-score > 1 within the corresponding cell type. To test whether the disease genes are enriched in any particular cell type, we conducted a one-sided Fisher’s exact test. Background genes were defined as those expressed in any of the considered cell types (TPM>1). We corrected for multiple testing across all cell types within each disease category (high-confidence CHD genes from CHDGene and adult-onset cardiac diseases) using Bonferroni correction. VICs were significantly enriched for CHD genes at all thresholds (except z-score > 1), and the top 300-gene threshold maximized the number of cell types discovered as significantly enriched for CHD genes (**Fig. S16G**).

### Assessing cell type heritability enrichment with stratified LD score regression

We used stratified LD score regression^157^ (S-LDSC, v1.0.1) to estimate the enrichment of disease or trait heritability within cell type-specific enhancers. These enhancers were defined as all distal enhancers predicted by scE2G for each cell type. We preprocessed each GWAS summary statistics using the munge_sumstats.py provided by LDSC^158^ and filtered to retain only SNPs curated by HapMap 3 as recommended^77,158^. We then performed S-LDSC with our cell type-specific scE2G enhancer annotations along with the baseline model (v.2.2). The 1000 Genome EUR Phase 3 genotype data was used as the LD reference panel. Full S-LDSC results can be found in **Table S15.**

### Fine-mapping of GWAS signals for quantitative cardiac traits

GWAS were conducted for deep learning-based left atrial (LA) measurements^159^ and left ventricular (LV) measurements^160^. Deep learning semantic segmentation (pixel labeling) models for UK Biobank cardiovascular magnetic resonance imaging (MRI) 2-, 3-, and 4-chamber long-axis views and short axis stacks were trained using PyTorch^161^. These were reconstructed into 3D surfaces separately for the LA^162^ and LV^37^ using Poisson surface reconstruction^163^, permitting volume measurements in systole and diastole. GWAS was conducted with REGENIE using data from 63,196 participants with LA and LV measurements who did not have atrial fibrillation (which alters cardiac chamber volumes due to eliminating atrial filling and contraction). Adjustment was made for covariates including age, age^2^, genetic PC1-10, sex, the genotyping array, and the imaging device.

Semantic segmentation was performed to measure aortic root diameter and ascending aortic diameter from UK Biobank MRI, and GWAS was conducted with REGENIE using data from 62,936 participants with aortic root diameter measurements, 62,716 with ascending aortic diameter measurements^164^. A meta-analysis of thoracic aortic aneurysm disease GWAS was produced using the METAL software package^165^ on data from three sources: Million Veteran Program^166^, FinnGen release #10, and UK Biobank (effective sample size: 53,516). Multi-trait analysis of GWAS (MTAG)^167^ was conducted using summary statistics from the three measurements (root diameter, ascending and descending thoracic aortic diameter) and the thoracic aortic aneurysm meta-analysis^91^ .

Semantic segmentation was performed to measure aortic valve hemodynamic measurements (AVA, mean gradient, and peak velocity) from UK Biobank MRI^88^, and GWAS was conducted with REGENIE using data from 59,569 participants with AVA measurements and 59,571 with mean gradient and peak velocity measurements^89^. Adjustment was made for covariates including age, age^2^, genetic PC1-10, sex, the genotyping array, and the imaging device. A meta-analysis of aortic stenosis disease GWAS was produced using the METAL software package configured to account for overlapping samples^165^ on data from four sources: Million Veteran Program^168^, FinnGen release #12, the TARGET multi-cohort GWAS^169^, and UK Biobank (maximum effective sample size: 164,971). MTAG^167^ was conducted using summary statistics from the hemodynamic measurements (AVA, mean gradient, and peak velocity) and the aortic stenosis meta-analysis^90^.

We defined fine-mapping regions based on a 3 Mb window around each lead variant and merged regions if they overlapped^170^. LD was then computed using LDstore v2.0^171^ for these regions using in-sample UK Biobank imputed genotypes. Fine-mapping was then performed using SuSiE-inf^172^ (v1.2) with default settings on the phenotypes derived from UK Biobank MRI data with in-sample, covariates-adjusted LD (see sections above for covariates). Credible set coverage is set to 0.95 and purity is set to 0.5, meaning that 95% of the credible sets contain a true causal variant and that the minimal pairwise LD between all variants in each credible set is 0.5.

### Preparing variants for overlapping with scE2G-predicted enhancers

We included three sets of variants for our analysis. The first set was fine-mapped using SuSiE-inf as described in Methods above. The second set was fine-mapped using SuSiE as implemented by FINNGEN^173^. The third set underwent Linkage Disequilibrium (LD) expansion. See **Table S14** for the method used for each trait.

Specifically, for variants that were fine-mapped using SuSiE-inf, we first removed all variants that are not in a credible set (cs= −1) and have posterior inclusion probability (PIP) values below 0.9. In the remaining variants that are in credible sets (cs != −1), the variants with the highest PIP were designated as the lead variants of their respective credible sets. Additionally, variants outside credible sets (cs= −1) but with PIP greater or equal to 0.9 were individually considered as lead variants.

For variants fine-mapped by FINNGEN^173^, the summary statistics and variants within 95% credible sets were downloaded from gs://finngen-public-data-r8. Both fine-mapped datasets were filtered to include only variants with PIP over or equal to 0.1.

For GWAS results with only summary statistics or lead SNPs available, we performed LD clumping using PLINK (v1.9) with parameters “--clump-p1 5e-8 --clump-p2 0.01 --clump-r2 0.1” to identify a single most significant SNP per LD block. These SNPs were defined as the lead SNPs and subsequently expanded to include variants in LD (r² ≥ 0.9) using PLINK parameters “--ld-window-kb 1000 --ld-window 99999 --ld-window-r2 0.9” and the 1000 Genome EUR Phase 3 reference panel.

### Linking GWAS variants to enhancers, target genes, cell types, and gene programs

We applied our gene regulation map to interpret noncoding GWAS variants by linking them to target cell types, enhancers, genes, and gene programs. To do so, we started with the list of fine-mapped or LD-expanded variants described above. We then linked variants to genes and cell types with scE2G. Specifically, we overlapped variants with scE2G-predicted enhancer-gene regulatory interactions and filtered for the top two target genes with the highest quantile-normalized scE2G scores for each enhancer. For this step, we also included variants linked to GWAS signals that overlapped coding regions or splice sites (n=129 GWAS signals) (“AnyVariantInCSOverlapCoding” and “AnyVariantInCSOverlapSpliceSite” in **Table S16**). This approach builds on previous methods we have developed to interpret GWAS signals using enhancer-to-gene (E2G) maps^56,58,65,147^, extending them to enable interpreting variants across a tissue atlas including many dozens of cell types. This yielded a list of 750 unique GWAS signals linked to one or more target genes.

To further annotate this set of 750 GWAS signals, we assessed whether the predicted target genes were part of a cNMF gene program in the same cell type as the scE2G prediction. We considered a gene to be part of a gene program in a given cell type if the gene was ranked among the top 300 genes in the program based on importance scores and at least 10% of cells expressing the gene program (defined as having a program usage score > 10%) belonged to the corresponding cell type.

For a subset of variants of interest, we interpreted the local effect of the variant on chromatin accessibility in that cell type using ChromBPNet. Specifically, we predicted the fold-change in chromatin accessibility at the predicted enhancer by substituting the alternative allele into the center of the ∼2-Kb input sequence of ChromBPNet. We also examined contribution scores of nucleotides across the window for both the reference and alternative alleles, and applied FiNeMo to identify predicted motif instances that are affected by the variant (see **Methods** above).

### Evaluation of gene regulation map components for noncoding variant prioritization

To evaluate the contribution of each component of our gene regulation map to prioritizing noncoding cardiac variants, we benchmarked four progressive feature sets following the method described above:

1. accessible peaks only, (2) scE2G predicted enhancers, (3) scE2G enhancers linked to target genes ranked among the top 300 of a cNMF gene program active in the corresponding cell type, and (4) variants overlapping enhancers from (3) and predicted by ChromBPNet to induce significant changes in chromatin accessibility in relevant cell types. Ground truth comprised 180 unique noncoding variants associated with quantitative cardiac traits with fine-mapping PIP > 0.90, benchmarked against a background of 98,024 unique distal noncoding variants.

Recall was calculated as the proportion of ground-truth variants captured by each feature set, and fold enrichment was defined as this recall divided by the corresponding background annotation rate. Nominal 95% confidence intervals for recall were computed using the Agresti–Coull (plus-four) method^174^. Nominal 95% confidence intervals for fold enrichment were estimated on the logarithmic scale using the Katz method and exponentiated to yield asymmetric confidence limits^175^.

### VIC trajectory and gene association analyses

We used Slingshot (v2.6) to construct developmental trajectories of VIC subpopulations^176^. Trajectories were modeled in the PCA space using normalized expression data, with Seurat-defined clusters (resolution 0.5) serving as nodes for the minimum spanning tree (MST). For each cell type, the most relevant trajectory was selected based on a heuristic that prioritized the greatest cell density and the strongest Spearman correlation with chronological developmental stages.

To link these trajectories with functional gene programs, we calculated the Spearman correlation (ρ) between Slingshot pseudotime and gene program scores. Associations were deemed significant where ∣ρ∣>0.1 and P<0.05.

Gene expression dynamics were characterized by fitting a negative binomial generalized additive model (NB-GAM) to the lineages using the tradeSeq (v1.12) package^177^. The optimal number of knots (K) for each trajectory was determined using the evaluateK function, testing K from 3 to 10 on a subset of highly variable genes (top 2,000 genes expressed in more than 5% of cells within the lineage). The final K was selected by applying the elbow method to the average Akaike Information Criterion (AIC).

The “associationTest” function was used to identify genes with significant temporal variation. P-values were adjusted using the FDR method (FDR<0.05). The magnitude of these associations was quantified by the deviance explained, representing the proportion of gene expression variability attributed to the trajectory.

### Analysis of 23 enriched high-risk CHD genes in mouse and human VICs

The murine AV valve development atlas was obtained from the Broad Single Cell Portal (accession SCP1897), which spans E7.75 hearts to E12.5 AV canals. The authors’ EndMT was used as annotated (n = 10,354 cells) with stage and pseudotime. For human VIC analysis, the log-normalized atlas was downloaded from the Heart Cell Atlas portal (www.heartcellatlas.org/foetal), which spans PCW4-20. Annotations were used as provided and VICs were selected as the “ValveInterstitialCells” fine-grain annotation. Percentage of cells with non-zero counts was computed per gene for each cell type x PCW group. In total there are 7,398 cells with 31-2,577 cells per week. For fetal heart map analysis, the 13,636 cells assigned to the five VIC subpopulations were used after linking to donor and PCW.

In all three datasets, single cell gene expression was calculated with log1p(CP10K) and the mean was determined across all the cells per group, along with percentage of cells with at least one count. Cell-type annotations and developmental stages were used as provided by the dataset authors.

### Assessing enrichment of genes associated with valve traits in Fetal VIC program 27

Genes associated with valve traits are defined as ones that are predicted to be regulated by GWAS variants linked to aortic stenosis, AVA, aortic root diameter, aortic valve peak velocity, aortic valve mean gradient, mitral valve prolapse. To test whether these genes are enriched in Fetal VIC program 27, we conducted a one-sided Fisher’s exact test. Background genes were defined as those expressed in any of the 5 VIC cell types (TPM>1).

### Perturb-seq in hiPSC-VICs

#### Design of gRNA library for Perturb-seq

To test whether CHD genes have cell-autonomous effects in VICs, we designed a CRISPRi-Perturb-seq experiment in which we knocked down each of 23 CHD genes, along with other control genes, and measured their effects on gene expression with single-cell RNA-seq. To design the library, 15-30 guides were generated to target the TSS-sites of the 23 CHD genes highly expressed in VICs/cardiac fibroblasts (Fig. 2E). Promoters were defined by TSS sites predicted by ABC or GENCODE. 200 negative control guides (100 safe-targeting^178^ and 100 negative targeting gRNAs^179^), 10 guides targeting essential genes, and 10 positive control guides (known to have functional activity based on previous CRISPRi experiments) were also included in the library.

To construct the CHD guide library, we first obtained the sgRNA protospacer sequences from the Dual sgRNA CRISPRi Libraries of the Weissman lab (Addgene #187246, #187247, #187248)^180^. Pool of sgRNA oligos containing the guide protospacer sequence flanked by partial human U6 promoter (TATCTTGTGGAAAGGACGAAACACCG) and partial scaffold (GTTTAAGAGCTAAGCTGGAAACAGCATAG) sequences were synthesized by Agilent Technologies. The guide pool was amplified with a forward primer (GGCTTTATATATCTTGTGGAAAGGACGAAACACCG) and a reverse primer (CTTATTTAAACTTGCTATGCTGTTTCCAGCTTAGCTCTTAAAC), extending the Gibson arms for subsequent cloning step into sgOpti-CS vector (Addgene #256351). Plasmid backbone was prepared by digesting with BsmBI (New England Biolabs), followed by purification with DNA gel extraction (New England Biolabs) and 1X Ampure XP SPRI bead clean-up. Gibson assembly was performed for 1 hour at 50°C using NEBuilder HiFi DNA Assembly Master Mix (New England Biolabs), 330 ng backbone, and 24 ng of purified PCR-amplified sgRNA pool in a final volume of 20 µL. The cloned plasmid was purified with 0.7X Ampure XP SPRI, electroporated into Endura competent cells (Lucigen #602422), and cells were then expanded in liquid culture for 18 hours at 30°C. Plasmid library was then obtained with the ZymoPURE II Plasmid Midiprep kit (Zymo Research #D4200). To sequence-validate sgRNAs and determine their relative abundance, we PCR amplified and sequenced the sgRNAs from the plasmid library on Illumina MiSeq standard v2 50-cycle kit. Analysis of the sequencing data showed that all designed sgRNAs were represented with relatively equal coverage of each (the difference in count frequency between the top and bottom 10th percentiles of sgRNAs was 2.5).

#### Lentivirus generation

550,000 HEK293T cells per well were seeded on 6-well plates and incubated for 24 hours. Then, a DNA mix was prepared containing 900 ng psPAX2 packaging vector (Addgene #12260), 360 ng pMD2.g VSV-G envelope vector (Addgene #12259), and 1.2 µg of purified plasmid library. These were transfected using 192 µL of OMEM and 5.8 µL of X-tremeGENE HPTM DNA Transfection Reagent (Millipore Sigma #06366236001). Viral supernatant was harvested 48 hours later with 0.45 µm filtration and immediately stored at -80°C.

#### Culturing hiPSC-Valvular Interstitial Cells (hiPSC-VICs)

Human WTC11 iPSCs (harboring doxycycline-inducible CRISPRi) were cultured and passaged after reaching confluency, then seeded into Matrigel-coated 6-well plates at a density of 100k cells per well in mTeSR plus media (Stemcell, 100-0276) with 10 µmol/L Y-27632. To achieve appropriate guide library coverage, we infected 1 million cells with the CHD guide library at a MOI of 10 (measured through digital droplet PCR). 24 hours after transfection, puromycin selection (0.8 µg/ml) was conducted for 48 hours, after which 1 million puromycin-selected cells were replated for differentiation according to a previously published hiPSC-VIC/VEC differentiation protocol.^97^

After 24 hours, the media was replaced with cardiac lateral plate mesoderm (hiCLPM) media, which was a 1:1 mixture of Neurobasal medium (Life Technologies, 21103049) and DMEM/F12 (Life Technologies, 11320-033), with 1.94% B27 (Life Technologies, 12587010), 0.97% N2 (Life Technologies, 17502048), 8 µmol/L CHIR-99021 (Selleck, S2924), and 25 ng/mL BMP4 (Peprotech, 120-05). hiCLPM media was changed every 24 hours for 72 hours total, after which heart valve differentiation media was added consisting of StemPro-34 SFM medium (Gibco, 10639011) supplemented with 200 ng/mL VEGF-165 (Peprotech, 100-20) and 1 µmol/L forskolin (Selleck, S2449). Heart valve differentiation media was changed every 24 hours for 48 hours. At day 6 of differentiation, the cells were sorted via CD144 magnetic beads (Miltenyi Biotec, 130-097-857). CD144-cells (the VICs) were cultured in DMEM (Gibco, C119955000BT) with 10% fetal bovine serum (Gibco, 10099). After 24 hours, fresh media was added with 1 µg/mL doxycycline (Selleck, 50-194-7977) to induce CRISPRi expression for 48 hours. Following CRISPRi induction, hiPSC-VICs were harvested via 0.05% Trypsin-EDTA (Gibco, 25300054), sorted for mCherry signal to denote CRISPRi activity (with gating determined by untreated hiPSC-VIC control), and approximately 2 million cells in two biological replicates were frozen in Bambanker (Fisher, 50-999-555) at a concentration of 1 million cells/ml.

#### CC-Perturb-seq in hiPSC-VICs

To measure effects of each gene perturbation on the transcriptome, we used CC-Perturb-seq as previously described^98^. In brief, CC-Perturb-seq combines combinatorial barcoding with an optimized approach for gRNA detection. In this approach, we conduct split-pool single-cell RNA-seq library preparation^181,182^, with several modifications. First, we use an extra reverse transcription primer targeting the Cas9 gRNA scaffold. Second, following whole-transcriptome amplification, we circularize short cDNA fragments and PCR to isolate the gRNAs.

Here, we first fixed hiPSC-VICs in 1% formaldehyde (Thermo Fisher Scientific, 28906) for 10 min on ice, permeabilized the cells with 0.2% Triton X-100 (Millipore Sigma, T8787) for 3 min on ice, and quenched the formaldehyde with Tris. We then washed the cells in PBS supplemented with RNase inhibitors and resuspended them in the same buffer. Immediately before use, we freshly prepared BS3 (Thermo Fisher Scientific, A39266) in water and added it to the cells at a final concentration of 2 mM. We crosslinked the cells with BS3 for 20 min on ice, followed by an additional 3-min permeabilization with Triton X-100 and quenching with Tris. We then washed the fixed cells and resuspended them in 0.5x PBS supplemented with RNase inhibitors. Fixed cells were maintained overnight at 4°C with agitation.

Following completion of the three rounds of combinatorial barcoding, we recovered 300,960 cells in 144 µL (approximately 2,090 cells/µL) and divided the cells into seven sublibraries: five containing 50,000 cells each, one containing approximately 33,000–40,000 cells, and one containing 10,000 cells. We omitted the 40-µm filtration step after Round 3 barcoding.

After sublibrary generation and lysis, we captured the barcoded cDNA on streptavidin beads, performed template switching, and amplified the barcoded cDNA. Because the protocol recommends optimizing the number of amplification cycles according to the number of cells in each sublibrary, we used cycle numbers previously optimized for hiPSC-VICs. After the five initial PCR cycles specified in the protocol, we performed two additional cycles for sublibraries 1–6 (approximately 33,000–50,000 cells each) and six additional cycles for sublibrary 7 (10,000 cells). We then separated the amplified products into the gene expression libraries (GEX) and gRNA fractions by SPRI cleanup (Beckman Coulter, B23319).

We next prepared sequencing libraries from the GEX fraction. Following fragmentation, end repair, A-tailing, and adapter ligation as described in the CC-Perturb-seq protocol, we performed the final GEX index PCR. We used nine PCR cycles for sublibraries 1–5 and 7 and ten PCR cycles for sublibrary 6. We assessed the resulting GEX libraries by Bioanalyzer (Agilent, 5067-4626) and Qubit (Thermo Fisher Scientific, Q33231).

In parallel, we used the gRNA-containing fraction to generate the gRNA libraries. During gRNA Feature PCR, which re-enriches the gRNA-derived cDNA before circularization, we performed the five initial PCR cycles specified in the protocol followed by two additional cycles for sublibraries 1–6 and three additional cycles for the 10,000-cell sublibrary (sublibrary 7).

Following gRNA enrichment, strand separation, and circularization, we performed the final indexed PCR of the circularized gRNA library using five initial cycles followed by ten additional cycles. We assessed the final gRNA libraries by Bioanalyzer and Qubit.

After library preparation and quality control, we initially sequenced the GEX and gRNA libraries on an Illumina NextSeq 550 for quality-control assessment and subsequently performed deeper sequencing on an Illumina NovaSeq X Plus. We targeted 30,000–50,000 read pairs per cell for GEX libraries and approximately 5,000 read pairs per cell for gRNA libraries.

#### Perturb-seq data analysis

To generate counts matrices, GEX reads were pseudo-aligned using kallisto bustools^183^ and gRNA reads were pseudo-aligned using kite^183,184^ given a reference list of indices (https://github.com/tkzeng/perturb_pipeline). Per sublibrary, GEX and gRNA counts matrices were combined into a single object with genes annotated using GENCODE v46. Per cell quality metrics (such as mitochondrial %, ribosomal %, total UMIs, and genes detected) were calculated to filter out low-quality cells and doublets, which retained 240,504 cells. Post-QC-filtered sublibrary counts matrices were concatenated into a single object and analyzed using scanpy^185^ (PCA, UMAP, and Leiden clustering at a resolution of 0.25).

SCEPTRE^76^ was used to assign guides to cells using the mixture model method in a high MOI setting with appropriate covariates (sublibrary, # of gRNAs per cell, # of gRNA UMIs per cell, # counts per cell, # of GEX UMIs per cell, mitochondrial %). Target knockdown was confirmed; successful perturbation was determined to be on-target knockdown >9-13% based on SCEPTRE’s differential expression testing. Following appropriate calibration with NT guides, trans analysis was conducted to determine differential gene expression of all 15,882 detected protein coding genes given a specific gRNA perturbation in comparison to a “complement” control group consisting of cells not containing guides towards that specific perturbation target. SCEPTRE fits a negative binomial generalized linear model (GLM) of the response gene’s UMI counts based on the cell covariates alone, then computes a GLM score statistic for the perturbation indicator and covariate-permutes the indicator for a p value from the tail of a skew-normal fitted to the null statistics. Benjamini-Hochberg correction was applied to all p-values and perturbation-response pairs with alpha < 0.1 were denoted as significant, with effect sizes reported as log2fold change in comparison to the complement control group.

Gene programs were inferred by cNMF^59^ (see **Defining gene programs with cNMF)** with a GPU-accelerated PyTorch reimplementation (https://github.com/EngreitzLab/PerturbNMF/tree/main), with input as the filtered counts matrix. We set K to be 30 and calculated gene spectra/gene spectra score, with the 300 top-scoring genes defined to be the makeup of the program for functional annotation.

To determine if specific gene perturbations changed program activity, we used a propensity-score-matched regression framework with appropriate covariates (mitochondrial and ribosomal UMIs, cell cycle score, total counts, # of detected genes, guide UMIs per cell, guides per cell, and sublibrary). Guides were collapsed to the target levels (ex. guides targeting the same gene were grouped together), with the 200 NT guides providing an empirical null generated by randomly sampling 15 NT guides per group without replacement. For each perturbation, logistic regression was used to estimate a propensity score of the perturbation indicator, and perturbed cells were matched 1:1 with unperturbed cells by nearest-neighbor matching. Program usage per cell was scaled and log-transformed with a program-specific pseudocount to accommodate for zero-inflation of cNMF usages. For each program, this usage value was regressed by weighted ordinary least squares (OLS) on the perturbation indicator. The calculated perturbation effect is then the coefficient of this perturbation indicator (natural-log fold change in program usage); standard errors were calculated with matched pairs which generated a two-sided Wald’s test per perturbation and program pair.

To correct for per-program bias and variance misspecification post-matching, test statistics were calibrated per program using the NT groups. This generated a directional bias and variance inflation factor which were used to calibrate p-values (β_cal = β − bias, SE_cal = SE × t_std, with two-sided p-values from the standard normal distribution). Benjamini-Hochberg procedure was used to correct for multiple testing and anything with adjusted p < 0.05 was denoted as statistically significant.

### Single guide knockdown of *MEIS2* in hiPSC-VICs

hiPSCs were transduced either with a NT gRNA or with the top-performing guide targeting MEIS2 in four biological replicates at MOI of around 10. Following 48 hours of puromycin selection, hiPSCs were seeded for differentiation into VICs at a density of 10,400 cells/cm². Following MACS separation on day 6, hiPSC-VICs were replated at a density of 35,300 cells/cm² and doxycycline (1 µg/ml) was added for CRISPRi induction for 72 hours. Cells were harvested using 0.05% Trypsin-EDTA (Gibco, 25300054) and then sorted for mCherry signal. RNA was harvested by resuspending the cell pellet in RNA/DNA Shield (Zymo, R1100-50) and submitted for bulk RNA sequencing analysis via Plasmidsaurus at 150,000 cells per replicate. DESeq2^186^ was used to determine differential gene expression of all detected protein coding genes (using Wald’s test with Benjamini-Hochberg correction of p values) which were reported as log2fold-change in comparison to the NT control condition.

### Validation of GOSR2 enhancer with Variant-EFFECTS

#### Culturing TeloHAEC Cells

TeloHAECs were purchased from the American Type Culture Collection (ATCC, CRL-4052) and cultured using the Lifeline Cell Technology Vasculife^®^ VEGF Endothelial Cell Medium Kit (VWR #102970-922). Cells were plated at a density of 1.7×10^4^ cells/cm^2^ and passaged every 2-3 days or at 80-90% confluence.

#### Design and cloning of pegRNAs for GOSR2 enhancer screen

Edits for the *GOSR2* enhancer screen were designed to target a region encompassing the rs17608766 SNP located at chr17:45013271 (all coordinates for this experiment are based on hg19). All three single nucleotide variants (T to A/C/G) were tested at the variant locus along with seven additional edits that were selected based on their predicted ability to disrupt the KLF motif surrounding this variant without introducing a repressor (**Table S22**).

The pegRNAs encoding these edits were designed using previously established criteria^64^. Briefly, these pegRNAs encode 11 bp primer binding sites and RT templates (RTT) ≤30 bp in length, including a 10-bp extension beyond the target edit. All pegRNAs in this experiment used the same spacer (GCTGGGAGTGCTTCATTCAC). The pegRNAs were ordered as oligo libraries with PCR tags (ordered from Twist Bioscience) and cloned into sgOPTI:U6-gRNA-blastR (Addgene #256626). Sub-pools of pegRNA oligos were selectively amplified using subpool-specific PCR primers, followed by a second PCR to add homology arms for Gibson assembly into sgOpti, as previously described^187^. Each PCR was followed by a 1.5X Ampure XP bead cleanup. The sgOPTI:U6-gRNA-blastR vector was digested with BsmBI, ClaI, and EcoRI-HIF (New England Biolabs) and purified with a 0.7X AMPure XP bead cleanup. We performed Gibson assembly for 1 hour at 50°C using 500 ng digested vector and 70 ng purified, PCR-amplified pegRNA library in a final volume of 40 µL. The assembled plasmid library was purified with 0.7X AMPure XP bead cleanup, then expanded and sequence validated as previously described^64^.

#### TeloHAEC infection and selection

To deliver pegRNAs, 1.5 x 10^6^ TeloHAECs were plated per well of a 24-well plate in media supplemented with 10 µg/mL of Polybrene Infection / Transfection Reagent (Sigma-Aldrich #TR-1003). TeloHAEC cultures were spinfected with pegRNA lentivirus (generated as in^64^) at 935 x g for 2 hours and 37°C, using low MOI to ensure ≤1 pegRNA integrated per cell. The cultures were then returned to the incubator, and the media was changed after an additional 2 hours. Selection with 15 µg/mL blasticidin was started the next day and continued for an additional 4 days, after which the cultures were transitioned to 3 µg/mL blasticidin for maintenance.

#### Generating and delivering PE7 mRNA

The plasmid template for PE7 (pT7-PE7; Addgene #214813) was PCR amplified as previously described ^188^, followed by a cleanup using the DNA Clean and Concentrator-25 kit (Zymo #D4034) according to the manufacturer’s instructions. In vitro transcription to generate PE7 mRNA from the purified template was performed using the HiScribe T7 mRNA Kit with CleanCap reagent AG (New England Biolabs #E2080S) with 1 µg template per reaction, following the manufacturer’s instructions. The resulting mRNA was purified using the RNA Clean and Concentration-100 kit (Zymo #R1019) and eluted in ∼30 µL water per reaction to achieve a final concentration of ≥1 µg/µL. IVT reactions were performed in bulk to generate high volume, high concentration mRNA.

TeloHAECs were seeded the day prior to nucleofection to reach 70–80% confluence at time of harvest, and media was replaced 1 hour beforehand. Using the Thermo Fisher Neon Transfection System (100 µL; #MPK10096), 3 million cells were nucleofected with 20 µg PE7 mRNA per 100 µL tip, and cells from multiple nucleofections were pooled to achieve the desired cell coverage. Sample preparation for nucleofection followed the manufacturer’s protocol, using the following parameters: 1350 V, 10 ms pulse width, and 3 pulses. Cells were immediately plated at a concentration of 120,000 cells/cm^2^ (3M cells into one T-25) and allowed to recover overnight. Media was changed the following day, and cells were expanded for 7–10 days post-nucleofection before harvest for FlowFISH.

#### PrimeFlow RNA assay and cell sorting

6 x 10^6^ TeloHAECs per RNA-FlowFISH replicate were stained with target probes for *GOSR2* (Thermo Fisher Scientific ‘Type 1’ probe set #VA1-3005956) and *RPL13A* (a positive housekeeping gene; Thermo Fisher Scientific ‘Type 4’ probe set #VA4-13187) using the PrimeFlow RNA Assay Kit (Thermo Fisher Scientific #88-18005-210) and sorted as previously described^64^.

#### Library preparation for amplicon sequencing and data analysis

Genomic DNA extraction and sequencing library preparation were prepared as previously described^64^, with minor modifications. The *GOSR2* enhancer target region was amplified from the genomic DNA of sorted cells using the following primers (Fwd: CACGACGCTCTTCCGATCTCTGGGAGTGGTTTACAGAAACATT, Rev: GACGTGTGCTCTTCCGATCTAACGTACTGAAGTCCTGTGGAG) and thermocycler parameters: 98℃ for 1 minute, 28 cycles of 98℃ for 10 seconds, 66℃ for 20 seconds, and 72℃ for 20 seconds, followed by a final extension at 72℃ for 2 minutes. The completed libraries were sequenced on an Illumina MiSeq, and effects on gene expression were computed using the Variant-EFFECTS snakemake pipeline (v1.0.0; https://github.com/EngreitzLab/Variant-EFFECTS)^64^.

### Generation of PAEC genomic coverage tracks

ChIP-seq datasets for KLF4 and H3K27ac in pulmonary arterial endothelial cells (PAECs) exposed to LSS–like versus control conditions were obtained from GEO accession GSE152894. scATAC-seq profiles from PAECs exposed to LSS versus static culture conditions (ST) were obtained from GSE152890. To generate genomic coverage tracks, input WIG files were processed into bedGraph format using bedtools genomecov -bg -i <SAMPLE>.wig -g hg19.chrom.sizes. The resulting coverage tracks were coordinate-sorted using sort -k1,1 -k2,2n and converted into indexed BigWig format (.bw) for visualization using the UCSC utility bedGraphToBigWig (bedGraphToBigWig <SAMPLE>.bedgraph hg19.chrom.sizes <SAMPLE>.normalized.bw).

### Differential expression and gene expression program enrichment analysis

Gene-level RNA-seq count data comparing PAECs subjected to LSS versus ST were obtained from GEO accession GSE152898, and differential expression analysis was performed using DESeq2 (v1.46.0). Ensembl gene identifiers were trimmed of version decimal suffixes, and mapped to gene symbols using CollapsedGeneBounds.hg38.intGENCODEv43.TSS500bp.bed from https://github.com/EngreitzLab/scE2G. For genes associated with duplicate gene symbols, read counts were aggregated by summing counts across identical symbols prior to differential testing. The processed count matrices were modeled with a single-factor design (∼ condition), and differential expression between LS and ST was assessed using Wald tests. Genes with a Benjamini–Hochberg adjusted *P* value (Padj) < 0.05 were designated as DEGs. To evaluate whether shear-stress-responsive DEGs were significantly enriched within the fetal GEPs, one-sided Fisher’s exact tests (alternative = “greater”) were conducted for each program. Contingency tables (2×2) were constructed classifying genes by GEP membership and DEG status relative to a background universe of all expressed genes evaluated for differential expression (genes passing DESeq2 independent filtering with defined adjusted *P* values). Resulting *P* values across all tested GEPs were corrected for multiple hypothesis testing using the Benjamini–Hochberg procedure.

### Cryopreservation, Cryosectioning, and Imaging of Embryonic Tissue

E11.5 mouse embryos were received previously fixed in 4% paraformaldehyde (PFA). Embryos were washed three times in 1× PBS for 10 min per wash at room temperature with gentle agitation and subsequently cryoprotected in 30% sucrose in PBS overnight at 4°C. Embryos were then embedded in OCT (Optical Cutting Temperature Compound; Fisher HealthCare, Cat. No. 4585) and stored at −80°C until cryosectioning. Embryos were cryosectioned at 10µm thickness. Representative brightfield images of embryonic heart sections were captured using a Zeiss AxioImager M1 microscope (Carl Zeiss, Germany) with the AxioCam MRc at 10x and 40x objectives. Images were processed using Fiji (ImageJ 1.64p).

### Enhancer reporters: Mouse transgenic assay

All animal work was reviewed and approved by the Lawrence Berkeley Laboratory Animal Welfare and Research Committee. Transgenic E11.5 mouse embryos were generated as described previously^189^. Briefly, super-ovulating female FVB mice were mated with FVB males and fertilized embryos were collected from the oviducts. Enhancer sequences were selected to encompass the relevant epigenetic signal and regions of high evolutionary conservation. The selected sequences were synthesized by Twist Biosciences and cloned into the donor plasmid containing minimal Shh promoter, lacZ reporter gene and H11 locus homology arms (Addgene, 139098) using NEBuilder HiFi DNA Assembly Mix (NEB, E2621). The sequence identity of donor plasmids was verified using long-read sequencing (Primordium). Plasmids are available upon request. A mixture of Cas9 protein (Alt-R SpCas9 Nuclease V3, IDT, Cat#1081058, final concentration 20 ng/μL), hybridized sgRNA against H11 locus (Alt-R CRISPR-Cas9 tracrRNA, IDT, cat#1072532 and Alt-R CRISPR-Cas9 locus targeting crRNA, gctgatggaacaggtaacaa, total final concentration 50 ng/μL) and donor plasmid (12.5 ng/μL) was injected into the pronucleus of donor FVB embryos. The efficiency of targeting and the gRNA selection process is described in detail in Osterwalder 2022.^189^

Embryos were cultured in M16 with amino acids at 37 °C, 5% CO2 for 2 hours and implanted into pseudopregnant CD-1 mice. Embryos were collected at E11.5 for lacZ staining as described previously^189^. Briefly, embryos were dissected from the uterine horns, washed in cold PBS, fixed in 4% PFA for 30 min and washed three times in embryo wash buffer (2 mM MgCl2, 0.02% NP-40 and 0.01% deoxycholate in PBS at pH 7.3). They were subsequently stained overnight at room temperature in X-gal stain (4 mM potassium ferricyanide, 4 mM potassium ferrocyanide, 1 mg/mL X-gal and 20 mM Tris pH 7.5 in embryo wash buffer). The stained transgenic embryos were washed three times in PBS and imaged from both sides using a Leica MZ16 microscope and Leica DFC420 digital camera.

PCR using genomic DNA extracted from embryonic sacs digested with DirectPCR Lysis Reagent (Viagen, 301-C) containing Proteinase K (final concentration 6 U/mL) was used to genotype the embryos^73,189^. Specifically, a PCR spanning the 3’ H11 homology arm was used to confirm successful integration of the reporter construct at the H11 locus and a PCR to detect the presence of the bacterial backbone of the reporter plasmid was used to classify the embryo as harboring a “tandem” (concatameric) rather than “single” integration. Only embryos with donor plasmid integration at H11 were used. Embryos with “tandem” integrations tend to have stronger reporter activities than those with “single” integrations, which was taken into consideration when assessing the impact of genetic variant introduction on enhancer activity.

## Data Availability

Interactive exploration and browsing of single-cell RNA-seq, ATAC-seq, gene regulation predictions, and spatial mapping can be found at https://www.engreitzlab.org/heartmap. Processed, de-identified 10x Multiome data are available at https://www.synapse.org/Synapse:syn63997960. Raw sequencing data (10x Multiome) are available via dbGaP (phs003778.v1.p1), in compliance with the study design and consent. Variant-EFFECTS data is available at NCBI Gene Expression Omnibus (GSE334609) and CRISPRi-TAP-seq data is available from the IGVF Data Portal at IGVFDS4617SJVM, respectively. hiPSC-VIC Perturb-seq and single guide MEIS2 knockdown bulk RNA-sequencing data are available at NCBI Gene Expression Omnibus (GSE349107 and GSE349277, respectively).

## Code Availability

The data analysis code is available at: https://github.com/rosaxma/Heart-2024

## Data Availability

Interactive exploration and browsing of single-cell RNA-seq, ATAC-seq, gene regulation predictions, and spatial mapping can be found at https://www.engreitzlab.org/heartmap. Processed, de-identified data are available at https://www.synapse.org/Synapse:syn63997960. Raw sequencing data (10x Multiome) are available via dbGaP (phs003778.v1.p1), in compliance with the study design and consent. Variant-EFFECTS data is available at NCBI Gene Expression Omnibus (GSE334609) and CRISPRi-TAP-seq data is available from the IGVF Data Portal at IGVFDS4617SJVM, respectively. hiPSC-VIC Perturb-seq and single guide MEIS2 knockdown bulk RNA-sequencing data are available at NCBI Gene Expression Omnibus (GSE349107 and GSE349277, respectively). Data analysis code is available at https://github.com/rosaxma/Heart-2024.

https://www.engreitzlab.org/heartmap

https://www.ncbi.nlm.nih.gov/projects/gap/cgi-bin/study.cgi?study_id=phs003778.v1.p1

https://www.ncbi.nlm.nih.gov/geo/query/acc.cgi?acc=GSE334609

https://data.igvf.org/analysis-sets/IGVFDS4617SJVM/

https://www.ncbi.nlm.nih.gov/geo/query/acc.cgi?acc=GSE349107

https://www.ncbi.nlm.nih.gov/geo/query/acc.cgi?acc=GSE349277

https://github.com/rosaxma/Heart-2024

## Acknowledgements

We thank Salil Deshpande, Laksshman Sundaram, and members of the Engreitz Lab and BASE Initiative for helpful discussions. We thank Mo Ameen, Simon Miao, Betty Liu, and William Greenleaf for technical advice on single-cell tissue dissociation methods. We thank James Galante for assistance with analysis of TAP-seq data. We thank Stanford University and the Stanford Research Computing Center for providing computational resources and support for the Sherlock high-performance compute cluster. We thank the Stanford Genomics Service Center for generating sequencing data used in this publication.

## Author Contributions

X.R.M. and J.M.E. led analysis to create the atlas and interpret genetic variants.

S.D.C., X.R.M., W.R.G., and J.M.E. led fetal tissue sample acquisition and multiome data collection. S.D.C., B.D.R.L., and I.A.G. collected fetal tissue.

S.D.C., X.R.M., D.A., C.J.M., O.M.R., M.G., J.M.E., and W.R.G. developed experimental methods for multiome data collection.

S.D.C., L.D., S.T., K.D., and W.R.G. conducted multiome experiments.

X.R.M. and H.Y.K. developed and applied methods to analyze multiome data. X.R.M., C.A.G., M.R., T.Q., J.M.E., and W.R.G. annotated cell types and states. Y.L., S.C., X.R.M., and X.Q. analyzed spatial transcriptomics data.

D.B., S.Kundu, and A.K. trained ChromBPNet models to interpret variants and identify TF motif instances. X.R.M., M.U.S., W.Q., R.A., and J.M.E. developed and applied the scE2G model.

D.A., M.S.C., C.V.M., S.V., C.J.M., and J.M.E. designed, conducted, and analyzed CRISPRi-TAP-seq experiments.

X.R.M., M.S.C., W.R.G., and J.M.E. designed validation experiments in hiPSC-VICs.

M.S.C., R.P., C.B., Y.C.L., S.D.C., T.Z., Y.A.M., J.V., T.N., W.R.G., and J.M.E. conducted and analyzed experiments in hiPSC-VICs.

M.T.M., S.F.X., X.C., and J.M.E. designed, conducted, and analyzed Variant-EFFECTS experiments. J.R.M., D.A., and M.R. analyzed endothelial cell flow data.

J.P.P., J.R.P., S.Kany., S.Z., J.D., A.P., J.O., and S.M.D. contributed genome-wide association study results.

R.C. and J.P.P. conducted statistical fine-mapping and interpreted analysis of GWAS variants.

S.C. and X.Q. analyzed cell-cell interactions.

M.K., N.S., X.R.M., Z.A., K.R., L.A.P., A.V., W.R.G., and J.M.E. designed, conducted, and interpreted enhancer reporter experiments.

S.D.C., J.M.E., and I.A.G. managed regulatory compliance.

W.R.G. and J.M.E. obtained funding and supervised the study.

X.R.M., S.D.C., M.S.C., W.R.G., and J.M.E. wrote the manuscript with input from all authors.

## Supplementary Notes

### Note S1. Regulatory wiring of cardiac conduction cells

The high cell number and ATAC sequencing quality allowed us to examine, for the first time, the regulatory landscape of rare but crucial cell types in the developing human heart. For example, our atlas includes 8,906 cardiac conduction cells—specialized cell types that coordinate the heart’s rhythmic contractions—and 13,103 transitional cells, which serve as a cellular bridge between conduction cells and the surrounding working myocardium, previously discovered in the murine heart^41^ (**Figs. S8A**, **S11**). The cardiac conduction cells included cell populations from the four major substructures of the conduction system: SAN (1,211 cells), AVN (2,795 cells), His bundle (939 cells), and Purkinje fibers (PF1 and PF2, 1,830 cells and 2,130 cells, respectively) (total cell frequencies: 0.13-0.38%; **Figs. S8A, S11B**). Each of these populations was distinguished by established markers, such as *ISL1* and *SHOX2* for the SAN and *HS3ST3A1*^49,190^ for the His bundle (**Fig. S8B,C**).

We examined our maps to identify enhancers and TFs that distinguish these CCS cell types. As expected, their global profiles of enhancer-gene regulatory interactions were distinct from other cell types (*e.g.*, 26% overlap for the SAN vs. capillary endothelial cells) (**Fig. S8D**). Among the CCS cell types, there was increased overlap in predicted enhancers within the nodes (SAN vs. AVN: 59% overlap) as compared to the ventricular conduction system (VCS) components (SAN vs. His bundle or PFs: 43-48%), consistent with distinct gene expression profiles and physiologic functions of these cell types (**Fig. S8D**). Toward understanding this molecular wiring, we examined the enhancers that regulate *ISL1*, a key TF in the development and function of the SAN (**Fig. S8E**). We identified 8 predicted enhancers for *ISL1* in SAN cells, including the one known enhancer that has been previously characterized^71^. Motif instances of other TF families involved in SAN development are predicted to strongly influence the accessibility of the *ISL1* enhancers. These included several GATA motifs with stronger predicted contributions to chromatin accessibility in SAN cells than other conduction cell types (**Fig. S8F**). Furthermore, *GATA6* and *ISL1* are both prominent members and predicted regulators of the gene program for SAN identity (**Figs. S12A, S13**), suggesting a potential regulatory cascade where GATA6 drives both *ISL1* up-regulation and the broader SAN-specific gene program.

Examining the gene programs that defined each CCS cell type, we unexpectedly found many genes previously known to be involved in neural adhesion and axon development (**Figs. S8G, S13**). Neuronal cell adhesion molecules play a crucial role in mediating cell-cell interaction (CCI) to achieve cell patterning and discrete cellular niches within the developing brain. Their involvement in the developing CCS, however, is not understood. By examining gene expression patterns and exploring CCI via analysis of ligand-receptor pairs with CellChat^156^ (**Table S9**), we indeed found multiple ligand-receptor genes classically related to CNS development (*SLIT/ROBO*, *NRXN/NLGN*, *EPH/EPHRIN*) within the CCS and surrounding cell types (**Figs. S8H-M, S12B, S13**). For example, ligand *SLIT2* was expressed in PF and SAN cells, and its receptors (*ROBO1* and *ROBO2*) were expressed by surrounding cell types, including transitional SAN cells and fibroblasts (for the SAN) and VCMs (for PFs) (**Figs. S8H,K, S12B, S13A,E**). Receptors *EPHA4* and *EPHA7* were expressed in the His bundle and PF cells (**Figs. S8I, S12B, S13D,E**), and their ligand *EFNA5* was expressed in surrounding VCMs as well as transitional cells (**Fig. S8L**). Ligands NRXN1 and NRXN3 were found in AVN (**Figs. S8J, S12B, S13B,C**), and their receptors, including LRRTM3 and/or NLGN1, were enriched within the surrounding cells of the ACM and VCM (**Fig. S8M**). Consistent with a possible role for these factors in CCS development, variants associated with *SLIT2* have been associated with abnormal QRS duration^191^, suggestive of an abnormal VCS, and *ROBO1* variants have also been associated with abnormal EKG findings^192^. Further, systemic knockdown of *Epha4* in rats resulted in, among other phenotypes, QRS prolongation^193^. These expression patterns showed similarities but also notable differences with recent single-cell studies in the mouse fetal heart^190^ and human adult CCS^194^, and a study also observed upregulation of *SLIT2* and *NRXN3* in SAN and AVN, respectively, in the human fetal heart^49^. The established role of these cell adhesion molecules in neuronal differentiation and establishing CCI hints at a similar role for the normal differentiation of the VCS and/or patterning within the developing trabecular myocardium. Together, this provides the first regulatory map of these clinically important CCS cells and illustrates the insights possible with this deep atlas.

### Note S2. Enrichment of congenital heart disease risk genes in cardiac fibroblast progenitors

Cardiac fibroblasts are diverse mesenchymal cell populations found throughout the heart including in the septa, myocardium, and valve annuli^49,195^. The cell type in our atlas with the strongest enrichment of high-confidence CHD genes was cardiac fibroblast progenitors. This population corresponds to an epicardial-derived transitional state, characterized by high expression of *WT1 and TBX18* compared to other mature fibroblasts and fibroblast-like lineages in the atlas (**Fig. 2B, Fig. S5C**)^4^. This transitional status was independently cross-validated by the projection of the spatially resolved reference fetal heart atlas^48^: notably, ∼70% of the reference cells mapping to this cluster were annotated as epicardial-derived progenitor cells (EPDCs), while the remainder aligned with mature atrial fibroblast and valvular interstitial lineages (**Fig. S9; Table S3**). Strikingly, these progenitor cells were enriched for genes associated with ASD, valve defects, and malformations of the OFT (8-10-fold enrichment, P_Bonferroni ≤ 2 × 10_−4_; **Fig. 2A; Table S12**).

Of the 16 CHD genes highly expressed in cardiac fibroblast progenitors, 4 have been shown to lead to heart defects in animal models using conditional knockouts in the epicardial lineage (**Fig. 2E, Table S13, Note S2**). For example, *MEIS2* mutations have been observed in patients with ASD or VSD^196,197^, coarctation of the aorta^198^, Tetralogy of Fallot, hypoplastic right ventricle, and Ebstein’s anomaly^199^. In mice, *Meis2* appears to play roles in both epicardial-derived fibroblasts and VICs. In the epicardial lineage, double knockout of *Meis1/2* (using Cre recombinase driven by *Wt1* promoter) leads to a decrease in the proportion of epicardial-derived cardiac fibroblasts, and leads to VSD, malalignment of the great vessels, asymmetric septation of the OFT, and defects in coronary artery development^200^. Relevant to VICs, knockout of *Meis2* in the cardiac neural crest lineage (using *Sox9*-Cre) leads to severe defects in the aortic and pulmonary valves^201^. In our heart atlas, *MEIS2* is a member of Fetal VIC program 27 (see above) and also of FB program 6 (“Migration of epicardial-derived cardiac fibroblasts”), a gene program expressed in cardiac fibroblast progenitors that includes many genes involved in cell migration (*e.g.*, *SPOCK1*, *FAT4*, *SEMA6D*, *PLXNA4*, *TIAM1*, and *ENAH*). FB program 6 also includes 13 additional high-confidence CHD genes (odds ratio=6) (**Fig. S17**), including *PBX1*, which encodes a known interaction partner of MEIS2^202^, and six genes (*TBX18, CACNA1A*, *GATA6*, *GATA4*, *SMAD6*, *ZEB2*) that carry a significant burden of heterozygous *de novo* or rare transmitted damaging variants in a large-scale CHD case-control cohort^7^.

### Note S3. Evidence linking CHD genes to functions in VICs and cardiac fibroblasts

We found that genes previously associated with multiple subtypes of CHD were enriched for high expression in VICs and cardiac fibroblast progenitors (**Fig. 2**). In total, 27 genes were highly expressed in either VICs or cardiac fibroblasts. Many of them have been studied using mouse knockouts, including via conditional knockouts in relevant cell lineages (endocardial lineages, neural crest, and epicardial for VICs^203^); and epicardial lineage for cardiac fibroblasts^195^. This supports the notion that CHD genes can have direct effects on these cell types to lead to heart defects. We describe these previous studies below, and summarize in **Table S13**.

- ***BMPR2***: *BMPR2* (bone morphogenetic protein receptor type 2) plays an essential role in heart valve development, particularly through its involvement in endocardium and regulation of valvulogenesis. Conditional knockout studies using Cre-loxP lineage tracing have revealed tissue-specific functions of *BMPR2* during cardiogenesis. Specifically, conditional deletion of *Bmpr2* with *Tie2*-Cre, which targets endocardial cells and derived mesenchymal cells, leads to AV cushion defects, including membranous VSDs and thickened valve leaflets, but does not block EndMT^204^. In addition to its function in the endocardial lineage, neural crest lineage deletion of *Bmpr2* (*Wnt1*-Cre) leads to abnormal positioning of the aorta^204^. In contrast, myocardial lineage knockout of *Bmpr2* did not affect heart development^204^. In humans, mutations in *BMPR2* are observed in patients with pulmonary arterial hypertension (PAH)^204,205^, ASD, VSD, PDA, and malformation of the OFT^205^.
- ***FGFR2***: *FGFR2* (Fibroblast Growth Factor Receptor 2) is important for cardiac development, particularly in the formation and alignment of the cardiac OFT. In humans, mutations in *FGFR2* are associated with CHD, including VSD, double outlet right ventricle (DORV), and hypoplastic ventricles^206–209^. Conditional knockout studies in mice using the Cre-loxP system have shown tissue-specific roles of *FGFR2* during cardiogenesis. *Fgfr2* deletion in mesodermal cells of the SHF using *Nkx2.5*-Cre, which targets SHF-derived OFT progenitors, results in severe OFT defects, such as overriding aorta, DORV, and persistent truncus arteriosus (PTA). These defects show *FGFR2*’s role in OFT morphogenesis^209^. Notably, dual inactivation of *Fgfr1* and *Fgfr2* in the SHF lineage further confirms *Fgfr2*’s function in regulating EMT and neural crest cell recruitment to the OFT cushions^209^. Germline knockout of *Fgfr2b* resulted in thin-walled myocardium and smaller hearts at embryonic day 17.5 (E17.5), indicating that FGFR2b signaling is essential for proper heart development^210^. In a conditional knockout study, *Fgfr1* and *Fgfr2* were inactivated in the epicardium and epicardial-derived cells of mice using *Wt1*-Cre. These mice exhibited reduced epicardial *Fgfr2* expression, decreased myocardial proliferation, and thinner myocardial walls^210^.
- ***GATA4***: GATA4 (GATA Binding Protein 4) is a TF essential for heart development, particularly in heart valve formation and maturation, through its involvement in EndMT and mesenchyme formation within AV cushions. Conditional knockout studies using *Tie2*-Cre lineage tracing, which targets endocardial cells and endocardial cell-derived VICs, show that *Gata4* deletion disrupts AV cushion development^211^. This disruption results in hypoplastic valves with few VICs, leading to defective AV septation, hypocellular cushions, and malformed valves. Specifically, *Gata4* inactivation in the endocardial lineage impairs EndMT, reducing mesenchymal cell formation and resulting in unseptated ventricular inlets and incomplete cushion remodeling^211^. In our atlas, *GATA4* is expressed across multiple cell types, including VICs, where it is predicted to regulate gene programs involving other CHD-related genes, such as *LTBP2*, *MEIS2*, and *TBX20*. Human *GATA4* mutations are associated with various CHDs, including AVSD, pulmonic stenosis, and hypoplastic right ventricle, as well as ASDs and VSDs and valve malformations^211,212^.
- ***GATA6***: GATA6 (GATA Binding Protein 6) is a TF involved in heart development, especially in valve formation and septation. *GATA6* mutations in humans are also associated with PTA, ASDs, and DORV, which are linked to defects in OFT development and valve morphogenesis^24,103^. *Wnt1*-Cre-mediated conditional *Gata6* knockout in neural crest-derived cells in mice results in CHD, such as PTA and interrupted aortic arch. These defects arise from disrupted morphogenetic patterning and impaired migration of cardiac neural crest cells, accompanied by downregulation of key signaling molecules like semaphorin 3C essential for cardiac neural crest cell function^103^. Additionally, deletion of *Gata6* with an *Isl1*-Cre mouse model targeting SHF progenitors leads to defects in the AV canal, including valve malformations and ASD, by impacting the proliferation and differentiation of SHF-derived cells^213^. Epicardial-specific knockout of *Gata4/Gata6* causes a thin compact myocardium and coronary artery formation defects, which are present in some patients with Tetralogy of Fallot. Furthermore, *GATA6* is highly expressed in epicardial-derived fibroblasts, regulating gene programs vital for myocardial and coronary vessel development^212,214^.
- ***GLI3***: Disruption in GLI3’s (GLI Family Zinc Finger 3) activity, often linked to ciliopathies, is associated with defects in heart valve development, such as myxomatous mitral valve and BAV^215,216^. These conditions are marked by abnormal extracellular matrix composition and VIC differentiation^215^. Primary cilia, which are present on VICs during early development, play a mechanosensory and signaling role^216^. Ablation of genes critical for ciliogenesis, like *Ift88*, results in loss of cilia and subsequent valve enlargement, ECM disruption, and fibroblast-like cell differentiation^216^. Genetic models using Cre drivers like *Nfatc1*-Cre (targeting endocardial cells and VICs) to knock out cilia-related genes show a higher prevalence of CHD, including BAV and myxomatous mitral valve, with affected mice displaying altered valve morphology, increased ECM components like collagen and proteoglycans, and decreased cell density^215,216^.
- ***GPC3***: To our knowledge, *GPC3* (Glypican-3) has not yet been studied in conditional knockout models within endocardial, neural crest and epicardial lineages. In humans, mutations in *GPC3* result in Simpson-Golabi-Behmel syndrome (SGBS), an X-linked disorder that presents a range of cardiac defects. These include ASDs, VSDs, PDA, patent foramen ovale, BAV, dysplastic tricuspid and pulmonary valves, hypoplastic left pulmonary artery, and, in some cases, arrhythmias like supraventricular tachycardia and conduction defects^217^. In *Gpc3* knockout mouse models, the loss of *Gpc3* function leads to similar structural defects, including VSD, common AV canal, DORV, and coronary artery fistulas^218^. Valve abnormalities observed in *Gpc3*-deficient mice involve AV canal defects, with bridging valve leaflets and fibrous discontinuity between the aortic and mitral valves^218^. In our atlas, *GPC3* is expressed primarily in mesenchymal cells, with high expression in epicardial-derived cardiac fibroblasts, OFT fibroblasts, and VICs (**Fig. S15**).
- ***INVS***: In mouse models, global *INVS* (inversin) mutation (insertional mutagenesis) leads to a range of CHD, including pulmonary infundibular stenosis, VSDs, and abnormalities in the right ventricular OFT^219^. To our knowledge, this gene has not been studied using conditional knockouts in the endocardial, neural crest, or epicardial lineage. *INVS* (also known as *NPHP2*) mutations in humans are associated with VSDs^220,221^, mitral insufficiency, and pulmonary valve stenosis^221^.
- ***LTBP2***: In *Ltbp2* (Latent-transforming growth factor beta-binding protein 2) knockout mouse models, absence of Ltbp2 results in a high prevalence of mitral valve prolapse^83^. To our knowledge, this gene has not been studied using conditional knockouts in the endocardial, neural crest lineage, or epicardial. In humans, mutations in *LTBP2* have been linked to CHD such as mitral valve prolapse and polyvalvular heart dysplasia^80,83^.
- ***MEIS2***: MEIS2 (Meis homeobox 2) is a TF involved in heart and craniofacial development, particularly influencing the morphogenesis of neural crest-derived tissues. In humans, *MEIS2* haploinsufficiency has been associated with a range of CHD, including VSD, ASD, coarctation of the aorta, Tetralogy of Fallot, hypoplastic right ventricle, and Ebstein’s anomaly^199^. Conditional knockout studies in mice have shown the essential roles of MEIS2 across different cardiac lineages. Neural crest-specific deletion of *Meis2* (*AP2α*-Cre) leads to CHD such as PTA, where the aorta and pulmonary arteries fail to separate, as well as valve malformations including defects in the aortic, pulmonary, tricuspid, and mitral valves. These defects are associated with embryonic lethality around E14 due to hemorrhaging and impaired cardiac neural crest migration^201^. Additionally, epicardial-specific deletion of both *Meis2* and its paralog, *Meis1*, using *Wt1*-Cre has been shown to reduce the proportion of epicardial-derived cardiac fibroblasts, leading to VSDs, misalignment of the great vessels, asymmetric septation of the OFT, and abnormalities in coronary artery development^201^.
- ***NF1***: *NF1* (neurofibromatosis type 1) plays a role in heart development, in particular by regulating EndMT during valvulogenesis^222^. In humans, mutations in *NF1* are associated with CHD, including pulmonary valve stenosis, ASD, and VSD. Conditional knockout studies in mice using Cre-loxP lineage tracing have shown tissue-specific functions of *NF1* in cardiogenesis: epicardial-specific deletion of *Nf1* using Wt1CreERT2 results in accelerated EMT, leading to an increased number of epicardial-derived cells, such as fibroblasts, migrating into the myocardium and contributing to defects in the ventricular septum and AV valves^222^. Additionally, endothelial cell-specific knockouts show thickened endocardial cushions due to enhanced EMT, which correlates with CHD like VSDs and OFT abnormalities^222,223^.
- ***NOTCH2***: *NOTCH2* (Neurogenic locus notch homolog protein 2) plays a role in heart development, particularly through its involvement in neural crest-derived mesenchymal cells and the morphogenesis of the OFT. In humans, mutations in *NOTCH2* are associated with CHD such as peripheral pulmonary stenosis, aortic coarctation, and complex cardiac anomalies linked to Alagille syndrome^224^. Conditional knockout studies in mice using Cre-loxP lineage tracing have uncovered tissue-specific functions of *NOTCH2* during cardiogenesis. Specifically, deletion of *Notch2* with *Pax3*-Cre, which targets cardiac neural crest cells, results in narrowed aorta and pulmonary arteries, mimicking defects seen in Alagille syndrome. Similarly, *Tag1*-Cre-mediated inactivation of *Notch2* also leads to OFT defects, characterized by smaller vessel diameters due to decreased proliferation of SMCs derived from mesenchymal cells. In contrast, deletion of *Notch2* in secondary heart field derivatives using *Mef2c*-AHF-Cre did not cause OFT abnormalities, suggesting a neural crest lineage-specific function for *Notch2*^224^.
- ***PBX1***: PBX1 (PBX Homeobox 1) is a TALE-class homeodomain TF that contributes to the septation of the OFT, endocardial development, VSDs, conotruncal malformations, semilunar valves, and patterning of the great arteries^202,225^. In *Pbx1*-null mouse models, severe cardiovascular anomalies are observed, including PTA, VSDs, and abnormal great artery patterning. These defects primarily result from the failure of OFT septation, which leads to the formation of a single arterial trunk, as well as impaired valve development, where a single valve forms instead of distinct semilunar valves. *Pbx1* mutants also display additional vascular abnormalities, such as cervical aortic arch and aberrant subclavian artery origins, indicating a broader role in arterial patterning. Lineage tracing studies using a *Wnt1*-Cre mouse model, which targets neural crest cells, shows that cardiac neural crest cells can still migrate to the OFT in *Pbx1*-null embryos^202,225^.
- ***PRKD1***: In humans, *PRKD1* (Protein kinase D1) mutations are linked to CHD such as AVSD, BAV, pulmonary stenosis, and coarctation of the aorta^226^. In a *Prkd1* transgenic model with a CRISPR/Cas9-mediated deletion of exon 2, homozygous mice exhibited severe CHD, such as AVSD, BAV, dysplastic pulmonary valves, and VSDs^226^.
- ***RBFOX2***: RBFOX2 (RNA Binding Fox-1 Homolog 2) is an RNA-binding protein that regulates alternative splicing networks essential for cardiac development, particularly in the formation of heart chambers and valves in humans. Conditional deletion of *Rbfox2* with *Nkx2.5*-Cre, which affects early cardiac progenitors, disrupts chamber heart structure and valve development^227^. *RBFOX2* loss impairs cell-ECM adhesion and EndMT, processes critical for valve formation and the proliferation of endocardial and myocardial cells. The model also suggests that *RBFOX2* is required across multiple cardiac cell lineages, as demonstrated by its broad expression in myocardial, endocardial, and contractile cells, which collectively contribute to normal cardiovascular morphogenesis^227^.
- ***SETD5***: *SETD5* (SET Domain Containing 5) haploinsufficiency is linked to CHD, including VSDs and OFT abnormalities in humans^228^. Deletion of *Setd5* using the *Mesp1*-Cre driver, which targets early mesodermal progenitors including the first and second heart fields, results in severe cardiac malformations, such as shortened OFTs, abnormal chamber ballooning, and embryonic lethality by E12.5^228^.
- ***SMAD6***: During valve development, *SMAD6* (SMAD Family Member 6) is expressed in endocardial and mesenchymal cells, contributing to regulated mesenchymal cell proliferation. In humans, disruptions in SMAD6 function have been associated with CHD, including septation abnormalities and valve malformations. In *Smad6*-deficient knockout mouse models, a lack of SMAD6 leads to hyperplasia of the cardiac valves, marked by excessive mesenchymal cell proliferation and an enhanced EndMT. Additionally, mutants show OFT defects, often resulting in a narrowed ascending aorta or an enlarged pulmonary trunk^229^.
- ***TBX20***: *TBX20* (T-Box Transcription Factor 20) plays a role in cardiac valve development, exerting lineage-specific influences that contribute to CHD across endocardial, myocardial, and VIC lineages. *TBX20* mutations in humans are linked with a wide array of congenital heart anomalies, such as ASD, VSD, tetralogy of Fallot, total anomalous pulmonary venous connection, and cardiomyopathies with defective valvulogenesis^230–232^. In murine models, conditional knockout studies illustrate cell-specific roles of *Tbx20* during valvulogenesis. *Nfatc1*-Cre-mediated knockout, targeting endocardial cells and their mesenchymal derivatives, leads to valve elongation defects, shortened leaflets, and structural abnormalities in all four cardiac valves without inhibiting EMT initiation^231^. This deletion disrupts Wnt/β-catenin signaling, resulting in reduced cellular proliferation and impaired extracellular matrix organization within valve cushions. In contrast, myocardial lineage deletion of *Tbx20*, important for early cardiac patterning, shows no effect on valve formation^231^. Partial knockdown of *Tbx20* in mice also results in severe valve developmental defects^233^. Furthermore, *TBX20*’s role may extend into regulatory functions within VICs, where it could influence factors like Fetal VIC program 27.
- ***TGFBR1***: *TGFBR1* (transforming growth factor-beta (TGF-β) receptor type 1) plays a role in myocardial wall formation and cardiac morphogenesis^234^. In humans, *TGFBR1* mutations are linked to VSDs, Marfan syndrome, and Loeys–Dietz syndrome^234^. Conditional deletion of *Tgfbr1* with *Tie2*-Cre, targeting endocardial cells, results in hypoplastic AV cushions, impaired EMT, and thin myocardium, indicating its role in EndMT and myocardial proliferation^235^. Epicardial-specific knockout using *Gata5*-Cre leads to detached epicardium, reduced myocardial proliferation, and defective smooth muscle formation around coronary vessels, showing its importance in epicardial-mesenchymal transformation^235^. Studies using myocardial-specific inactivation of *Tgfbr1* with *cTnt*-Cre mice demonstrate that while most embryos do not show overt cardiac defects due to compensation by other TGF-β receptor genes, constitutively active *TGFBR1* expression can arrest heart development at the looping stage and lead to ventricular hypoplasia^234^. In neural crest cells, targeted deletion of *Tgfbr1* with a *Wnt1*-Cre model results in severe aortic arch malformations and PTA, showing its role in OFT development^234^. Furthermore, *Tgfbr1* expression in VICs and its regulation by miRNAs show its importance in maintaining proper signaling balance during heart development to prevent non-compaction and spongy myocardial wall defects^234^.
- ***ZFPM2***: *ZFPM2* (Zinc finger protein), also known as *FOG2*, regulates coronary vessel formation and morphogenesis. In humans, mutations or disruptions in *ZFPM2* have been implicated in septal defects and OFT malformations^236^. Knocking out *Zfpm2* in mice leads to multiple CHD, including ventricular myocardium thinning, ASD, and features of tetralogy of Fallot, such as VSDs, overriding aorta, and subpulmonic stenosis^236,237^. The absence of ZFPM2 disrupts epithelial-to-mesenchymal transition in epicardium-derived cells, which affects the development of coronary vasculature, fibroblasts, and other mesenchymal derivatives. Rescue experiments indicate that ZFPM2 function in the myocardium is essential for inducing signals required for vascular formation^236,237^.

### Note S4. Additional examples of variants active in cell-type-specific enhancers

Variant rs6702619, near *PALMD*, is associated with a subtype of CHD (BAV), as well as with calcific aortic valve stenosis and quantitative traits related to the aorta and aortic valve in adults (**Fig. S22C**)^37,90,173,238^. Previous studies have shown that rs6702619 is an eQTL for *PALMD* in the adult aortic valve^239^, overlaps an enhancer in adult VICs, and affects a binding site for NFATC2^240^. The target gene, *PALMD*, has been shown to regulate EndMT and VIC remodeling^240,241^. In our map, rs6702619 overlaps an enhancer predicted to regulate *PALMD* not only in VIC but also valvular endocardial cells and aortic endothelial cells (**Fig. S22D**). ChromBPNet predicts that the disease-associated allele (G) reduces chromatin accessibility at the enhancer by 43%-68% in valvular cells, with the strongest reduction in endocardial cells on the IF side of the valves (**Fig. S22E**), consistent with the direction of effect of the adult eQTL and NFATC2 binding^239,240^. In fetal endocardial cells, *PALMD* is part of a WNT signaling in valvular endocardial cells gene program (“Endocardial Program 29”) that is regulated by KLF2/4 and includes other factors important in valve development **(Fig. S21, Tables S7, 8**). Among all gene programs, Endocardial program 29 exhibited the highest enrichment of genes downregulated when *PALMD* was knocked down in human umbilical vein endothelial cells (HUVECs), both under the static condition (15 genes, 16.7-fold enrichment, *P* = 1.5 x 10^-10^, Bonferroni-corrected) and the flow condition (21 genes, 16.1-fold enrichment, *P* = 8.8 x 10^-15^, Bonferroni-corrected)^242^. This indicates that Endocardial program 29 includes genes downstream of the effects of PALMD. Together, these data show that rs6702619 overlaps an enhancer active in fetal valve cells and that *PALMD* is co-regulated with genes involved in valve development, providing a molecular explanation for its association with congenital valve defects.

At the *MTSS1* locus, the variants rs35006907, rs34866937, and rs12541595 are in strong LD (*R*^2^ ≥ 0.978). These variants are associated with traits related to LV function, including decreased end-systolic and increased ejection fraction^36^ (**Fig. S22F**). The alternative haplotype is also associated with lower expression of *MTSS1* in the LV (GTEx)^243^. In our atlas, rs35006907 and rs34866937—but not rs12541595—overlap a single enhancer predicted to regulate *MTSS1* in all VCMs and a small subset of ACMs (**Fig. S22G**). ChromBPNet predicts that the alternative allele (A) at rs35006907 reduces chromatin accessibility of the enhancer by 10-30% in VCMs, with the highest reduction in a population of trabecular cardiomyocytes, by altering a motif instance that matches USF1 and MITF (**Fig. S22H**). *Mtss1* knockout in mice is embryonic lethal and leads to decreased LV end-diastolic and end-systolic dimension^244^, and knockout in medaka leads to a marked reduction in trabeculation^30^. A recent parallel study perturbed *MTSS1* in hiPSC-cardiomyocytes and found that it affects the production of sarcomeres, confirming a cell-autonomous role in cardiomyocytes^245^. Our results indicate that, because this enhancer is active early in fetal development, the effects of this variant on many myocardial traits could have developmental origins.

To validate the cell type specificity of these enhancers, we tested the expression patterns of these 2 enhancers in E11.5 mouse embryos using a transgenic enhancer reporter assay (**Fig. S22I; Table S17**). Both enhancers had spatially restricted activities that were consistent with the predicted cell types of action above (**Fig. S22J, K, Fig. S23**). The *PALMD* enhancer showed specific activity in the OFT, consistent with the scE2G-predicted activity in aortic and valvular endocardial cells in our human atlas (**Fig. S22J, Fig. S23**). The *MTSS1* enhancer showed specific activity in the ventricular septum (**Fig. S22K, Fig. S23F**). We further tested the effects of *PALMD* variants on enhancer activity, and found that introducing the alternative allele for rs6702619 and tightly linked rs11166276 led to undetectable enhancer activity, consistent with its strong predicted effect on accessibility (**Fig. S23**). Similarly, consistent with our ChromBPNet predictions, the risk allele at rs35006907 substantially reduced *MTSS1* enhancer activity, whereas the linked variant rs34866937 had no detectable effect. These results provide the first in vivo functional validation of the causal variant at the *MTSS1* locus^244,246^. The direction of effect of the *MTSS1* variant is consistent with its association with decreased end-systolic volume and increased ejection fraction. This is because MTSS1 operates as a negative regulator of sarcomere assembly and contractility^244,245^. The risk variant may decrease enhancer activity, reduce *MTSS1* expression, and therefore increase contractility and ejection fraction.

Together, these analyses suggest that noncoding variants can influence risk for CHD and other structural traits by regulating gene expression in specific cell types and subanatomic locations in the developing heart. Including the examples presented here, annotations of 750 GWAS signals for cardiac diseases and traits will provide a rich resource for future study (**Table S16**).

## Supplementary Figures

**Fig. S1.**
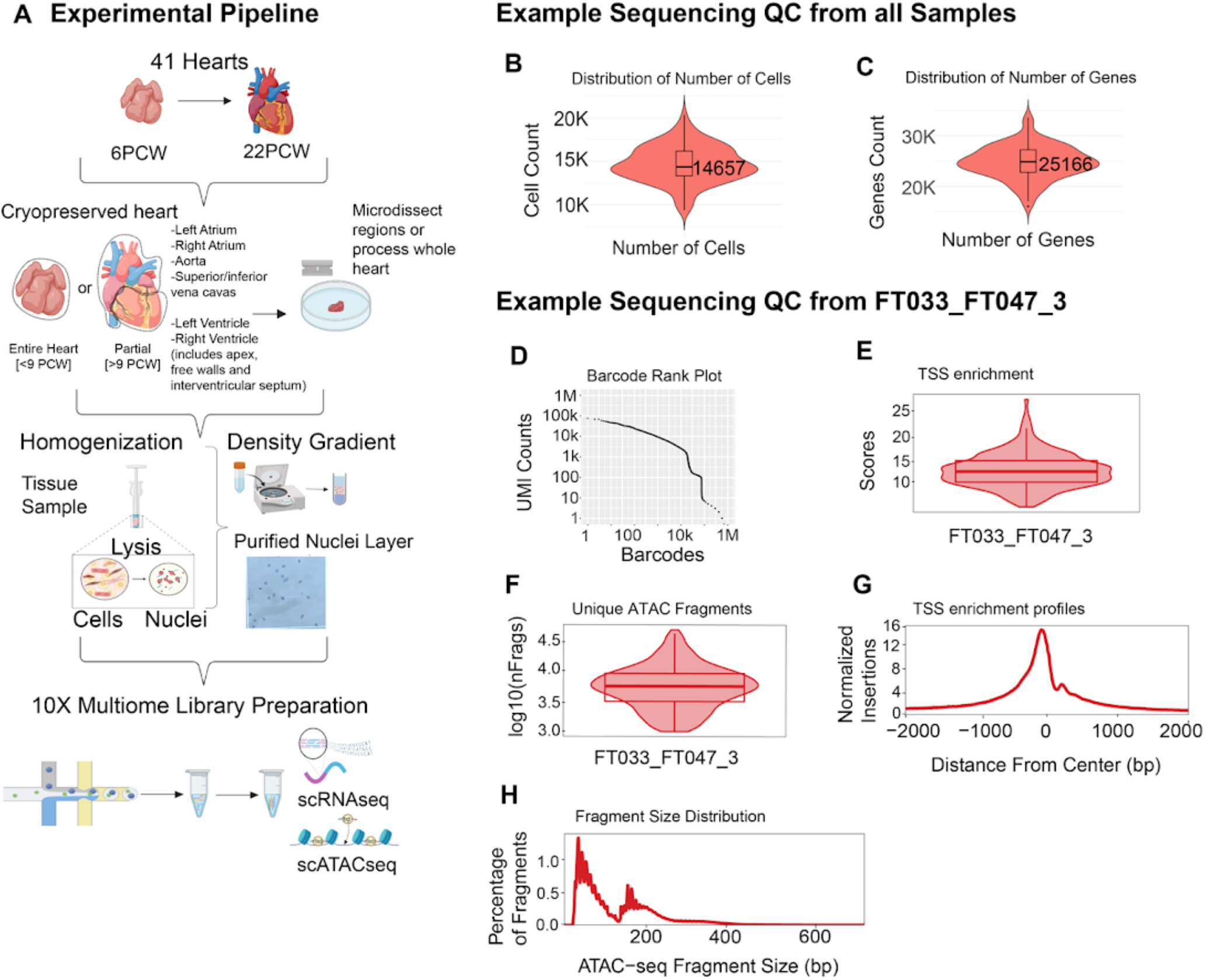
Experimental pipeline and quality control metrics for single-cell 10x Multiome data. **A.** Experimental pipeline: Illustration of the workflow used to process 41 human fetal hearts ranging from 6 to 22 PCW. After cryopreservation, specific heart tissue regions were dissected, followed by homogenization and lysis to release nuclei. A density gradient was applied to purify nuclei. The nuclei were used for 10x Multiome library preparation, generating single-cell RNA sequencing (scRNA-seq) and single-cell ATAC sequencing (scATAC-seq) libraries^247^. **B.** Number of cells: Violin plot depicting the distribution of cell counts per 10x lane. **C.** Number of genes: Violin plot showing the total number of genes detected per 10x lane. **D.** Barcode rank plot: UMI counts across barcodes in a representative sample (FT033_FT047_3). **E.** TSS enrichment scores: Violin plot illustrating the enrichment scores for transcription start site (TSS) regions in the sample FT033_FT047_3. **F.** Unique ATAC fragments: Violin plot displaying the distribution of unique ATAC fragment counts per cell on a log10 scale for the sample FT033_FT047_3. **G.** TSS enrichment profiles: Line graph showing the normalized insertion profile centered around transcription start sites for sample FT033_FT047_3. **H.** scATAC-seq fragment size distribution: Histogram representing the distribution of scATAC-seq fragment sizes for sample FT033_FT047_3.

**Fig. S2:**
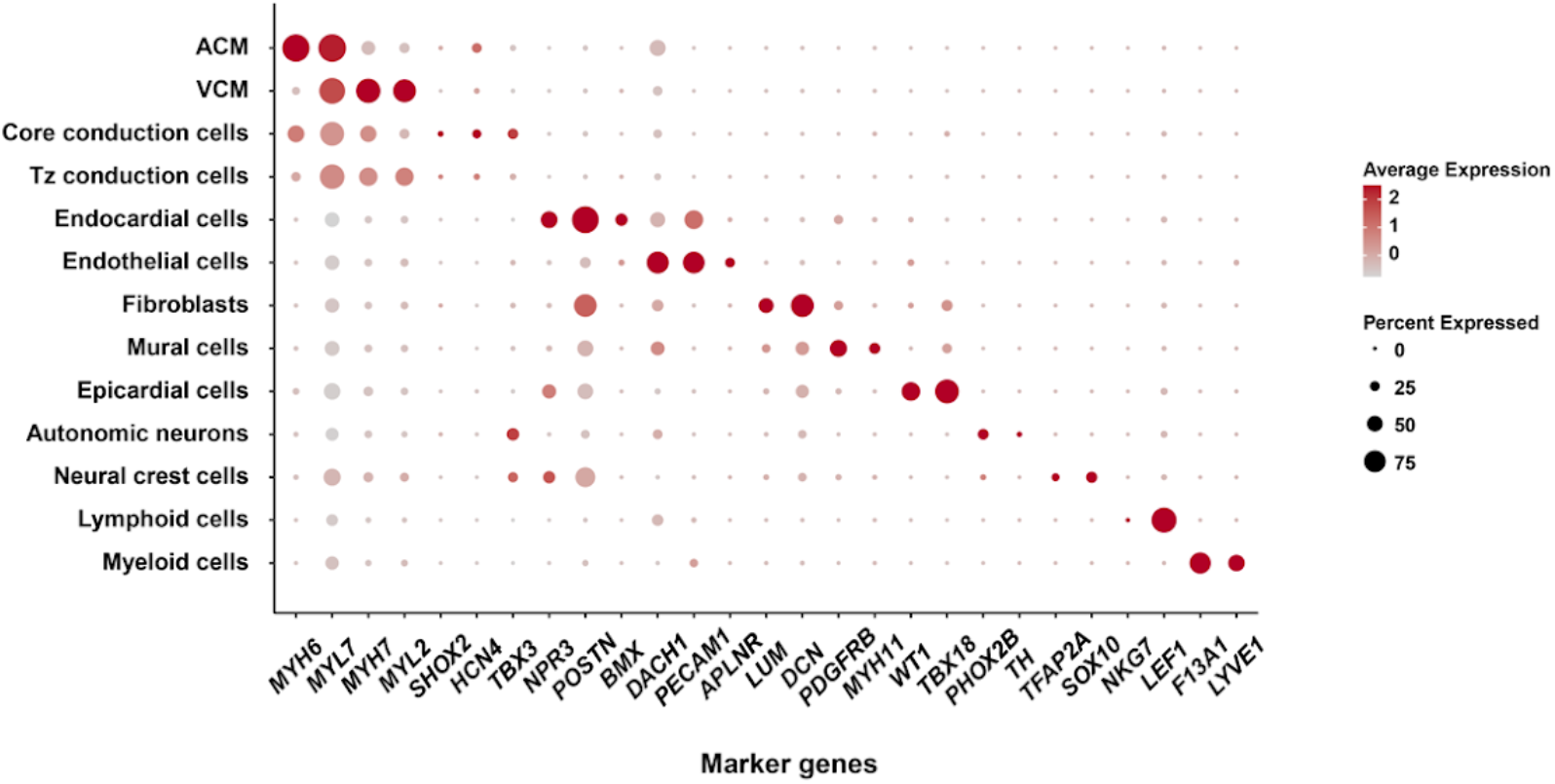
Annotation of major cell types. Dot plot of known marker genes for major cell types in the fetal heart atlas. Size of the circle represents the percentage of cells within a given cell type that express the gene of interest, and the color represents the normalized expression level of that gene across all cell types shown.

**Fig. S3:**
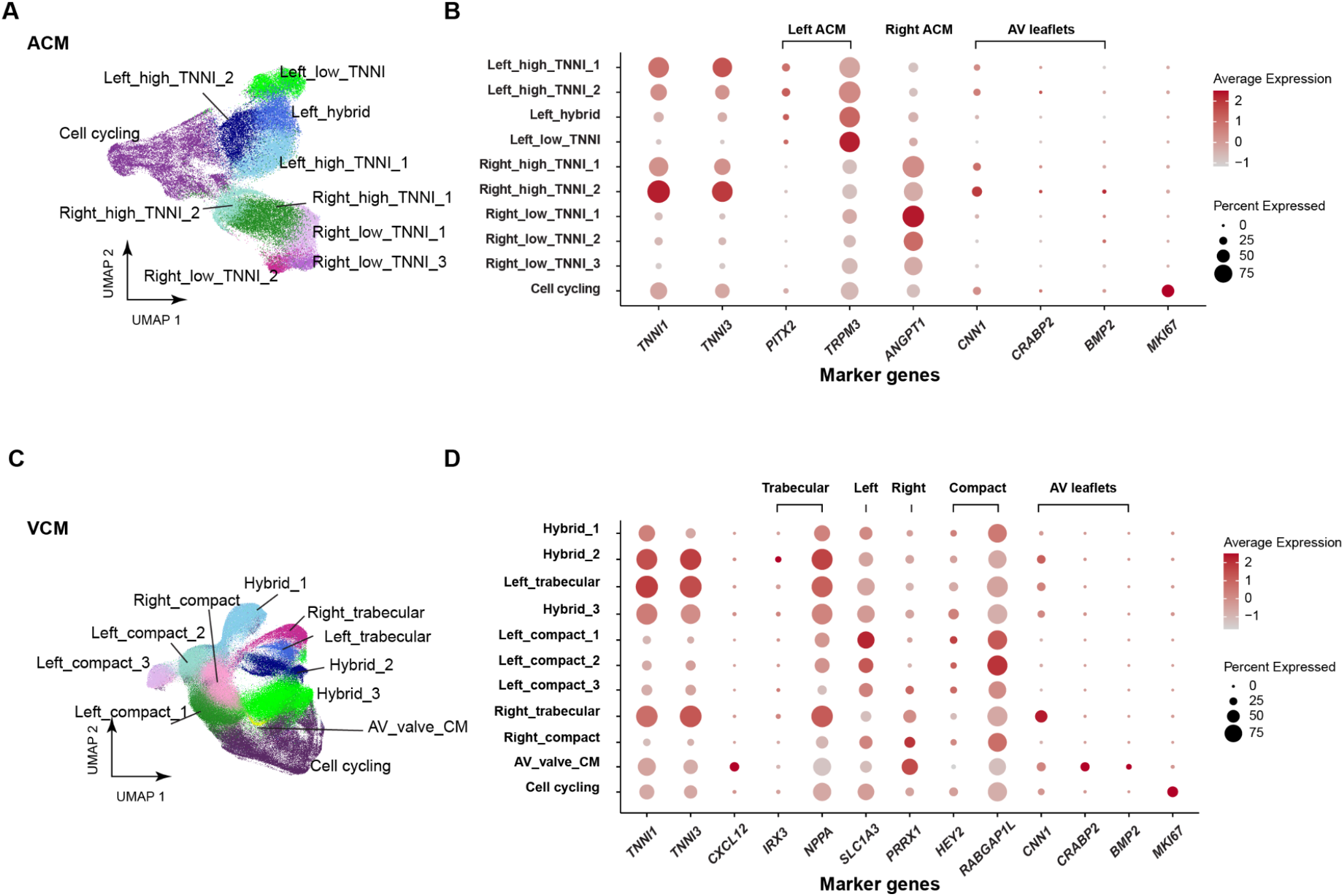
Annotation of cardiomyocytes. **A.** UMAP of ACMs. **B.** Dot plot of ACM marker genes. The size of the circle represents the percentage of cells within a given cell type that express the gene of interest, and the color represents the normalized expression level of that gene across all cell types shown. Left ACMs were identified using markers *PITX2* and *TRPM3*^48^. Right ACMs were identified using marker *ANGPT1*. As expected, ACMs with high *PITX2* and *TRPM3* expression exhibited lower *ANGPT1* expression, and vice versa. In addition to mapping ACMs to their spatial location (left vs. right atrium), ACMs can be categorized by their expression levels of Troponin (*TNNI1* and *TNNI3*). Certain ACM subtypes, termed “_high_TNNI,” express Troponin at a much higher level compared to others, designated “_low_TNNI.” *CNN1*, *CRABP2*, and *BMP2*, markers of cardiomyocytes in atrioventricular (AV) valve leaflets, were not significantly expressed in most ACMs, except for slightly elevated expression levels in ACM_right_high_TNNI_1. *MKI67* was used to identify cells undergoing active cell cycling. **C.** UMAP of VCMs. **D.** Dot plot of VCM marker genes. The size of the circle represents the percentage of cells within a given cell type that express the gene of interest, and the color represents the normalized expression level of that gene across all cell types shown. *SLC1A3* and *PRRX1* were used to distinguish left and right VCMs, respectively^48^. *IRX3*^48^ and *NPPA*^248^ were used to label trabecular VCM, while *HEY2* and *RABGAP1L* were used to annotate compact VCM. The “hybrid” cluster expressed both trabecular and compact VCM markers, suggesting a transitional state between the two cell types. Notably, trabecular VCM cell types exhibited much higher expression levels of Troponin compared to compact VCM. Additionally, *CRABP2*, *CNN1*, and *BMP2* were markers for cardiomyocytes in AV valve leaflets, and *CXCL12* was a marker for trabecular VCM near the lumen. *MKI67* was used to identify cells undergoing active cell cycling.

**Fig. S4:**
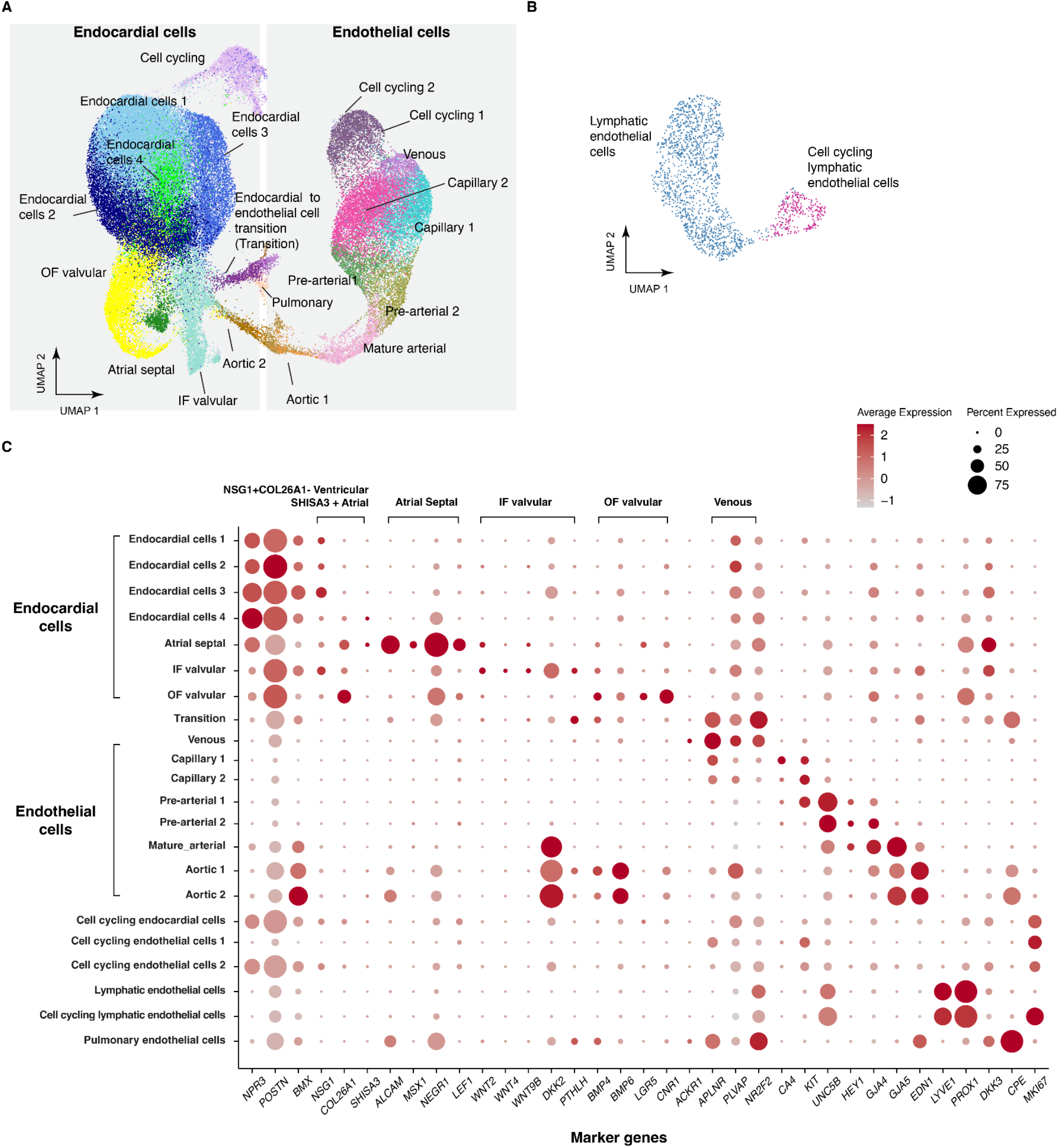
Annotation of endocardial cells and endothelial cells. **A.** UMAP of endocardial cells and endothelial cells (excluding lymphatic endothelial cells). IF: inflow. OF: outflow. **B.** UMAP of lymphatic endothelial cells. **C.** Dot plot of the marker genes for endocardial cells and endothelial cells. The size of the circle represents the percentage of cells within a given cell type that express the gene of interest, and the color represents the normalized expression level of that gene across all cell types shown. The expression of *NPR3*, *POSTN,* and *BMX* were used to distinguish endocardial cells from endothelial cells. Atrial septal endocardial cells were identified based on the expression levels of *ALCAM*, *MSX1*, *NEGR1*, and *LEF1*^49^. We identified endocardial cells on the IF side of the valves based on the expression of *WNT2*, *WNT4*, *WNT9B*, *DKK2*, and *PTHLH*, while those on the OF side were identified by *BMP4*, *BMP6*, *LGR5*, and *CNR1* expression. Finally, endocardial cells transitioning to endothelial cells (“transition”) were identified by the expression of *APLNR*, *ACKR1*, and *NR2F2* ^53,54^. Venous endothelial cells were identified by expression of *NR2F2*, *PLVAP*, and *APLNR*^53^. A cluster of cells likely to be pulmonary endothelial cells were labeled based on their expression of *DKK3* and *CPE*^249^. *CA4* and *KIT* were used to identify capillary endothelial cells. *UNC5B*, *HEY1*, *GJA4* and *GJA5* were used to identify arterial endothelial cells, with *GJA5* specifically marking mature arterial endothelial cells ^42,53^. Both *EDN1*^250^ and *BMX*^251^ were used to identify aortic endothelial cells. *MKI67* was used to identify cells undergoing active cell cycling.

**Fig. S5:**
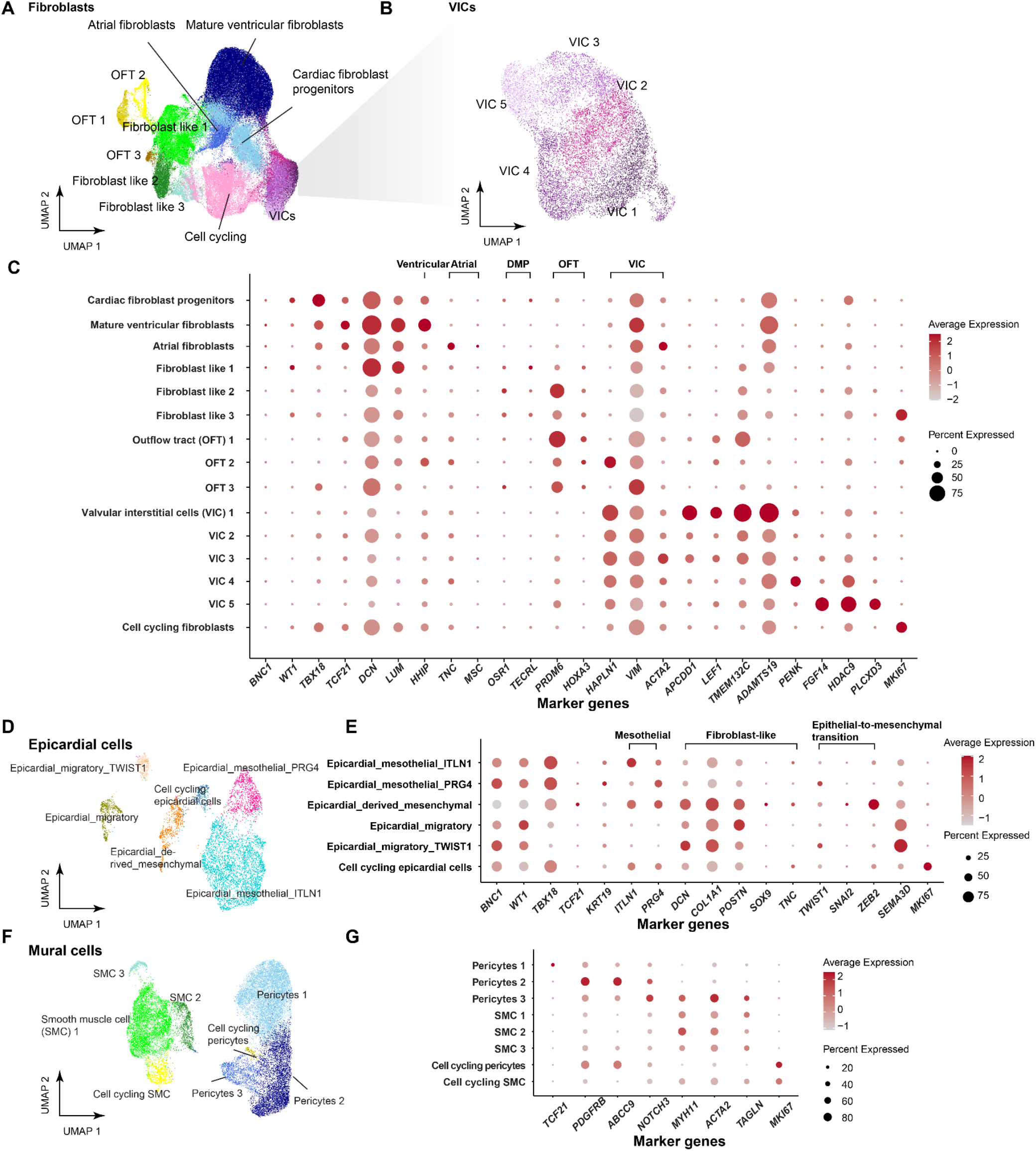
Annotation of mesenchymal and epicardial cells. **A.** UMAP of cardiac fibroblasts and VICs. OFT: Outflow tract. **B.** UMAP of VICs. **C.** Dot plot of the marker genes for fibroblasts and VICs. The size of the circle represents the percentage of cells within a given cell type that express the gene of interest, and the color represents the normalized expression level of that gene across all cell types shown. Cardiac fibroblast progenitors were identified by elevated expression of *WT1* and *TBX18*. *DCN* and *LUM* marked mature fibroblasts^4^. To identify chamber-specific fibroblasts, *HHIP* was used for ventricular fibroblasts, and *MSC* and *TNC* were used to identify atrial fibroblasts^48^. OFT cells were labeled with *PRDM6* and *HOXA3^4^*. OFT cells have lower expression of *TBX18* and *TCF21*, consistent with SHF origin. *HAPLN1*, *VIM,* and *ACTA2*^52,252^ were used to distinguish VICs from other fibroblast and fibroblast-like cells. VIC_1-3, particularly VIC_1, highly expressed *APCDD1*, *LEF1*, *TMEM132C,* and *ADAMTS19*, recently identified markers for VICs localized to the free segments of the valves^49^. Both VIC_4 and VIC_5 express recently observed markers of valve-related mesenchymal cells located in intervalvular fibrous tissue towards the valve roots. *PENK*, a marker for mesenchymal neural crest derivatives, was used to label VICs derived from neural crest (VIC_4), which were found to be located in and around the developing semilunar and AV valves. *FGF14*, *HDAC9*, *PLCXD3* are expressed in VIC_5, primarily observed around the semilunar valves. *MKI67* was used to identify cells undergoing active cell cycling. **D.** UMAP of epicardial cells. **E.** Dot plot of the marker genes for epicardial cells. The size of the circle represents the percentage of cells within a given cell type that express the gene of interest, and the color represents the normalized expression level of that gene across all cell types shown. Established epicardial markers such as *BNC1*, *TBX18*, *WT1*, *TCF21*, and *KRT19* were used to label epicardial cells^4,253^. We identified two distinct mesothelial epicardial populations: Epicardial_mesothelial_ITLN1 and Epicardial_mesothelial_PRG4. Epicardial_mesothelial_ITLN1 expresses *ITLN1*, a marker associated with epicardial adipose tissue; Epicardial_mesothelial_PRG4 expresses *PRG4*, a gene encoding lubricin secreted into pericardial fluid. Two migratory epicardial cell populations were identified: Epicardial_migratory and Epicardial_migratory_TWIST1, with the latter exhibiting higher *TWIST1* expression ^253^. Both migratory epicardial cells express mesenchymal cell markers *DCN* and *POSTN*, suggesting these cells are in the process of transitioning to mesenchymal cells. Epicardial_derived_mesenchymal cells, identified by markers of epithelial-to-mesenchymal transition (*SNAI2* and *ZEB2*) and cardiac mesenchymal cell differentiation (*SOX9* and *TNC*)^254^, likely represent epicardium-derived mesenchymal cells. The expression patterns of *TCF21* and *TBX18* also align with the previous observation^255^ showing sequential reduction of *TCF21* and *TBX18* expression in maturing epicardium, and upregulation of *TCF21* in epicardium-derived mesenchymal cells. *MKI67* was used to identify cells undergoing active cell cycling. **F.** UMAP of mural cells, including SMCs and pericytes. **G.** Dot plot of the marker genes for mural cells. The size of the circle represents the percentage of cells within a given cell type that express the gene of interest, and the color represents the normalized expression level of that gene across all cell types shown. Expression of *PDGFRB* and *ABCC9* were used to identify pericytes^4^. Pericytes 1 exhibited elevated levels of *TCF21*, suggesting it might represent early pericytes. Pericytes 3 has the highest expression of *NOTCH3*, a gene known to be expressed at the site of arterialization^256^, and higher expression of SMC markers *ACTA2* and *MYH11*, as well as early smooth muscle marker *TAGLN*, suggesting the cells may represent the transition from pericytes to SMCs. SMC1-2 express SMC markers *ACTA2*, *MYH11*, and *TAGLN*, suggesting they are SMCs. *MKI67* was used to identify cells undergoing active cell cycling.

**Fig. S6:**
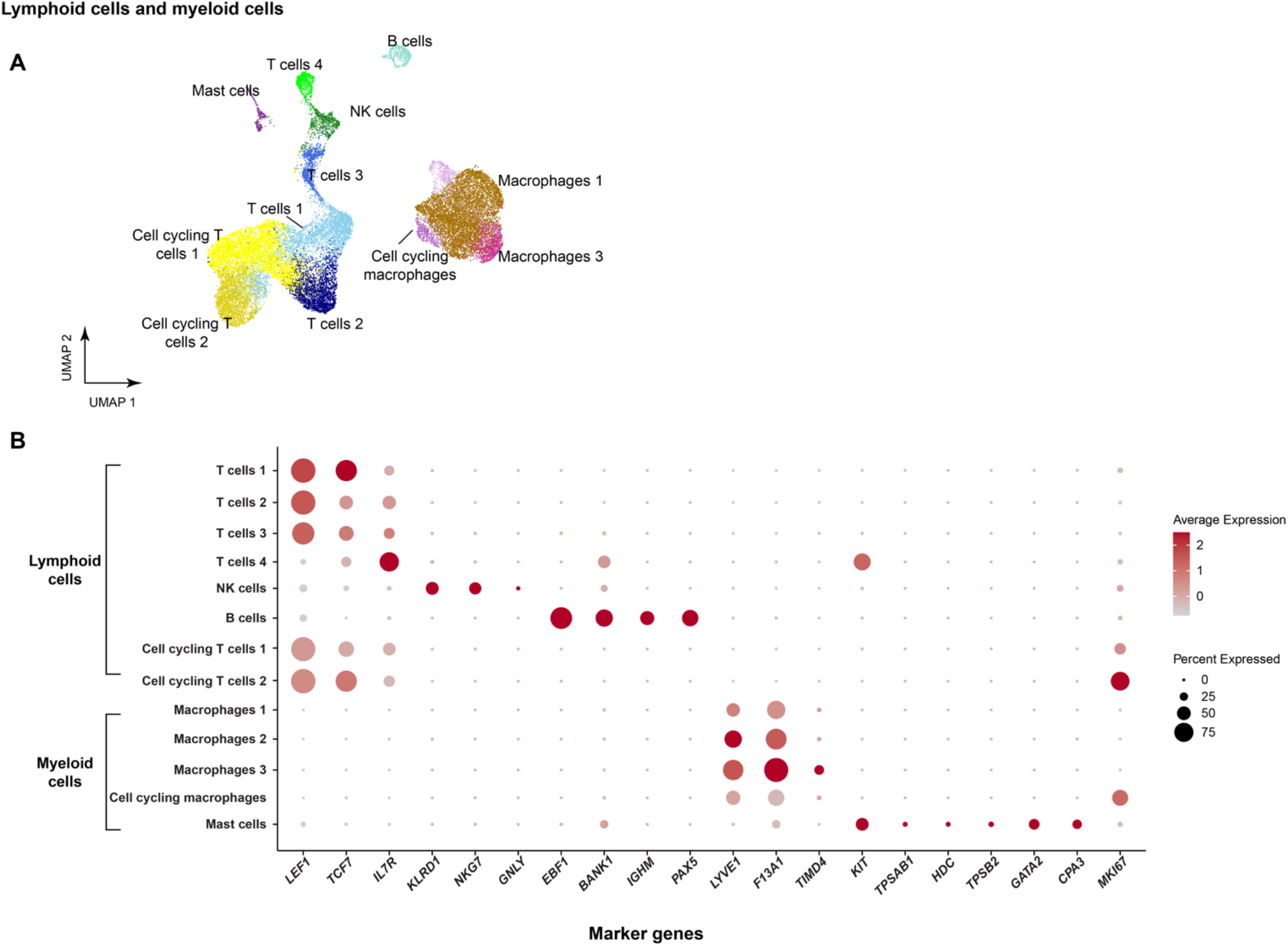
Annotation of immune cells. **A.** UMAP of lymphoid and myeloid cells. **B.** Dot plot of the marker genes for lymphoid and myeloid cells. The size of the circle represents the percentage of cells within a given cell type that express the gene of interest, and the color represents the normalized expression level of that gene across all cell types shown. Macrophages were identified using *LYVE1* and *F13A1*. Cells in “Macrophages_3” cluster are likely to be cardiac resident macrophages due to their expression of *TIMD4*^257,258^. T cells were identified using *LEF1*, *TCF7*, and *IL7R*^259^. *KLRD1*, *NKG7,* and *GNLY* were used to identify natural killer cells^260^. *EBF1*, *BANK1*, *IGHM,* and *PAX5* were used to identify B cells ^261,262^. Finally, *KIT*, *TPSAB1*, *HDC*, *TPSB2*, *GATA2*, and *CPA3* were used as markers of mast cells ^263–265^. *MKI67* was used to identify cells undergoing active cell cycling.

**Fig. S7:**
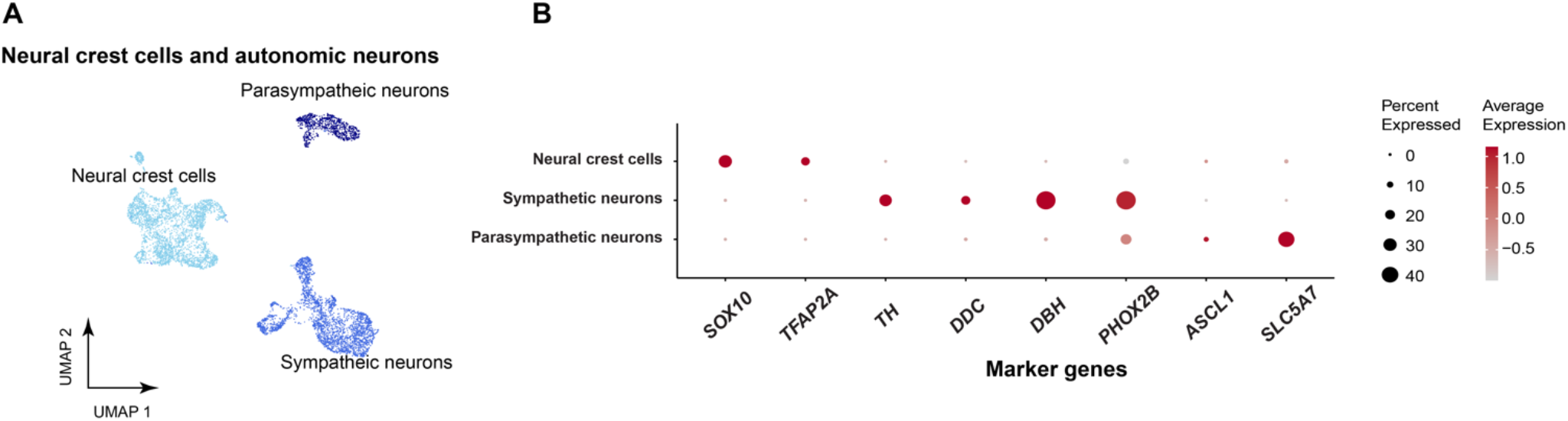
Annotation of neural crest cells and autonomic neurons. **A.** UMAP of neural crest cells and autonomic neurons. **B.** Dot plot of the marker genes for neural crest cells and autonomic neurons. The size of the circle represents the percentage of cells within a given cell type that express the gene of interest, and the color represents the normalized expression level of that gene across all cell types shown. Neural crest cells were identified using markers *SOX10* and *TFAP2A*^266^. *TH*, *DDC*, *DBH*, *and PHOX2B* were used to label sympathetic neurons^267,268^. *ASCL1*^269^ and *SLC5A7*^267^ were used to label parasympathetic neurons.

**Fig. S8.**
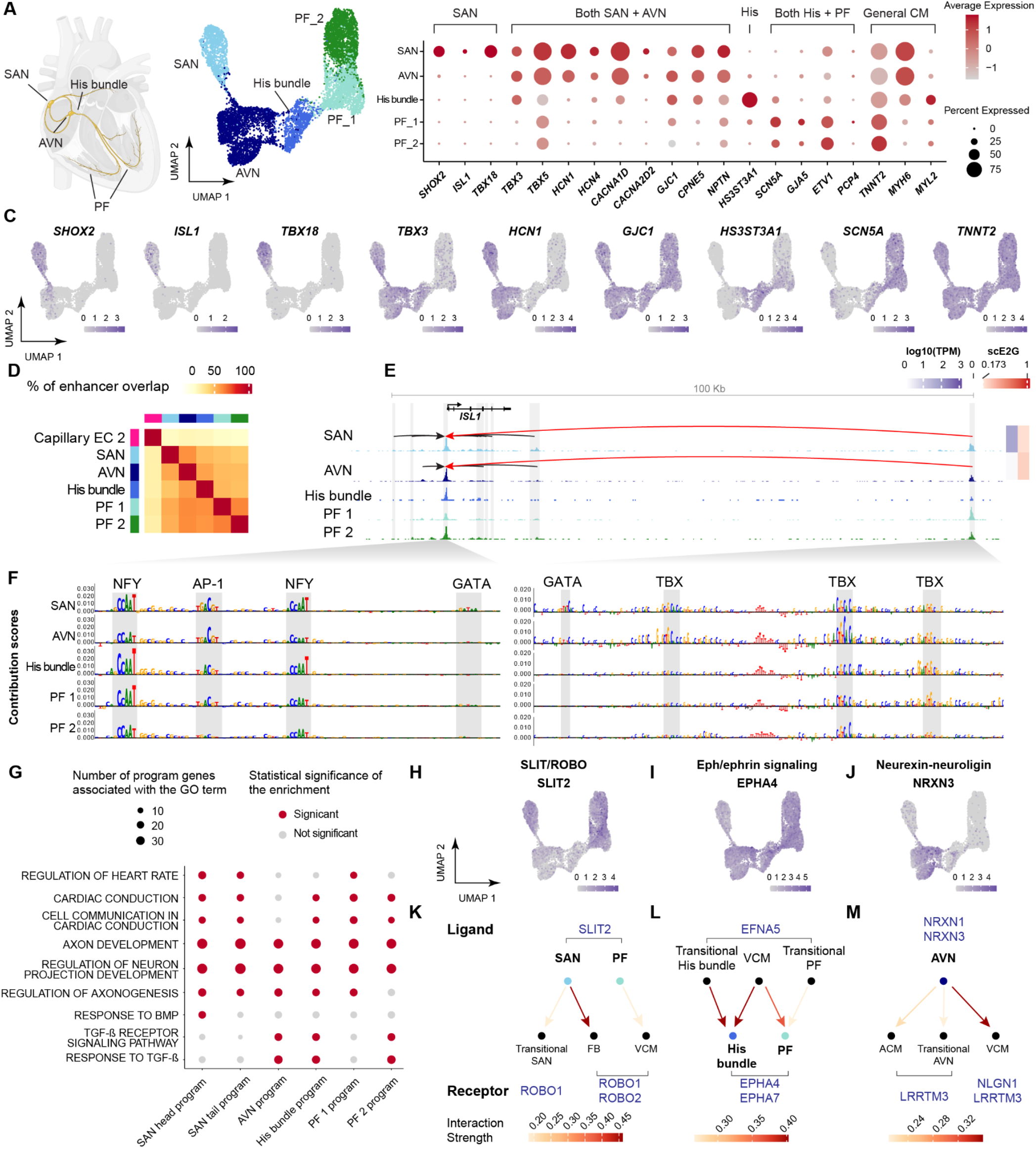
Regulatory wiring and neural adhesion molecules in cardiac conduction cells. **A.** Schematic and UMAP representations of cell types in the cardiac conduction system (CCS). SAN: Sinoatrial node. AVN: Atrioventricular node. PF: Purkinje fibers. **B.** Dot plot of known marker genes for major cell types in the CCS. The size of the circle represents the percentage of cells within a given cell type that express the gene of interest, and the color represents the normalized expression level of that gene across all cell types shown. **C.** Feature plots showing the expression of marker genes. Color scale: normalized gene expression level. **D.** Percentage of enhancer overlap between the cell types on the y-axis and cell types on the x-axis. Includes 5 cell types in the CCS and a population of capillary endothelial cells as an outgroup for comparison. Color scale: Percentage of enhancer overlap between pairs of cell types, with the total number of enhancers in the cell type on the y-axis as the denominator. **E.** scE2G predictions and normalized scATAC-seq signals for *ISL1*. Predicted enhancers and the *ISL1* promoter are highlighted in shaded regions. Arcs represent the predicted enhancer-gene regulatory interactions. Side color bars show the TPM expression of the target genes in each cell type (blue) and the corresponding scE2G scores (red, and white for scores below the threshold). **F.** Per-base DeepLIFT contribution score profiles of two selected regulatory regions from subpanel E. Gray highlights indicate predicted TF motif instances, including for GATA, NF-Y, and TBX family TFs. Note that GATA motif instances have higher predicted contribution scores in SAN. The left panel shows a subregion of the *ISL1* promoter (chr5:51,383,321-51,383,428). The right panel shows a subregion of an enhancer regulating *ISL1* (chr5:51,475,542-51,475,802). All coordinates in hg38. **G.** Representative enriched gene ontology (GO) terms in gene programs characterizing the CCS cell types, showing enrichment for genes related to axon development across all CCS cell types. **H. I. J.** Feature plot showing the expression of selected neural adhesion ligands or receptors in the CCS. **K. L. M.** Predicted CCI between CCS components and surrounding cell types. Nodes represent cell types, with CCS components highlighted in bold text and colored according to A. Arrows point from ligand-producing cells to receptor-expressing cells. Arrow color intensity reflects the interaction strength.

**Fig. S9:**
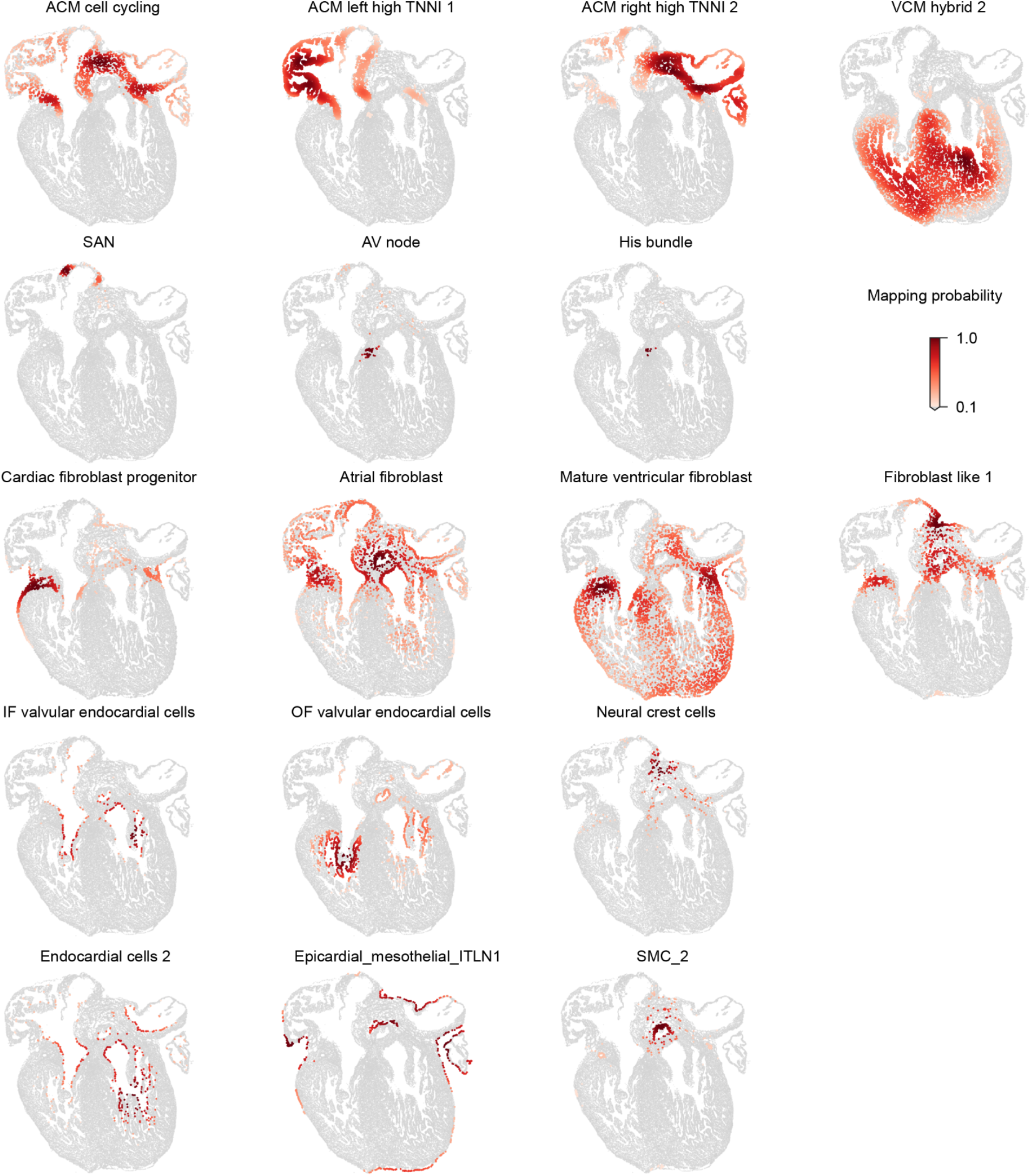
Orthogonal validation of spatial annotations using MERFISH spatial transcriptomics data. Representative images showing the fetal heart cell type atlas spatially mapped onto a cross-sectional heart image (R77_4C4)^48^. Mapping was restricted to cell types corresponding to the developmental stages of the MERFISH-profiled heart. Color intensity indicates the mapping probability for each cell type.

**Fig. S10:**
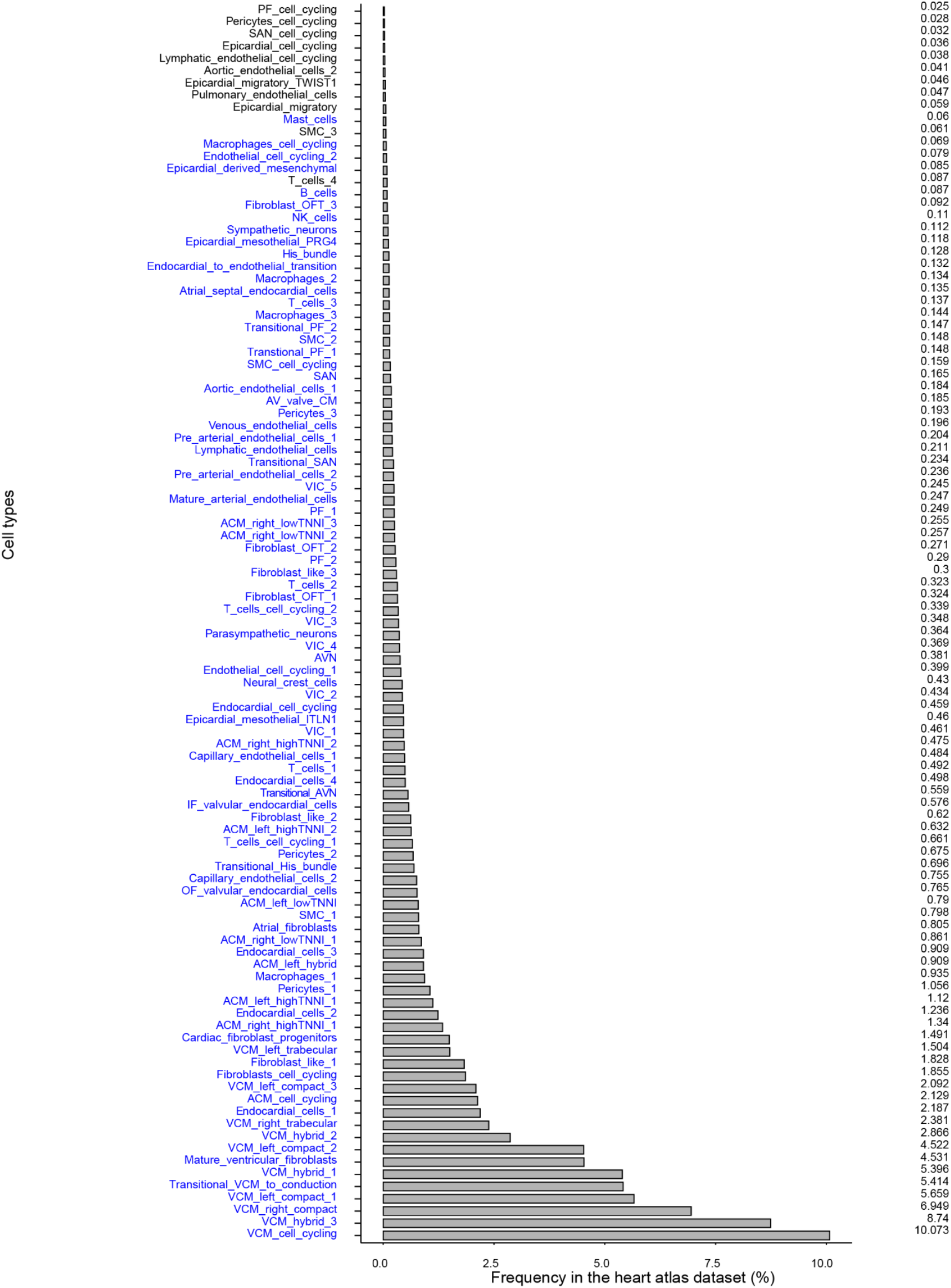
Cell type frequencies. Barplot showing the frequency of each cell type in the heart atlas. Highlighted in blue are cell types with sufficient ATAC fragment counts for scE2G and ChromBPNet predictions. PF: Purkinje fibers. SAN: Sinoatrial node. SMC: Smooth muscle cells. NK: Natural killer cells. OFT: outflow tract. AV: Atrioventricular. ACM: Atrial cardiomyocytes. VIC: Valvular interstitial cells. AVN: Atrioventricular node. VCM: ventricular cardiomyocytes.

**Fig. S11:**
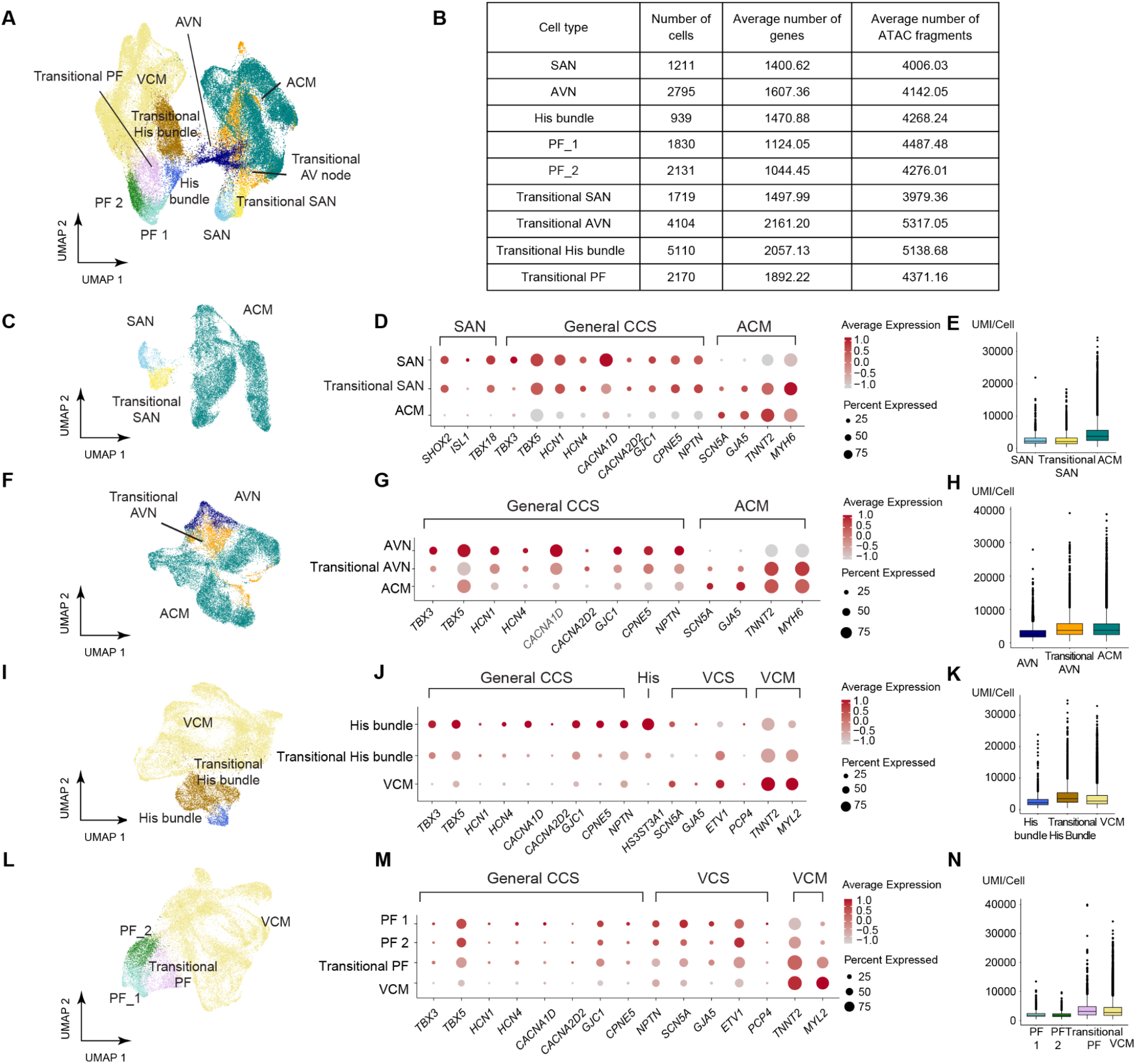
Transitional conduction cells in the heart atlas.A. UMAP of the conduction cells, transitional conduction cells, and the surrounding working myocardium. ACM: Atrial cardiomyocyte. AVN: Atrioventricular node. PF: Purkinje fiber. SAN: Sinoatrial node. VCM: Ventricular cardiomyocyte. **B.** Cell count, average number of genes per cell, and average number of ATAC fragments per cell for each conduction type and transitional conduction cell type. **C,F,I,L.** Each UMAP represents a conduction system cell type, its corresponding transitional cell types, and the surrounding working myocardium. **D,G,J,M.** Dot plots of known marker genes for conduction cells, corresponding transitional cell types, and surrounding working myocardium. CCS: Cardiac conduction system. VCS: Ventricular conduction system. **E,H,K,N.** Comparison of number of UMI/cell among conduction cells, corresponding transitional cells and surrounding working myocardium. The UMI/cell of transitional cells was not the sum of the average UMI/cell of conduction cells and the average UMI/cell of surrounding working myocardium. Given this finding and that doublets were pre-emptively removed during quality control, it is unlikely that these transitional cells reflect multiplets.

**Fig. S12:**
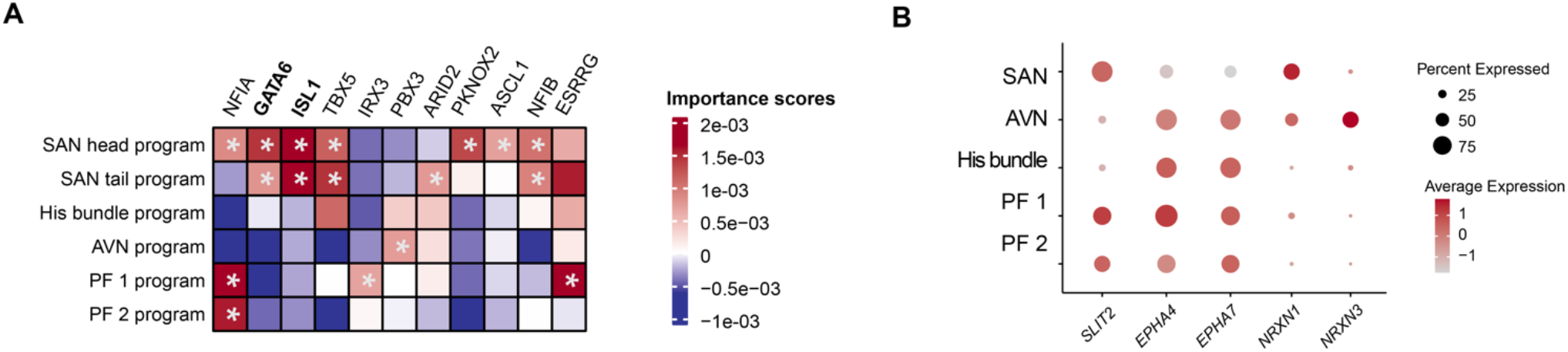
Cardiac conduction cells: TF regulators of gene programs and expression of neural adhesion and axon development genes. **A.** Importance scores of TFs predicted to regulate gene programs associated with the identity of each conduction system cell type. Bold TFs are discussed in the text. *: the TF is a regulator of the corresponding gene program. SAN: Sinoatrial node. AVN: Atrioventricular node. PF: Purkinje fibers. **B.** Dot plot showing the expression pattern of genes involved in neural adhesion and axon development (see also **Fig. S8K-M)**. The size of the circle represents the percentage of cells within a given cell type that express the gene of interest, and the color represents the normalized expression level of that gene across all cell types shown.

**Fig. S13:**
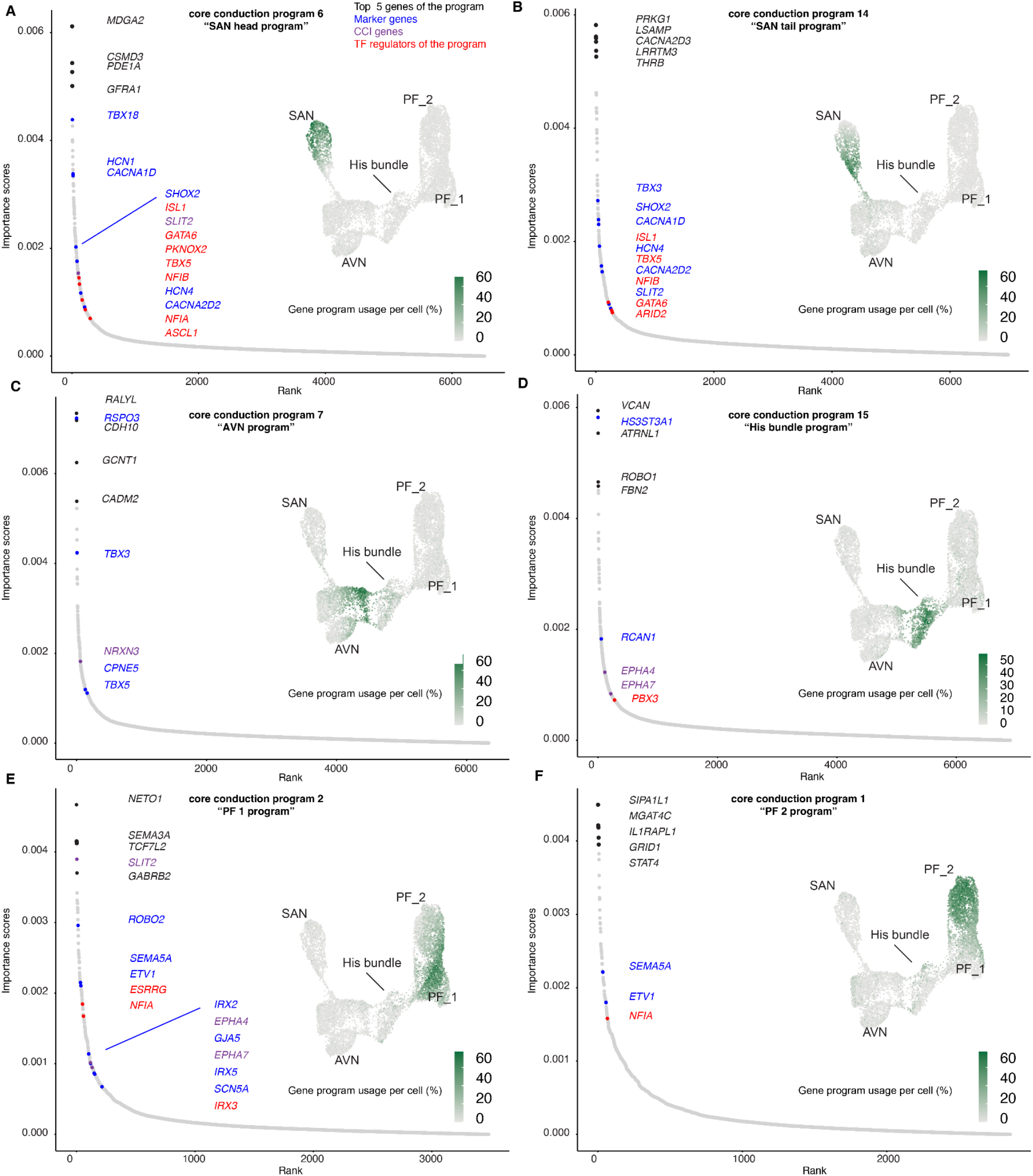
Gene programs of conduction system cell types. Annotated gene programs associated with the identity of each conduction system cell type: SAN head program (**A**); SAN tail program (**B**); AVN program (**C**); His bundle program (**D**); PF 1 program (**E**); PF 2 program (**F**). Waterfall plot shows importance scores for all considered genes, and selected genes from the 300 top program genes are colored. Black: 5 genes with the highest importance scores. Blue: Known marker genes for the corresponding cell type. Purple: Genes involved in important cell-cell interaction (CCI) pathways (from **Fig. S12**) are highlighted. Red: TFs predicted to regulate the program. Inset UMAPs: Projection of gene program expression on the cardiac conduction cell UMAP (see also **Fig. S8A**). Color scale: gene program usage per cell. SAN: Sinoatrial node. AVN: Atrioventricular node. PF: Purkinje fibers.

**Fig. S14.**
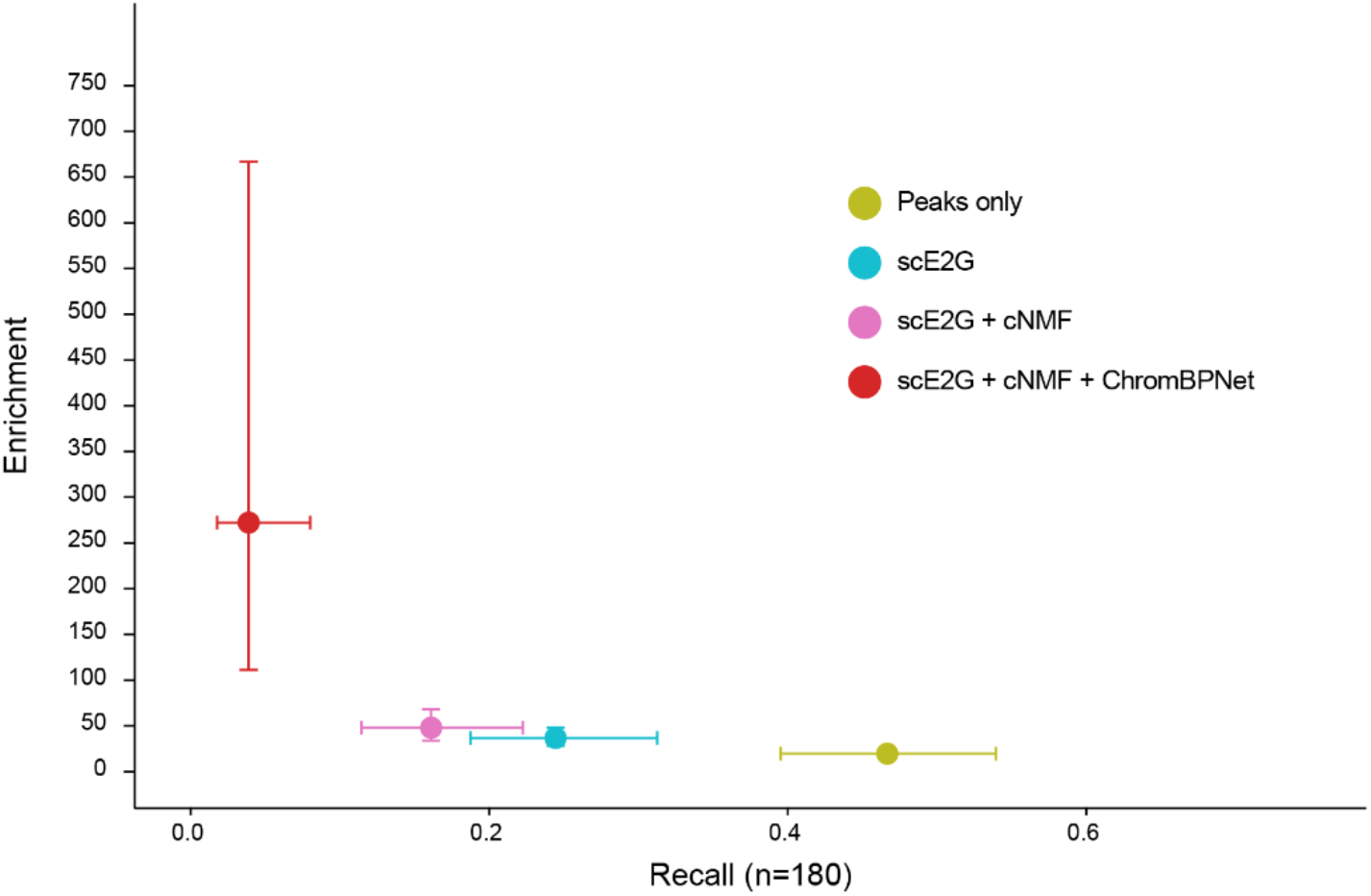
Integrating scE2G, cNMF, and ChromBPNet significantly enhances variant enrichment. Enrichment versus recall (n=180) for fine-mapped quantitative cardiac trait variants across four progressive feature sets: (1) Peaks only, (2) scE2G applied to peaks, (3) scE2G + cNMF (scE2G genes linked to concatenated cNMF-derived gene programs), and (4) scE2G + cNMF + ChromBPNet (further requiring variants to induce significant predicted changes in chromatin accessibility). Notably, the integration of all three methods yields an extremely strong ∼270-fold enrichment against background variants, outperforming the baseline methods alone. Horizontal and vertical error bars represent 95% confidence intervals of the fold enrichment.

**Fig. S15:**
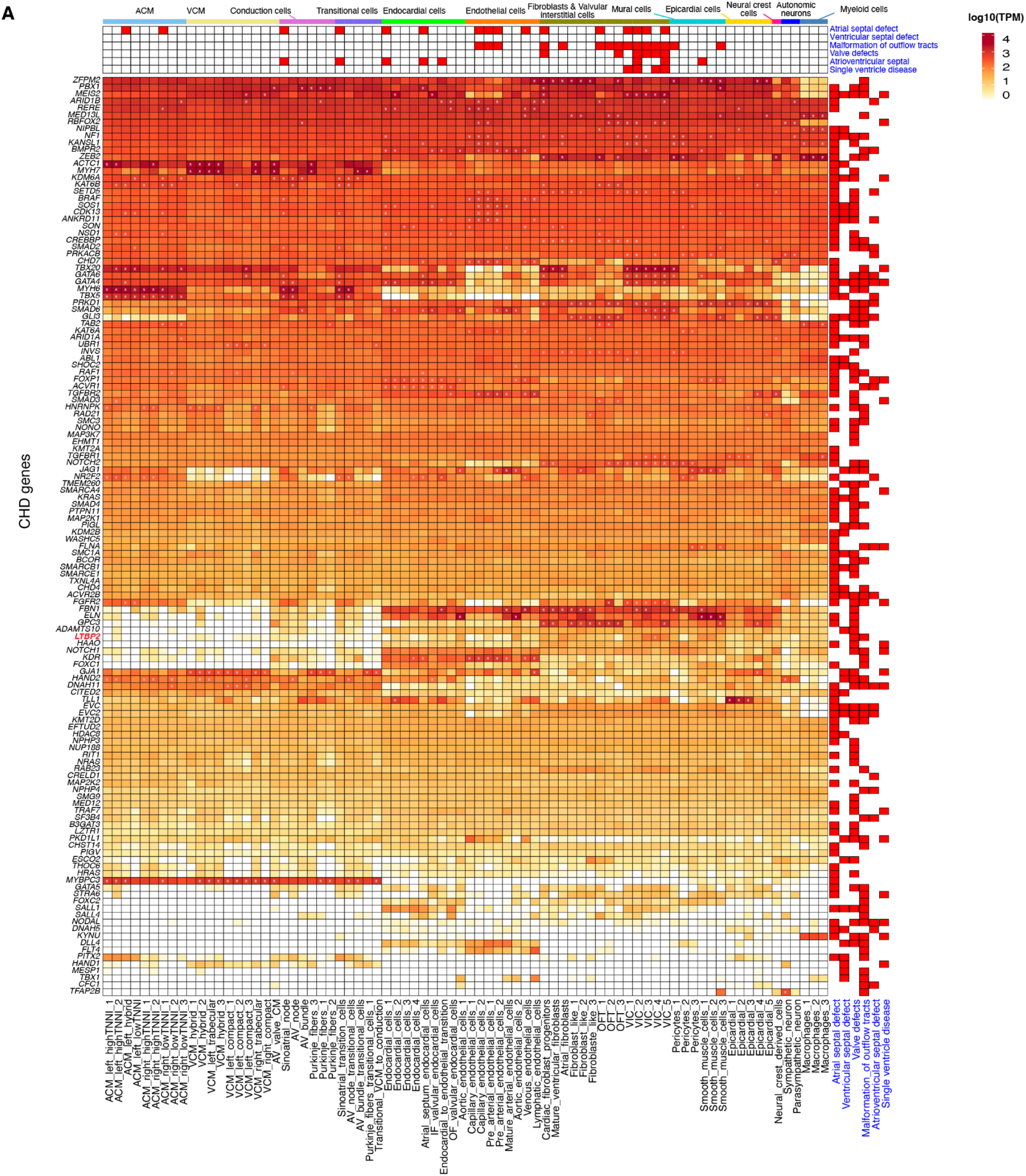
Expression of high-confidence CHD genes across cardiac cell types. Absolute expression of high-confidence CHD genes across cardiac cell types. Overall, many CHD genes are expressed in multiple cell types. However, certain cell types are enriched for having many CHD genes with particularly high expression. Main heatmap color scale: log_10_ TPM. Top annotation: red indicates that the cell type (column) is significantly enriched for high expression of the high-confidence genes associated with the corresponding CHD subtype (row) (see **Fig. 2A**). Note the absence of significant enrichment in the Ventricular Septal Defect (VSD) row; this is expected given that our earliest samples (6 PCW) capture the heart toward the tail end of ventricular septation (completed by 7 PCW), likely missing the earlier, transient progenitor windows that drive this process^270^. Side annotation: red indicates that the corresponding gene (row) is a high-confidence causal gene for a CHD subtype (columns). *: The gene is considered highly expressed in the corresponding cell type (see **Methods**). PF: Purkinje fibers. SAN: Sinoatrial node. SMC: Smooth muscle cells. NK: Natural killer cells. OFT: outflow tract. AV: Atrioventricular. ACM: Atrial cardiomyocytes. VIC: Valvular interstitial cells. AVN: Atrioventricular node. VCM: ventricular cardiomyocytes.

**Fig. S16:**
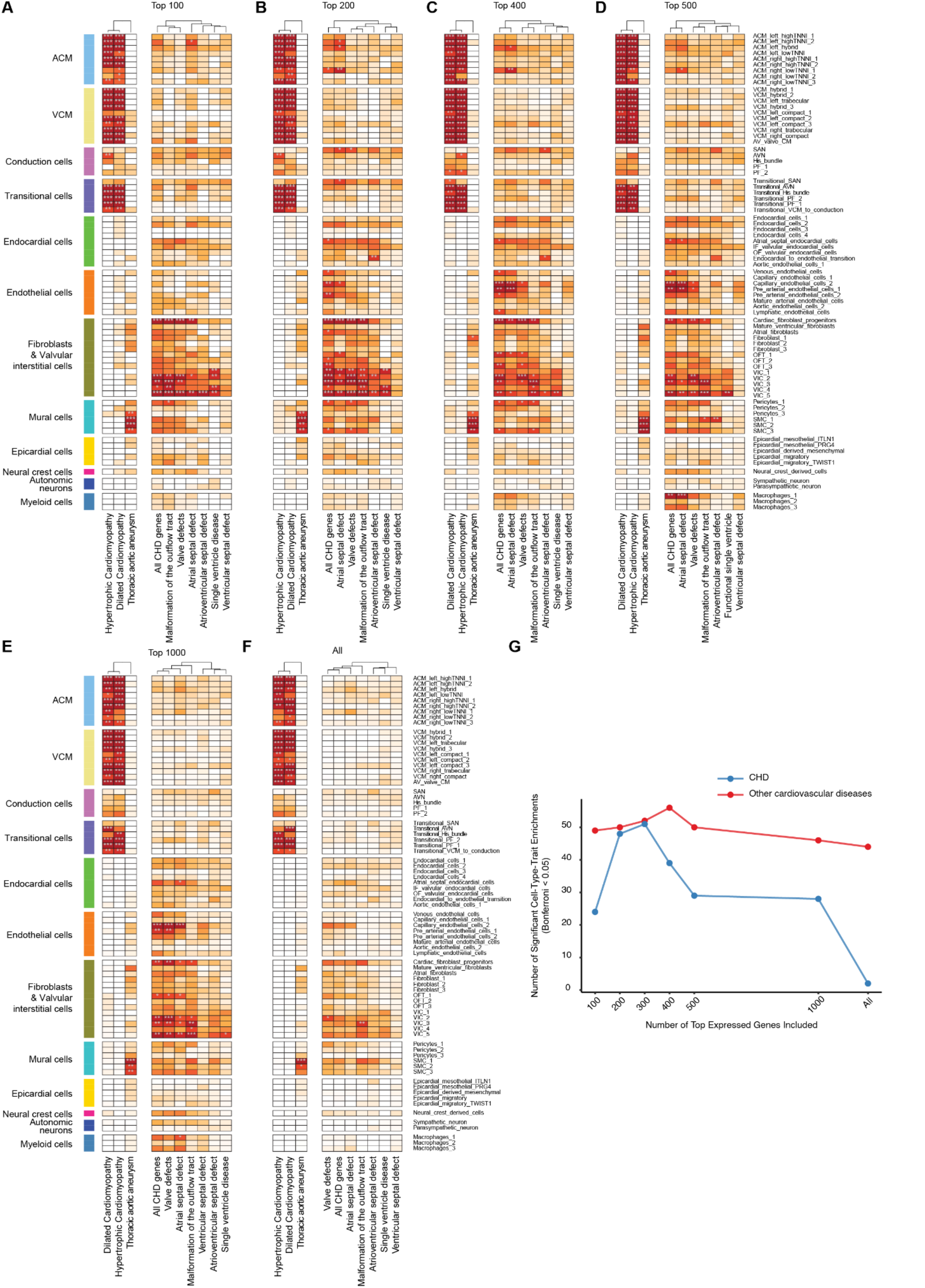
Parameter sensitivity analysis of disease enrichments.A-F. Enrichment for cardiovascular disease and CHD causal genes in cardiac cell types of the fetal heart map at different thresholds: Top 100, Top 200, Top 400, Top 500, Top 1000, and All genes (z-score >1). Color scale represents −log10-transformed P-values from a one-sided Fisher’s exact test with Bonferroni correction (∗P_Bonferroni<0.05, P_Bonferroni<0.01, ∗∗∗P_Bonferroni<0.001). High-confidence background genes include all genes expressed in any considered cell type (TPM>1). **G.** Total number of significant cell-type-trait enrichments (P_Bonferroni<0.05) detected across all tested thresholds. CHD: Congenital heart defect. Other cardiovascular diseases: Dilated Cardiomyopathy, Hypertrophic Cardiomyopathy, Thoracic Aortic Aneurysm.

**Fig. S17:**
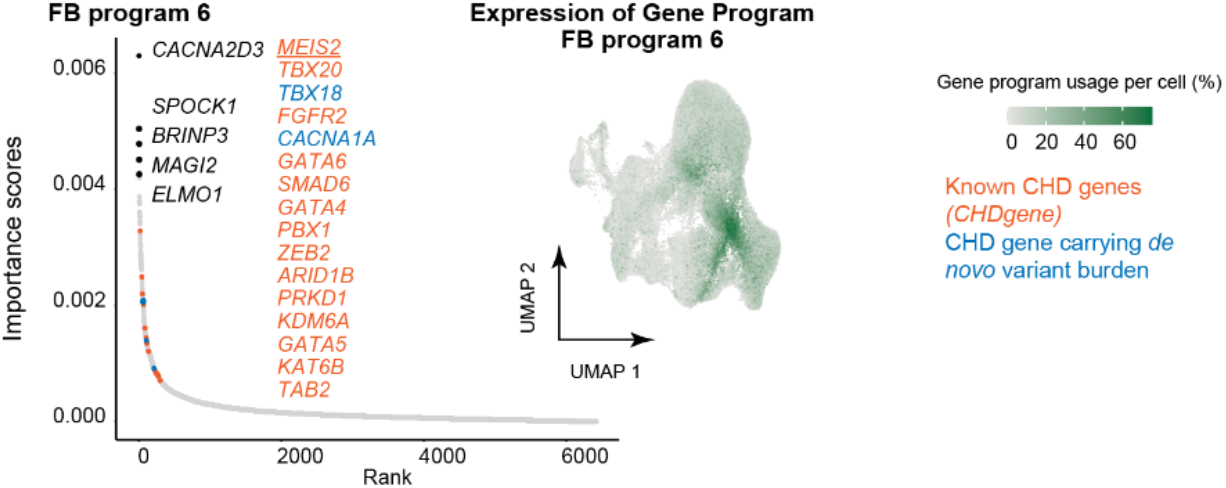
Gene importance score ranking and expression profile of FB program 6. Ranked importance scores of genes associated with FB program 6. Labels denote selected program genes (among the 300 by importance score). Black: Top 5 genes associated with FB program 6. Orange: high-confidence CHD genes from CHDgene. Blue: CHD genes carrying a significant burden of heterozygous *de novo* or rare transmitted damaging variants^7^. *GATA6*, *SMAD6*, *GATA4* and *ZEB2* are both high-confidence CHD genes and genes carrying two or more *de novo* variants. Inset: Projection of FB program 6 expression on the fibroblast UMAP, showing high expression in cardiac fibroblast progenitors and some cell cycling fibroblasts.

**Fig. S18:**
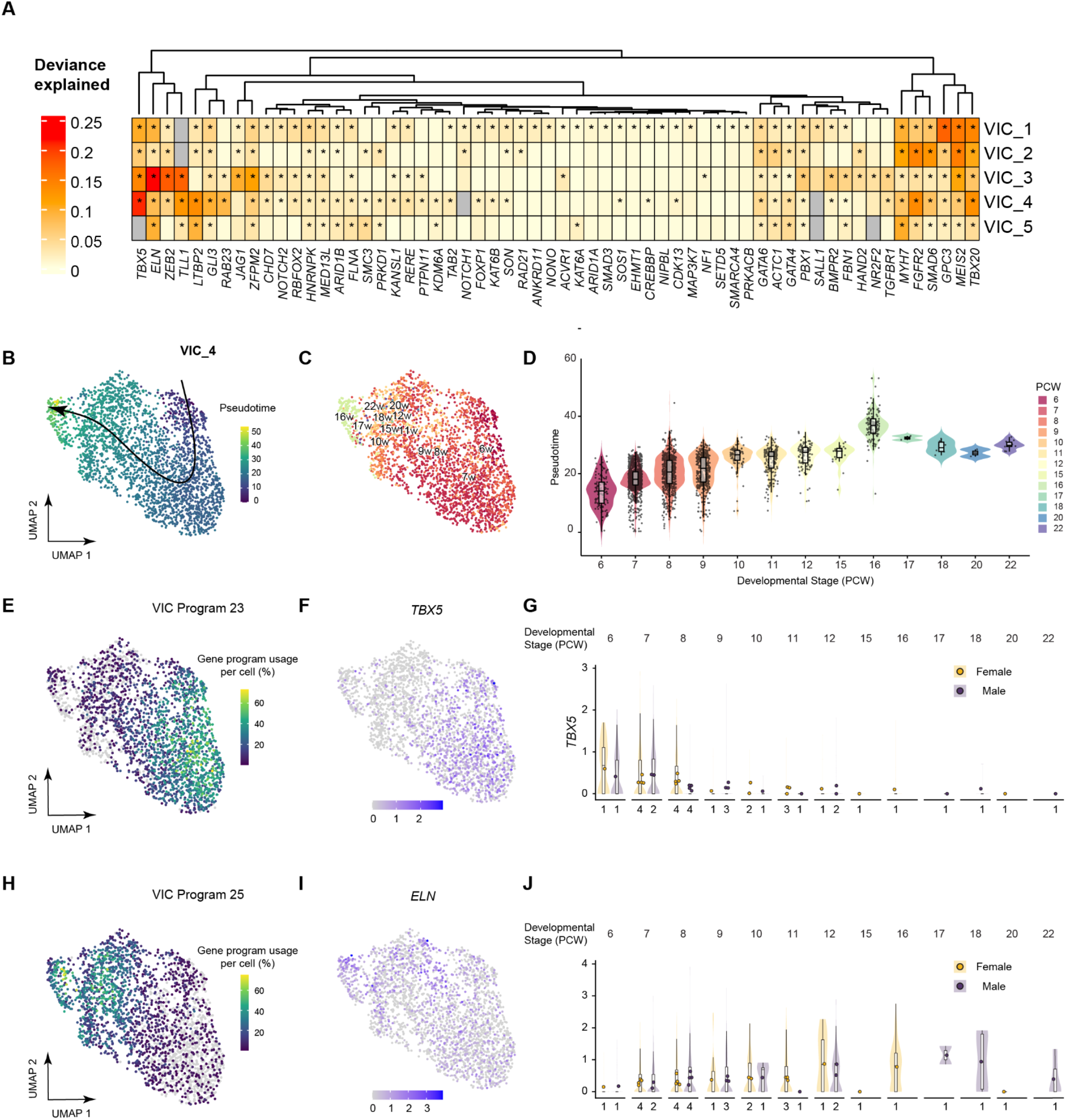
Pseudotime trajectories and gene expression dynamics of the VIC subpopulations during human heart valve development. **A.** Heatmap of expression deviance explained by pseudotime trajectories for high-confidence CHD genes across VIC subpopulations. Asterisks denote significant lineage associations (FDR < 0.05). **B.** UMAP embedding of the VIC_4 subpopulation, colored by Slingshot-inferred pseudotime. The solid line and arrow indicate the developmental trajectory from early to late stages, with initiation and termination anchored by known PCW. **C.** UMAP embedding as in **B**, coloured by developmental stage (PCW). Cell ordering is concordant with the inferred pseudotime. **D.** Distribution of pseudotime values across developmental stages. Overlap in pseudotime between stages reflects differential cellular maturity among cells sampled at single time points. Box plots show the median (horizontal line) and interquartile range (IQR, extending from the 25th to the 75th percentile); whiskers extend to the furthest data point within 1.5 × IQR from the hinges. Points represent individual cells. **E.** UMAP projections showing the per-cell usage percentage of VIC gene program 23 (**E**) and program 25 (**H**). **F.** UMAP projections of normalized, log-transformed expression levels for TBX5 (**F**) and ELN (**I**). **G.** Violin plots of normalized, log-transformed expression for TBX5 (**G**) and ELN (**J**) across developmental stages, stratified by sex (female, yellow; male, purple). Box plot elements are defined as in **D**. Points denote the average gene expression per individual. The numbers below each violin indicate the sample size for each sex at that stage.

**Fig. S19:**
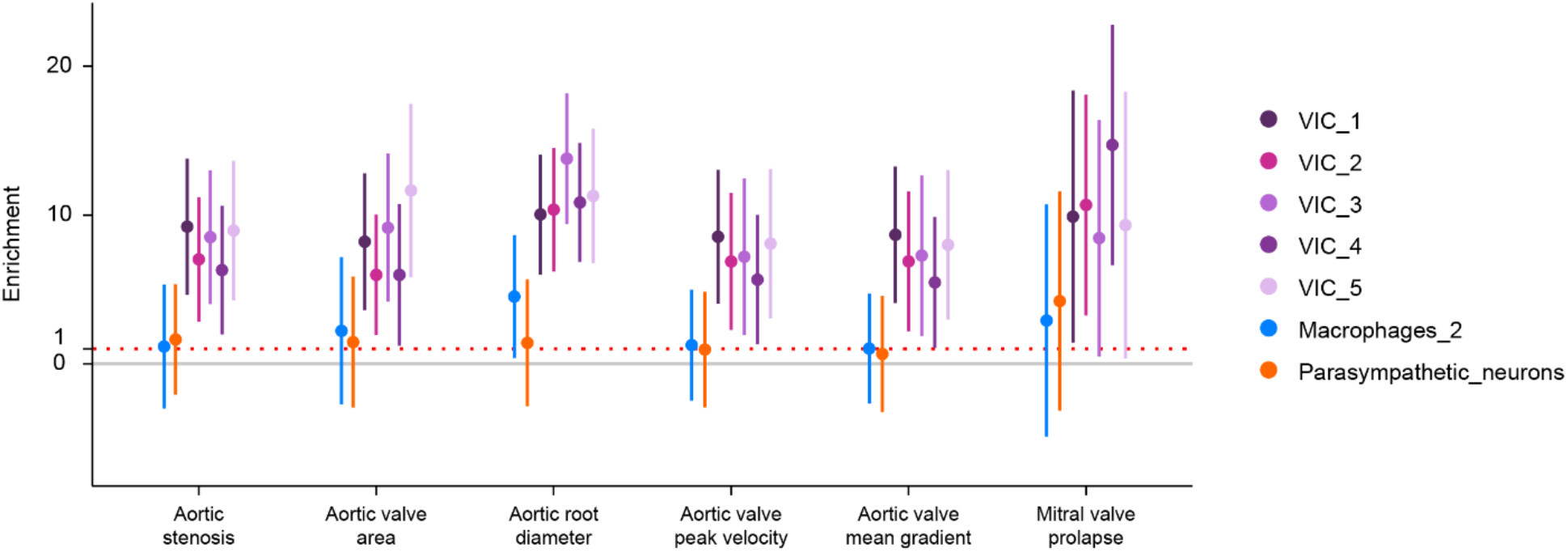
Enrichment of heritability for 6 valve and aorta-related traits in VIC enhancers. Y-axis: Genome-wide heritability enrichment of each trait in scE2G-predicted enhancers in each of the cell type, as calculated by S-LDSC. Error bars represent 95% confidence intervals; the horizontal dotted line represents a null enrichment of 1. See Table S15 for full results.

**Fig. S20:**
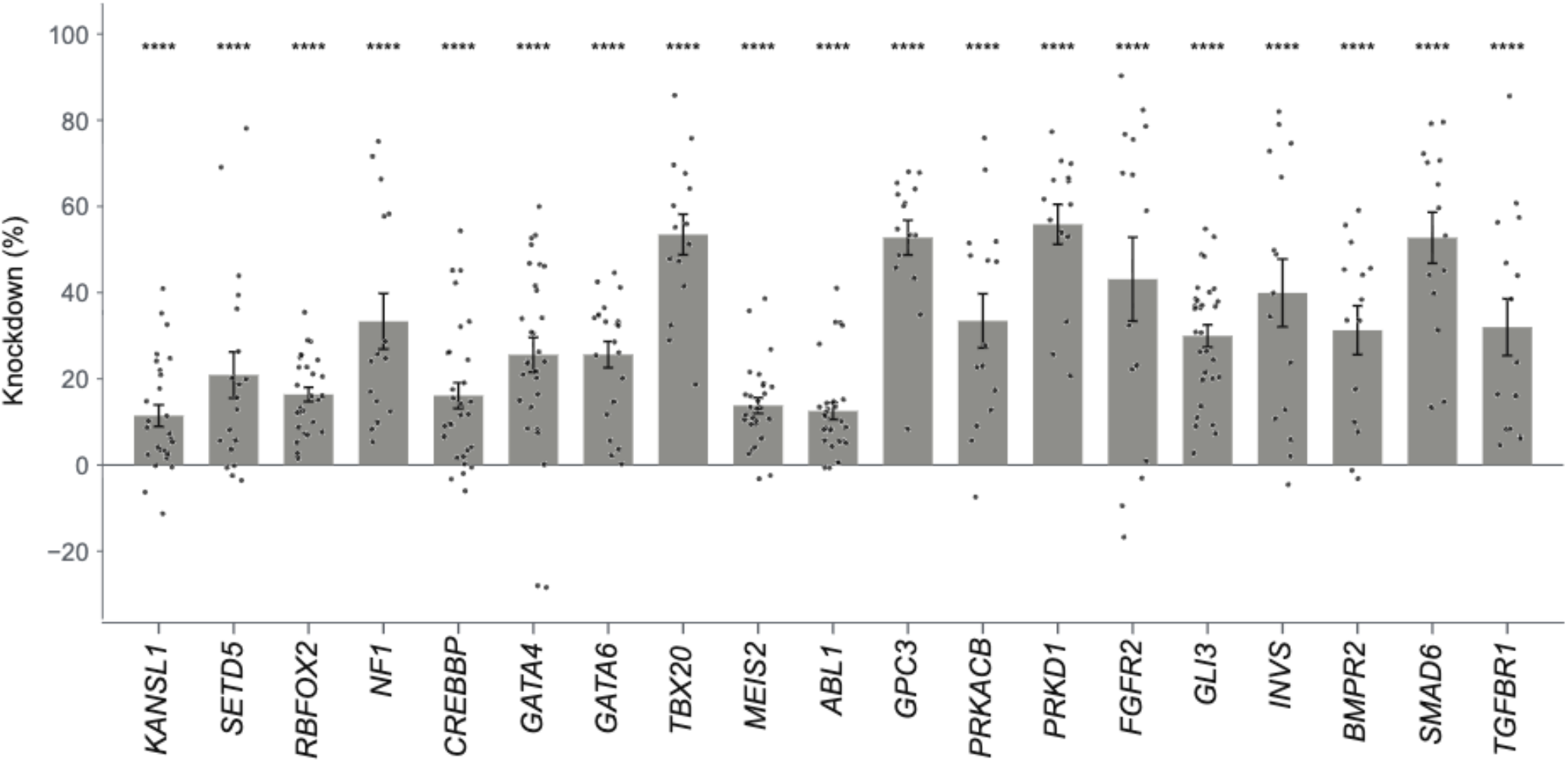
On-target CRISPRi knockdown of CHD risk genes in hiPSC-VICs. Per-guide target knockdown was calculated through a modified version of SCEPTRE where a treatment-free Poisson GLM of each target gene’s counts on batch and log library size was fitted across all cells, and each gRNA’s log2 fold change taken as log2(summed observed counts ⁄ summed fitted values) over the cells assigned that gRNA, expressed as percent knockdown. Each dot represents an individual gRNA. Error bars represent standard error of the mean across gRNAs per target. Significance was calculated using a two-sided Mann-Whitney U-test comparing each target gene’s per-gRNA pseudobulk expression against the 200 NT gRNAs, then adjusted using the Benjamini-Hochberg method across all panel genes. **** = q < 0.0001

**Fig. S21:**
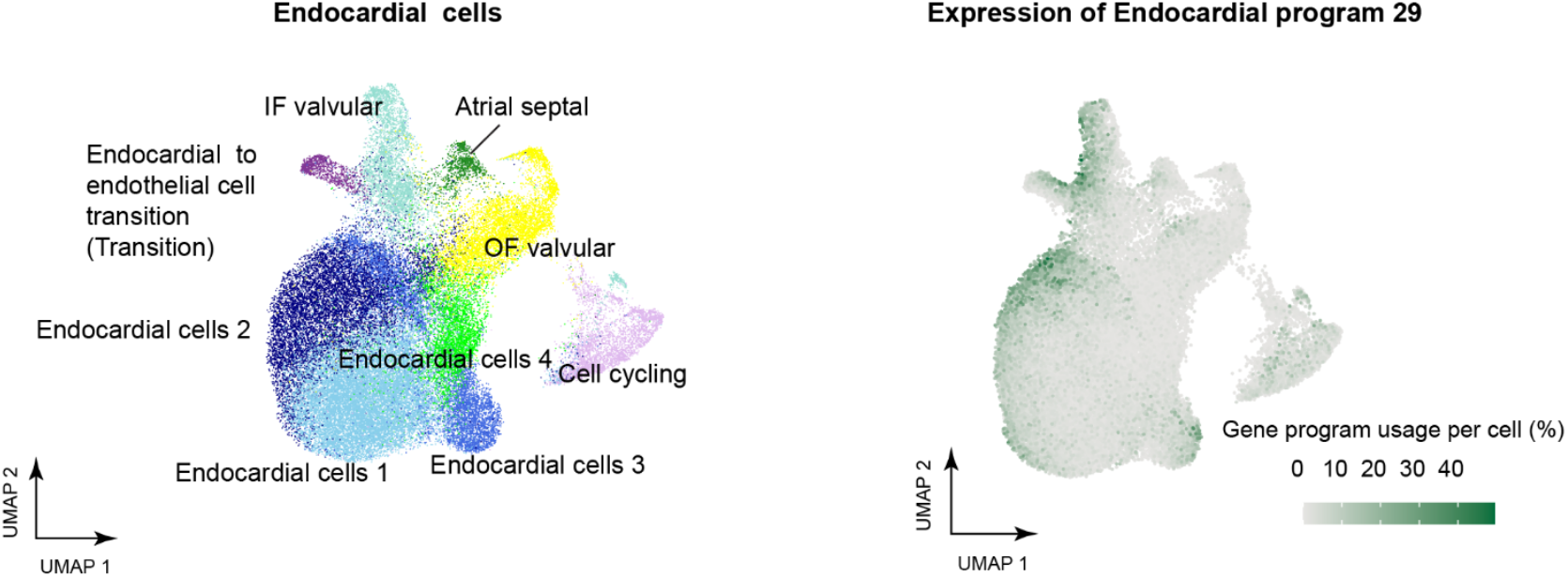
**Expression of endocardial program 29 in endocardial cells.**UMAP of endocardial cells and the projection of Endocardial Program 29 expression onto the endocardial cell UMAP. This program is expressed in subpopulations of multiple endocardial cell types. Color scale: gene program usage per cell.

**Fig. S22:**
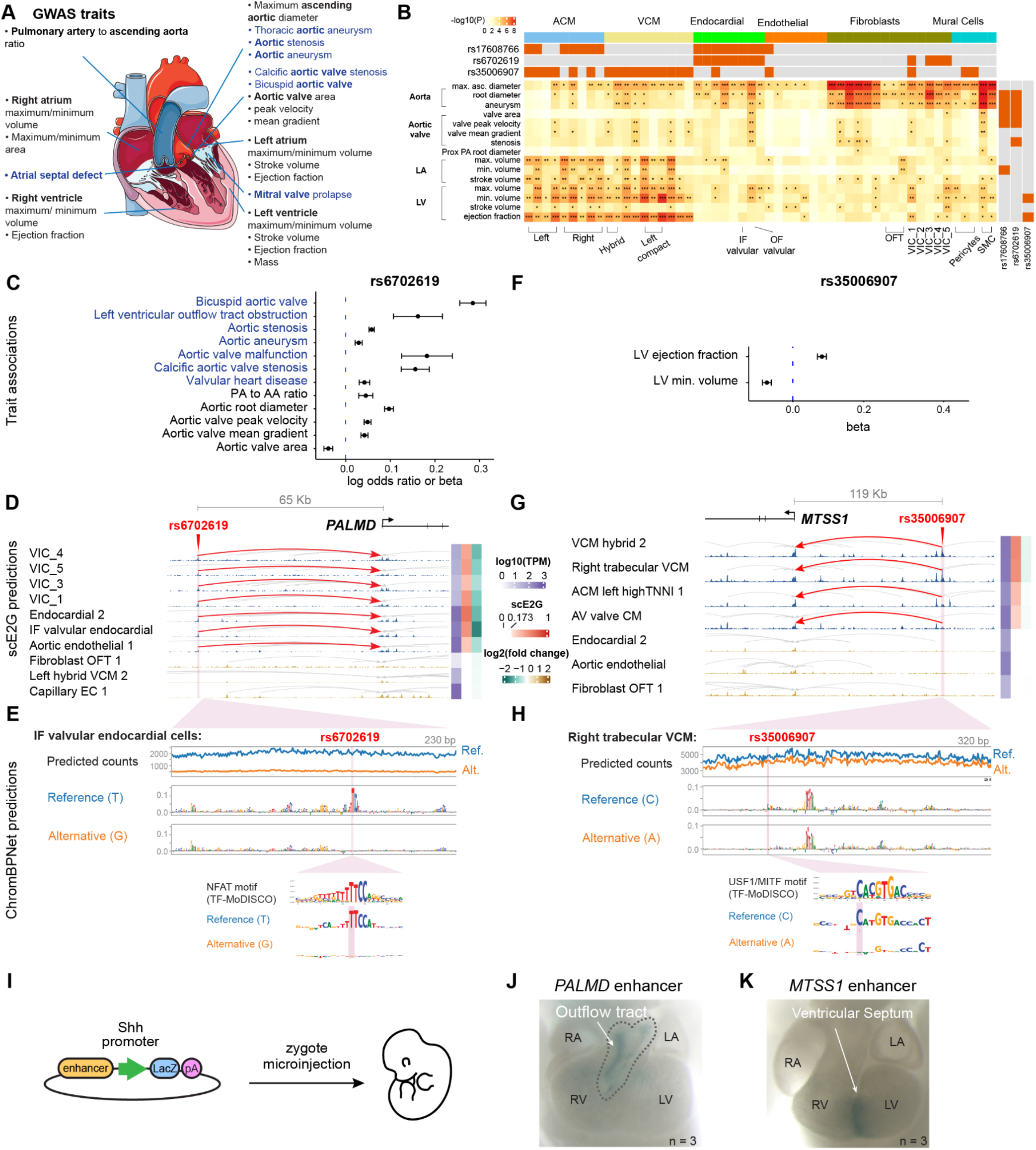
Noncoding variants affect enhancers with cell type and spatially restricted activities. **A.** Selected GWASs of quantitative measurements of the heart (gray) and cardiac diseases (blue) included in this analysis. **B.** Heritability enrichment (S-LDSC) of quantitative measurements of the heart and cardiac diseases in scE2G predicted enhancers of each cell type. Top column annotations indicate whether prioritized variants (e.g., rs17608766, rs6702619, rs35006907) overlap with an active regulatory element in the corresponding cell type. Right row annotations denote whether these specific variants are genetically associated with the respective trait. Asterisks indicate statistical significance following per-trait Bonferroni correction: * *P*adj < 0.05, *\*\* P*adj < 0.01, *** *P*adj < 0.001. Representative cell types are shown. See also Table S15. **C.** Trait associations for rs6702619. X-axis: the log odds ratio (for binary traits) or the beta per-unit change (for quantitative traits) associated with the alternative allele. Error bars represent 95% confidence intervals. The alternative allele increases the risk for aortic and aortic valve related diseases, and increases the quantitative measurement of aortic traits except for aortic valve area. See Supplementary Table S14 for data sources. **D.** rs6702619 overlaps an enhancer (hg38: chr1:99580436-99581202) predicted to regulate *PALMD* in valvular cells (red highlight and arcs). Signal tracks represent chromatin accessibility from scATAC-seq. Gray arcs represent other predicted enhancer-gene regulatory interactions in the locus. The side color bars show the TPM expression of *PALMD* in each cell type (blue), scE2G scores (red, and white for scores below the threshold), and ChromBPNet predicted log2 fold changes in chromatin accessibility upon substitution of the reference allele (T) with the alternative allele (G) (green indicates reduced accessibility, brown indicates increased accessibility). IF: inflow. **E.** ChromBPNet predictions for rs6702619 in IF valvular endocardial cells. The alternative allele is predicted to lead to a decrease in chromatin accessibility across the element (top). Middle and bottom tracks show contribution scores for the reference and alternative alleles. Inset: NFATC1 motif learned *de novo* by TF-MoDISCO, and contribution scores for the reference and alternative alleles. **F.** Trait associations for rs35006907. Beta: Per unit change in the outcome associated with the alternative allele. The alternative allele increases LV ejection fraction, and decreases the minimum LV volume. **G.** rs35006907 overlaps an enhancer (hg38: chr8:124846865-124849334) predicted to regulate *MTSS1* in VCMs (red highlight and arcs). Similar to **E.H.** ChromBPNet predictions for rs35006907 in right trabecular VCMs. The alternative allele is predicted to lead to a decrease in chromatin accessibility across the element (top). Middle and bottom tracks show contribution scores for the reference and alternative alleles. Inset: Motif learned *de novo* by TF-MoDISCO (similar to USF1 and MITF motifs), and contribution scores for the reference and alternative alleles. **I.** Schematics for the mouse transgenic assay. The predicted enhancer is cloned to drive the expression of LacZ. **J.** The enhancer overlapping rs6702619 is active in the OFT of the developing mouse heart at E11.5 (n=3). **K.** The enhancer overlapping rs35006907 is active in the interventricular septum of the mouse heart at E11.5 (n=3).

**Fig. S23:**
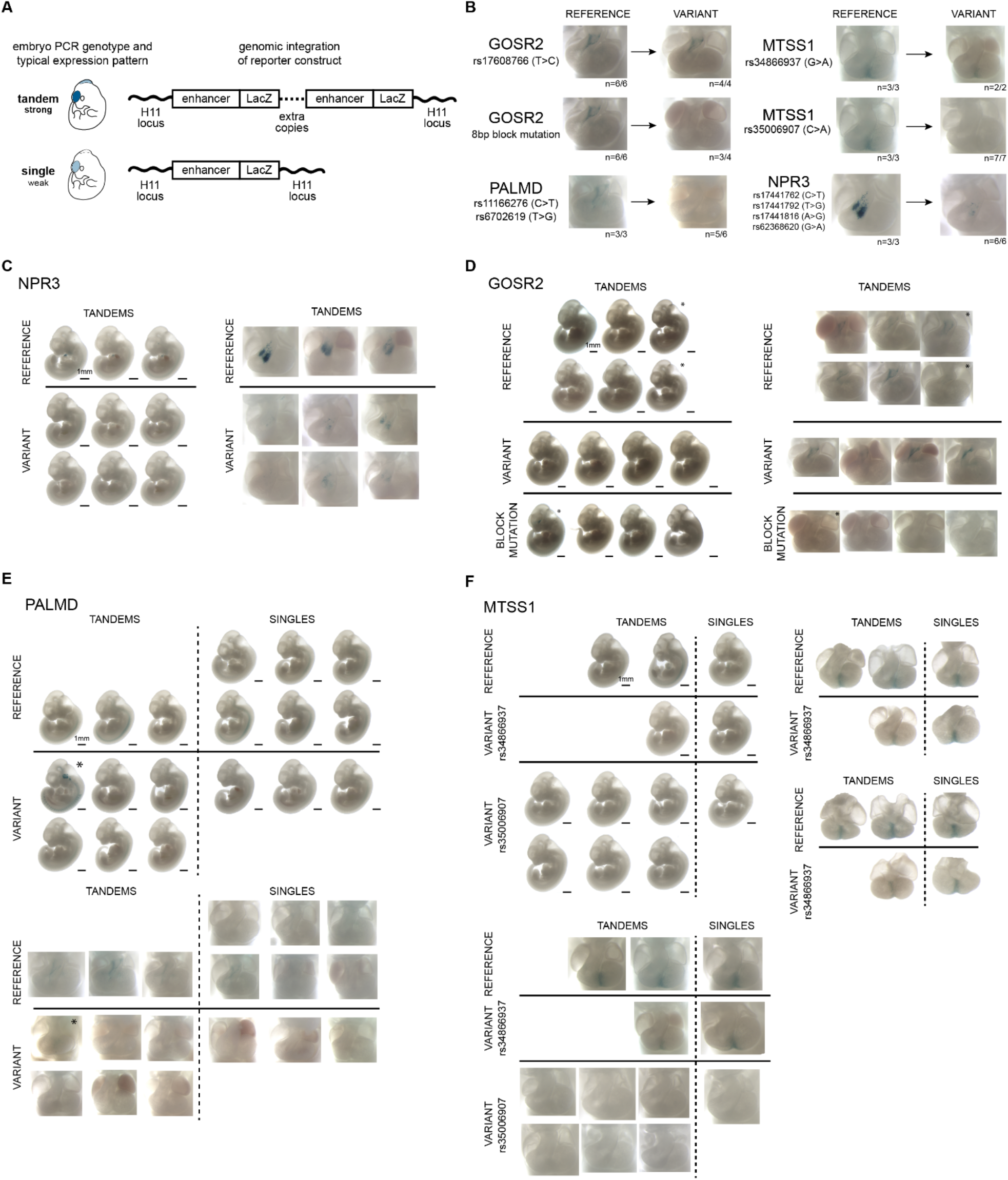
Human enhancer sequences containing risk variants have location-specific activities in transgenic enhancer reporter assay in mouse embryos. **A.** Mouse embryos injected with reporter constructs are genotyped using PCR to confirm successful integration at H11 safe harbor locus and establish approximate copy number of integration (single or tandem). Tandem embryos typically exhibit stronger staining patterns. See Methods for more details. **B.** Impact of common variants on cardiac activity of enhancer reporter in representative embryos at E11.5. Enhancers are identified by their putative target gene. Numbers indicate reproducibility with both tandems and single embryos considered for *MTSS1* elements, and only tandem embryos considered for the remaining elements^271^. *GOSR2* and *NPR3* embryos genotyped as ’singles’ exhibited no heart staining and were omitted from the dataset. Variants listed under the same enhancer ID were evaluated on a single, shared reporter construct. For *NPR3*, the lead variant (rs17441816) is in perfect linkage disequilibrium (LD) (r^2^=1, D′=1) with three additional variants, all four of which overlap the tested enhancer. **C.** Whole-embryo lateral views and frontal heart close-ups of all collected embryos with validated H11 integration. Asterisks indicate embryos exhibiting unusual ectopic staining, likely due to a random genomic integration in addition to H11 integration. Scale bars are approximate measurements based on E11.5 embryo average crown-rump length of 6 mm. *NPR3* enhancer activity observed in the conotruncal region is reduced by the introduction of risk variants. Single-integration embryos exhibited no cardiac staining and were omitted from the panels. **D.** Similar to panel C, for *GOSR2* enhancer. *GOSR2* enhancer activity is abolished by the introduction of a block mutation; positive staining is observed in four tandem reference embryos but is undetectable in three tandem variant embryos. Single-integration embryos exhibited no cardiac staining and were omitted from the panels. **E.** Similar to panel C, for *PALMD* enhancer. *PALMD* enhancer activity is reduced upon introduction of risk alleles for two linked variants (rs11166276 and rs6702619). **F.** Similar to panel C, for *MTSS1* enhancer. *MTSS1* enhancer activity is reduced by the rs35006907 risk allele, whereas the rs34866937 risk variant shows no visible effect. Front and back close-ups of isolated hearts are provided to further contrast the reference and rs34866937 risk alleles.

**Fig. S24:**
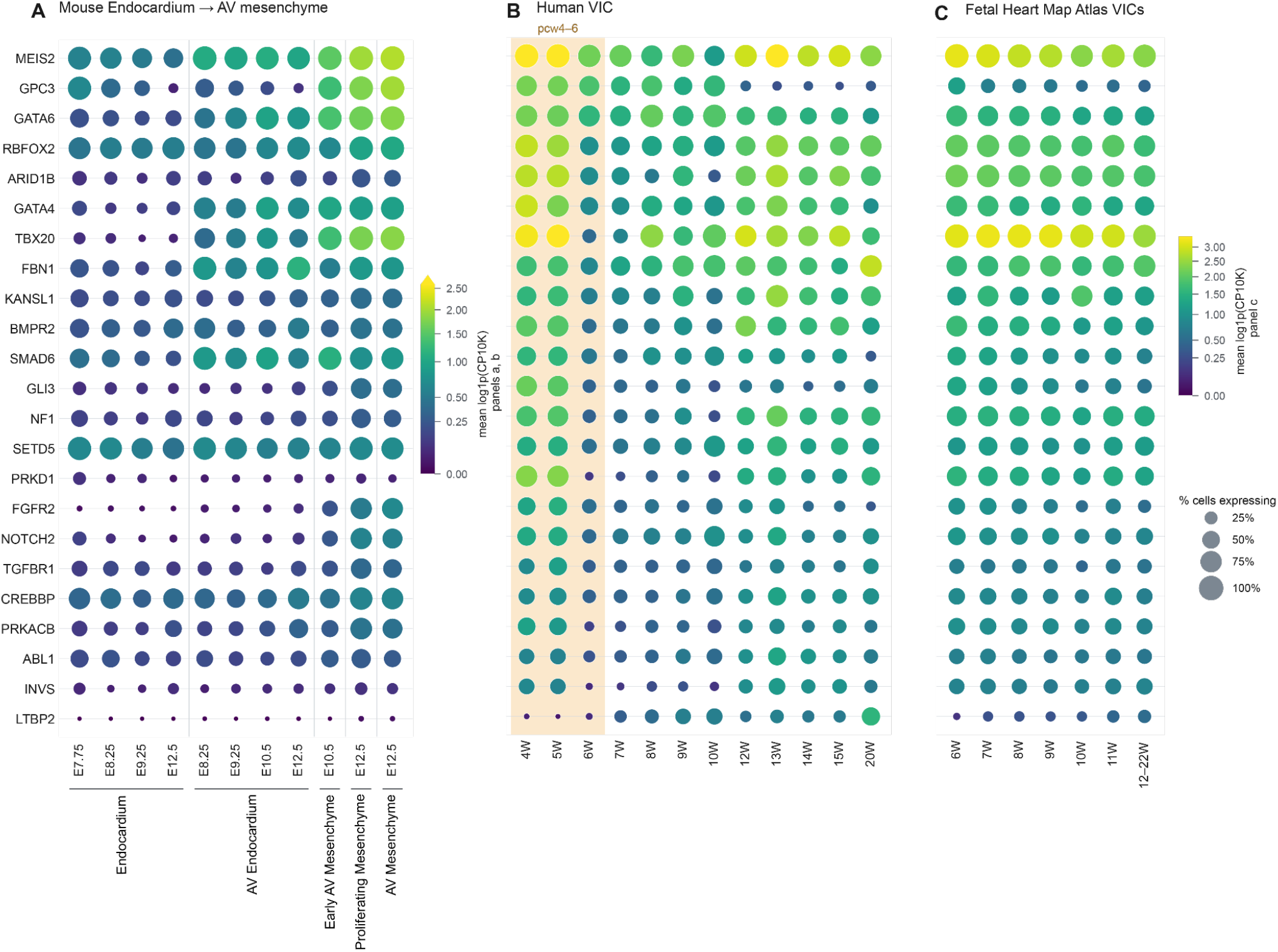
**23 high-confidence CHD genes are expressed across human and mouse VIC development.**A. Expression of the enriched high-risk CHD genes during mouse endocardial to mesenchymal transition (EndMT)^272^ from E7.75 to E12.5 (n = 8,317 cells in total). Dot color = mean absolute log1p(CP10K), dot area = % cells expressing. B. Expression of the enriched high-risk CHD genes in human VICs across 12 PCW (n = 7,398 cells). PCW4–6 is shaded.^47^ Dot color = mean absolute log1p(CP10K) shared between Panel A and B, dot area = % cells expressing. C. Expression of the enriched high-risk CHD genes in human fetal VICs across the fetal heart atlas, with 12-22 PCW pooled (n = 13,636 in total). Dot color = mean absolute log1p(CP10K) only for Panel C, dot area = % cells expressing.

## Supplementary Tables

### Table S1. List of human fetal heart tissue samples

This table contains information related to the sample attributes for a collection of heart tissues. Each row corresponds to a unique tissue sample, with details about the subject, sex, and age.

- Physical Sample: This column contains a unique identifier for each fetal tissue sample.
- Sex: Biological sex, as inferred from expression of genes on sex chromosomes (see Methods).
- Age (Quartiles): Quartile bin of developmental age in post-conception weeks (PCW).

### Table S2. 10x Multiome sample metadata

Each row corresponds to one 10x Multiome lane.

- Pooled Subject ID (dbGaP): Unique Subject IDs from dbGaP that can be used to cross reference information in this table.
- Physical Sample: Denotes individual human fetal heart samples.
- Physical Sample ID: Represents the specific tissue piece used in each experiment. The format “PhysicalSample_FT###_X” is used, where “FT###” refers to the Sample ID, and “_X” denotes the unique tissue piece from that subject. In some cases, the same heart sample was profiled in multiple experiments, and, in these cases, different physical samples of tissue from the same heart were used in each experiment.
- Lane ID: Represents a unique identifier for each 10X lane, including information about which heart samples were used to make each library. This unique identifier is the Sample ID from our dbGaP metadata.
- Experiment Number: Represents the unique identifier for each experiment conducted on a specific day. Each number corresponds to a distinct experimental setup.
- 10x Lane number: Refers to the specific lane on the loading chip used during the GEM (Gel Bead-in-Emulsion) creation process in the Chromium Controller by 10X Genomics.
- Tissue State Input: “Flash Frozen” or “Fresh”, according to whether or not the sample was frozen between tissue collection and nuclei isolation.
- Were samples mixed?: This column specifies whether the samples were mixed in the 10X lane (Yes or No).
- Mixing Ratio: This column indicates the ratio in which samples were combined in a 10X lane. For experiments with a single sample, the ratio is represented as ‘1’. When combining two, three, or four samples, the ratios are ‘1:1’, ‘1:1:1’, and ‘1:1:1:1’ respectively, reflecting equal ratio pooling.
- Region Where Tissue Was Taken: This column specifies whether the entire heart was used or the specific location from which tissue samples were collected from human fetal hearts.
- Protocol Version Used: Experimental protocol version (versions 1-3) applied to each sample during the experimental portion (see Methods). The majority of our dataset was processed with version 3.
- GEX index columns: Barcode information for the gene expression libraries (GEX), derived from 10x Multiome Dual Index Plate TT Set A (PN 3000431).
- ATAC i7_Index columns: Barcode information for the scATAC-seq libraries, derived from 10x Multiome Sample Index Plate N, Set A (PN 3000427).

**Table S3. Summary of annotation rationales and key marker genes for identified cell population.** This table provides a comprehensive overview of the gene expression evidence and spatial validation for each cell population.

- Cell_type: The name assigned to the cell population.
- Description: Detailed rationale for the specific cell type or state assignment.
- N_PCW12: Total number of cells from PCW12.
- N_MERFISH_mapped: Number of cells from the representative MERFISH heart section (R77_4C4) successfully mapped to this fetal heart cell population.
- Spatial_mapping: The distribution of the original anatomical annotation of the mapped cells from the representative MERFISH heart section (R77_4C4).
- Marker_genes: The marker genes supporting the cell type or state assignment.
- Evidence_source: Literature sources for the markers.
- Figure_reference: Cross-references to corresponding spatial mapping visualizations in the manuscript.

### Table S4: Metadata information of the cells in the fetal heart atlas

This table contains basic QC and identity information of each cell in the fetal heart atlas. This table is available at <u>syn77182533</u>

- cbc: Unique cell barcode identifier.
- orig.ident: Original sequencing library identifier.
- source: Donor identifier.
- Sex: Biological sex of the donor.
- nUMI: Number of UMIs detected per cell.
- nGene: Number of uniquely detected genes per cell.
- ratio.mt: Fraction of total UMIs mapping to mitochondrial genes.
- ratio.ribo: Fraction of total UMIs mapping to ribosomal protein genes.
- nUMI_non_MT: Total number of non-mitochondrial UMIs per cell.
- log10GenesPerUMI: log10Genes/log10UMI.
- GEX_saturation: Sequencing saturation percentage for gene expression libraries.
- nFrags: Total number of scATAC-seq nuclear fragments per cell.
- TSSEnrichment: Transcription start site (TSS) enrichment score for chromatin accessibility quality.
- S.Score: Calculated S-phase gene expression score.
- G2M.Score: Calculated G2\/M\-phase gene expression score.
- Phase: Predicted cell cycle phase.
- celltype: Cell type annotation.
- Plot_label: Cell type group label.
- HexColorCode: Color assigned to the corresponding cluster in Fig. 1C.
- UMAP_1: Cell coordinate along UMAP dimension 1.
- UMAP_2: Cell coordinate along UMAP dimension 2.
- PCW_quartile: Quartile bin of developmental age in post-conception weeks (PCW).

### Table S5: Enhancer-gene regulatory interactions identified for each cell type in the heart map

In this table, each row represents the effect of one element on one nearby gene in one cell type, as predicted by scE2G. This table is available at https://www.synapse.org/Synapse:syn64109184.

- chr: chromosome for element-gene pair
- start: start coordinate for element (hg38)
- end: end coordinate for element (hg38)
- name: unique identifier for element
- class: category describing element location, one of: “promoter,” “intergenic,” “genic”
- TargetGene: HGNC symbol for target gene of element-gene pair
- TargetGeneTSS: transcription start site of target gene
- TargetGeneIsExpressed: boolean indicating whether the target gene is expressed based on promoter accessibility, should be “True” for all predictions
- TargetGeneEnsembl_ID: Ensembl ID corresponding to target gene
- isSelfPromoter: boolean indicating whether this element-gene pair is a promoter linked to its own gene
- distance: distance between the midpoint of element and transcription start site of target gene
- normalizedATAC_prom: (model feature) normalized pseudobulk scATAC-seq signal at the promoter of the target gene
- contactFrequency: the three-dimensional physical contact frequency between an enhancer and a gene, based on a megamap (cell-type average) of Hi-C
- ABC.Score: Activity-by-Contact (ABC) score corresponding to this element-gene pair, calculated using pseudobulk scATAC-seq for activity and a distance-based power law estimation for contact
- normalizedATAC_enh: (model feature) normalized pseudobulk scATAC-seq signal at the enhancer
- numTSSEnhGene: (model feature) number of other transcription start sites between this element and the target gene transcription start site
- numNearbyEnhancers: (model feature) number of elements within 5 kb of this element
- ubiqExpressed: (model feature) boolean indicating whether this gene is classified as ubiquitously expressed across cell types
- RNA_meanLogNorm: mean log-normalized read count for this gene in this cell cluster
- RNA_pseudobulkTPM: pseudobulk transcripts per million for this target gene for this cell cluster
- RNA_percentCellsDetected: percent of cells in this cell cluster in which this gene was detected
- Kendall: Kendall correlation between accessibility at this element and expression of this gene across cells in this cluster
- ARC.E2G.Score: (model feature) Activity, Responsiveness and Contact (ARC)-E2G score that integrates ABC score and Kendall correlation
- E2G.Score: raw scE2G score directly output from logistic regression model
- E2G.Score.qnorm: Final score column used for thresholding, after quantile normalization to full-depth reference scores and setting scores for genes below TPM threshold to 0
- E2G.Score.qnorm.ignoreTPM: same as E2G.Score.qnorm, but without scores for genes with TPM below threshold set to 0

#### ● CellType: description of cell cluster for these predictions

### Table S6: Gene programs: cell type grouping for cNMF

- CellType: cell type name.
- NMFGroup: cell type group that the cell type was assigned to for cNMF analysis. The cell types grouped together are phenotypically similar.

### Table S7: Gene programs: 300 top genes expressed in each program

Each column corresponds to one gene program. Rows correspond to the 300 top genes ranked by importance scores in the program.

### Table S8: Gene programs: TFs predicted to regulate each program by chromVAR

- Program: Name of the gene program
- TF: Name of the transcription factor (TF)
- CellNumber: Number of cells expressing the gene program
- P: P-value of the Spearman correlation between per-cell motif accessibility of the TF and the expression level of the gene program
- Estimate: Spearman’s rank correlation coefficient
- P.adjust_bonferroni: Adjusted p-value after Bonferroni correction for multiple comparisons.
- TF_rank_in_Program: Rank of the TF in the gene program, with lower ranks indicating the expression of the TF is more highly associated with the expression of the gene program.

### Table S9: Significant ligand-receptor pairs between sender and receiver cell types in cell-cell interaction analysis

This data file is available at https://www.synapse.org/Synapse:syn64108840

- source: Source cell type, where molecular signals are sent from.
- target: Target cell type, where molecular signals are sent to.
- ligand: Ligand gene name, note that multiple subunits of the protein are separated with an underscore. The ligand list comes from CellChat v2.1.2 database.
- receptor: Receptor gene name, note that multiple subunits of the protein are separated with an underscore. The receptor list comes from CellChat v2.1.2 database.
- prob: Cell-cell interaction strength. Calculated with R package CellChat v2.1.2.
- pval: Permutation-based p-value for the interaction strength. Calculated with R package CellChat v2.1.2.
- interaction_name: Cell-cell interaction name.
- interaction_name_2: Cell-cell interaction name, format 2.
- pathway_name: Name of the pathway that the ligand-receptor pairs belong to. The membership comes from the CellChat v2.1.2 database.
- annotation: Type of the ligand-receptor pair, e.g. secreted signaling, Cell-Cell Contact, etc.
- evidence: Supporting information for the ligand-receptor pair, e.g. PubMed ID, KEGG ID, etc. The information comes from the CellChat v2.1.2 database.
- PCW: Post-conception weeks. The cell-cell interaction analysis was performed one week at a time.

### Table S10: Validated scE2G predicted enhancers

This data file is available at https://www.synapse.org/Synapse:syn64729142

- Fetal_heart_enhancer_chromosome: chromosome of enhancer in the heart map
- Fetal_heart_enhancer_start: start coordinate of enhancer in the heart map
- Fetal_heart_enhancer_end: end coordinate of enhancer in the heart map
- Fetal_heart_enhancer_cell_type: cell type of enhancer in the heart map
- Target_gene_if_applicable: target gene of enhancer if the enhancer-gene regulatory interaction was validated in the experiment
- Validated_enhancer_chromosome: chromosome of enhancer in the validation experiment
- Validated_enhancer_start: start coordinate of enhancer in the validation experiment
- Validated_enhancer_end: end coordinate of enhancer in the validation experiment
- Validated_enhancer_ID: ID of enhancer in the validation experiment
- Experimental_system: experimental system used for validation
- Overlap_base_pairs: number of base pairs overlapping between enhancer in the heart map and the validated enhancer
- PMID: PMID of the validation experiment

### Table S11: Curated list of CHD genes, genes with a current *de novo* variant burden, and other cardiovascular disease genes

Each row represents a unique gene-trait pair. A gene can appear in multiple rows if it is linked to several different traits.

- Gene: Gene name.
- Trait: Name of the disease.
- Category: The broad category of the disease genes. CHD: high-confidence genes from CHDGene; DeNovo_Transmitted_Damaging_Burden: CHD genes with a significant burden of monoallelic damaging variants, driven by both *de novo* and transmitted mutations; CardiovascularDisease: other cardiovascular disease genes (see **Methods**).

### Table S12: Enrichment of high-confidence CHD genes and other cardiovascular disease genes in cardiac cell types

- Category: The broad category of the disease genes. CHD: High-confidence CHD genes from CHDgene. CD: other cardiovascular disease genes.
- Disease: Name of the disease.
- n_genes_highly_expressed_in_Celltype_and_are_Disease_genes: Number of disease genes that are highly expressed in the cell type (see Methods)
- n_genes_highly_expressed_in_Celltype_and_are_NOT_Disease_genes: Number of non-disease genes that are highly expressed in the cell type
- n_genes_NOT_highly_expressed_in_Celltype_and_are_Disease_genes: Number of disease genes that are not highly expressed in the cell type.
- n_genes_NOT_highly_expressed_in_Celltype_and_are_NOT_Disease_genes: Number of non-disease genes that are not highly expressed in the cell type.
- odds_ratio: the enrichment of disease genes in the highly expressed genes of the cell type.
- CI_lower: The low bound of the 95% confidence interval of the odds ratio.
- CI_upper: The higher bound of the 95% confidence interval of the odds ratio.
- P_value: The *p*-value of the odds ratio.
- NumberOfTrueDiseaseGenes: Total number of known causal genes for the disease.
- recall: Percentage of disease genes that are highly expressed in the cell type.
- disease_genes_in_Celltype: List of disease genes highly expressed in the cell type.
- P_bonferroni: Adjusted p-value after correcting for multiple testing using the Bonferroni method.
- Celltype: Name of the cell type.

### Table S13. Evidence for 27 high-confidence CHD genes that are highly expressed in VICs and/or cardiac fibroblasts

Summary of evidence presented in **Note S3** regarding 27 high-confidence CHD genes that are highly expressed in VICs and/or cardiac fibroblasts. Each row represents a unique gene.

- Gene: Gene symbol.
- CHD Subtype(s): Subtype(s) of CHD associated with each gene (see Methods).
- Highly expressed in VIC (Yes/No): Indicates whether each gene is highly expressed in any of the 5 populations of VICs
- Highly expressed in cardiac fibroblasts (Yes/No): Whether the gene is highly expressed in Cardiac_fibroblast_progenitors.
- Knockout evidence in epicardial lineage (Yes/No): “Yes” indicates that gene knockout in the epicardial lineage led to developmental changes or defects. “No” indicates either a lack of impact from knockout experiments or that no knockout studies were conducted for this lineage to our knowledge.
- Knockout evidence in endocardial lineage (Yes/No): “Yes” indicates that gene knockout in the endocardial lineage led to developmental changes or defects. “No” indicates either a lack of impact from knockout experiments or that no knockout studies were conducted for this lineage to our knowledge.
- Knockout evidence in neural crest lineage (Yes/No): “Yes” indicates that gene knockout in the neural crest lineage led to developmental changes or defects. “No” indicates either a lack of impact from knockout experiments or that no knockout studies were conducted for this lineage to our knowledge.

### Table S14: List of GWAS traits and data sources

- Trait: Name of the trait in **Table S15** and **Table S16**. Quantitative traits with duplicate entries reflect alternative normalizations: covariate-adjusted joint modeling (“invnorm”; used for fine-mapping and variant overlapping) versus inverse-normal transformation of covariate residuals (“resint”).
- DescriptiveName: Description of the trait.
- Citation: PMID or zenodo ID of the corresponding GWAS.
- NumberOfLoci: Number of independent genome-wide significant loci or fine-mapped credible sets (if applicable).
- VariantProcessingMethod: Methods used for preparing variants for overlap analysis with scE2G-predicted enhancers (see **Methods**)
- LDSC: Whether the trait was included in the LDSC analysis.
- VariantPrioritization: Whether the trait was included in the variant prioritization analysis.
- ValveTraits: Valve-related traits analyzed in Fig. 3.

### Table S15: Heritability enrichment (S-LDSC) for GWAS traits in enhancers in each cell type

- Celltype: Name of the cell type.
- major_celltype: Broad category classification of the cell type
- Enrichment: The enrichment of the heritability of the trait in the enhancers of this cell type.
- Enrichment_std_error: Standard error of the enrichment estimate
- Enrichment_p: P-value for the significance of the enrichment
- Enrichment_p_bonferroni: P-value adjusted for multiple comparisons with Bonferroni correction across all annotations, including those in the baseline model.
- Coefficient_z.score: Z-score of the regression coefficient which measures the additional contribution of this cell type’s enhancers to the model
- Trait: Name of the trait.
- n_LeadSNPs: Number of lead SNPs associated with the trait.

### Table S16: Prioritized variants, enhancers, genes, and gene programs for GWAS signals for heart diseases and traits

Each row provides details about an scE2G predicted cell type-specific enhancer-gene link overlapped by a variant (listed in the “rsid” column) associated with a GWAS signal (listed in the “LeadVariant” column). Columns:

- enh_chr: Chromosome ID of the enhancer.
- enh_start: Start coordinate of the enhancer (hg38)
- enh_end: End coordinate of the enhancer (hg38)
- normalizedATAC_enh: (model feature) normalized pseudobulk scATAC-seq signal at the enhancer
- Trait: Name of the trait.
- DescriptiveName: Description of the trait.
- nLeadSNPs: Number of lead SNPs (credible sets) for the trait.
- CredibleSet: ID of the credible set.
- rsid: ID of the variant.
- distance: Distance between the enhancer and the promoter of the target gene.
- E2G.Score.qnorm: Quantile-normalized enhancer-to-gene (E2G) interaction score
- TargetGene: Target gene regulated by the enhancer.
- TargetGeneRank: Rank of E2G.Score.qnorm of the target gene among all target genes regulated by the same enhancer in this cell type (lower rank indicates higher E2G.Score.qnorm)
- AnyVariantInCSOverlapCoding: Whether any variant in the credible set (CS) overlaps a coding region.
- AnyVariantInCSOverlapSpliceSite: Whether any variant in the credible set (CS) overlaps a splice site.
- ProgramsContainingTargetGene: Gene programs that the target gene is involved in
- TargetGeneRankInProgram: Rank of the target gene within the gene program (lower rank indicates stronger association)
- ProportionOfProgramExpressingInCelltype: Proportion of cells expressing the gene program that belong to this cell type.
- TFsOfProgram: Transcription factors (TFs) predicted to regulate the gene program
- TargetGeneCausesAdultCardiovascularDisease: The adult cardiovascular diseases known to be caused by the target gene.
- TargetGeneCausesCHD_CHDgene: The CHD subtypes known to be caused by the target gene.
- TargetGeneCausesCHD_VariantBurden: Indicates whether the target gene carries a significant burden of heterozygous *de novo* or rare transmitted damaging variants.
- LeadVariant: Variant ID of the lead variant of the credible set.
- LeadVariant*P*value: P-value of the lead variant from the GWAS
- variant*P*value: P-value of the variant from the GWAS (if available)
- LeadSNP_PosteriorProb: Posterior probability of the lead SNP (if available)
- PosteriorProb: Posterior probability of the variant (if available)
- TPM: Transcripts Per Million (TPM) of the target gene
- Celltype: Cell type in which the enhancer-gene interaction occurs

### Table S17: Genomic sequences tested in mouse transgenic enhancer reporter assay

- VISTA ID: ID of the enhancer in the VISTA database
- target gene: Target gene predicted to be regulated by the enhancer
- coordinates (hg38): Genomic coordinates of the human enhancer sequence
- reference sequence: Reference DNA sequence cloned for testing the activity of the enhancer.
- variant sequence: DNA sequence with the variant(s) introduced for testing their impacts on the activity of the enhancer.
- variant rsID: IDs of the variant(s)

### Table S18: TF-MoDISco motifs

Each row represents one motif identified in one cell type by ChromBPNet and TF-MoDISco. This data file is available at: https://www.synapse.org/Synapse:syn65000518

- pattern: Pattern name
- num_seqlets: Number of seqlets supporting the pattern
- match0: Best match for the pattern in the reference motif database
- qva0: q-value of the best match
- match1: Second best match in the reference motif database
- qval1: q-value for match1
- match2: Third best match
- qval2: q-value for match2
- celltype: Cell type that the patterns were identified for

### Table S19: TF-MoDISco motif instances identified with FiNeMo

Each row represents one motif instance (*e.g.*, candidate TF binding site) in one cell type identified by ChromBPNet, TF-MoDISco, and FiNeMo. This data file is available at: https://www.synapse.org/Synapse:syn64109173

- chr, start, end: Chromosome, start and end of the match (trimmed).
- start_untrimmed,end_untrimmed: Untrimmed start and end of the match (30 bp).
- motif_name: The name of the pattern in the TF-MoDISco table for the corresponding cell type.
- hit_coefficient: The regression coefficient for the hit, normalized per peak region.
- hit_coefficient_global: The regression coefficient for the hit, scaled by the overall importance of the region.
- hit_correlation: The correlation between the untrimmed CWM and the contribution score of the motif hit.
- hit_importance: The total absolute contribution score within the motif hit.
- strand: The orientation of the hit (+ or -).
- peak_id: The numerical index of the peak region containing the hit in the input file for the respective cell type.
- celltype: Cell type that the scanning was performed in.

### Table S20: ChromBPNet performance metrics

Each row corresponds to one cell type. This data file is available at: https://www.synapse.org/Synapse:syn65000540

- celltype: Cell type that the models were trained for
- counts_pearsonr_mean: Pearson correlation coefficient of observed and predicted log(total counts) mean across five folds.
- counts_spearmanr_mean: Spearman correlation coefficient of observed and predicted log(total counts), mean across five folds.
- median_jsd_mean: Median Jensen-Shannon Distance (JSD) between the normalized base-resolution coverage profile and the predicted multinomial profile distribution, mean across five folds.
- median_norm_jsd_mean: Median of the min-max normalized JSD, mean across five folds.
- counts_pearsonr_std: Standard deviation across five folds for counts_pearsonr.
- counts_spearmanr_std: Standard deviation across five folds for counts_spearmanr.
- median_jsd_std: Standard deviation across five folds for median_jsd.
- median_norm_jsd_std: Standard deviation across five folds for median_norm_jsd.

### Table S21: sgRNA guide sequences for targeting *KDR* enhancer

- Name: Unique identifier for sgRNA
- Sequences: Sequence of sgRNA
- Target: Target of sgRNA
- PassedQC: boolean indicating whether the sgRNA containing cells passed QC

### Table S22: pegRNA sequences targeting the GOSR2 variant locus

- chr, variantStart, variantEnd: Genomic coordinates of the target edit locus (all coordinates in hg19).
- variantName: Unique identifier of the edit, including reference (ref) and alternative (alt) sequences.
- ref, alt: Reference and alternative sequences for the edit.
- guideStart, guideEnd, guideStrand: Genomic coordinates and strand orientation (+ or -) of the guide RNA.
- pbsStart, pbsEnd, pbsLength, primerBindingSite: Genomic coordinates, length, and sequence of the primer binding site (PBS).
- rtTemplateStart, rtTemplateEnd, rtTemplateLength, rtLengthPastEdit, rtTemplate: Genomic coordinates, length, extension length past the edit site, and sequence of the reverse transcriptase (RT) template.
- Spacer: Sequence of the gRNA spacer used
- editPositionRelativeToNick0Based, editOverlapsPam: 0-based distance of the edit relative to the nick site, and indicator of whether the edit overlaps the PAM.
- pegRNASequence: Complete prime editing guide RNA sequence.

### Table S23: Variant-EFFECTS measured values of ten pooled genomic variants

- **Variant ID:** Genomic locus and the specific edit introduced.
- **Frequency (%):** Relative frequency of the edit within the analyzed cell population.
- **Effect Size (%):** Measured impact of the variant on *GOSR2* expression levels.
- **Corrected *P*-value:** Statistical significance of the measured expression effect, adjusted for multiple comparisons.

